# An Interactive Trustworthy AI Pathology Copilot to Improve Biomarker-Driven Prognostic Stratification and Therapeutic Response Prediction

**DOI:** 10.64898/2026.05.17.26352870

**Authors:** Yixiao Mao, Chengjie Xie, Feng Li, Danyi Li, Wenyan Zhang, Yidan Zhang, Bingbing Li, Chenglong Zhao, Zhengyu Zhang, Ying Tan, Zhijian Cen, Haisu Tao, Jian Yang, Jian Wang, Qianjin Feng, Boxiang Liu, Li Liang, Cheng Lu, Yu Zhang, Zhenyuan Ning

**Author notes:** Corresponding authors (ZY. Ning), (Y. Zhang), (C. Lu), (L. Liang), and (B. Liu). These authors have contributed equally to this work and share first authorship (Yixiao Mao, Chengjie Xie).

## Abstract

Predictive assays for precision oncology increasingly rely on multi-scale biomarkers that manifest as morphological signatures in routine whole-slide images (WSIs). However, most computational pathology models treat biomarker profiling and outcome prediction (i.e., prognostic stratification and therapeutic response) as independent tasks, and lack the interactive and trustworthy capabilities required for clinical translation. Here, we present TEAM, an interactive trustworthy AI pathology copilot that improves biomarker-driven outcome prediction. Pretrained on 55,648 pan-cancer WSIs and 1,750,648 regions of interest (ROIs), comprising 360 million patches, TEAM learns risk-aware embeddings by conditioning on clinical metadata and aligning with the relative risk prior. Patch-level data (aleatoric) and model (epistemic) uncertainty are quantified for trustworthy assessment and then propagated to slide-level predictions. In outcome prediction, TEAM uses profiled biomarkers as intermediate features to contextualize prognostic and therapeutic estimates. Beyond passive prediction, we integrate vision-language models with agentic orchestration for clinical reasoning, and provide a web-based clinician-in-the-loop interface for interactive prediction refinement. TEAM outperforms existing methods on 88 benchmarks spanning 51 multi-institutional cohorts across biomarker profiling, prognostic stratification, and therapeutic response prediction, supporting trustworthy AI-assisted decision-making in computational pathology.

## Introduction

Advances in precision oncology have led to growing demand for predictive assays that quantify specific biomarkers to inform clinical outcomes, enabling risk-adapted management and individualized therapy [1]. Such biomarkers have evolved beyond single-omics readouts to encompass cancer stage/grade, tumor microenvironment (TME), and gene expression [2]. Despite these advances, many multi-scale biomarkers rely on costly and technically demanding assays and still exhibit limited sensitivity, specificity, and clinical reproducibility [3]. These biomarkers, however, reflect cellular processes that can be characterized by distinctive morphological patterns in H&E-stained tissues. The routine digitization of whole-slide images (WSIs) has enabled artificial intelligence in computational pathology to automatically and effectively mine such patterns for biomarker profiling and outcome prediction (i.e., prognostic stratification and therapeutic response) [4, 5].

Recent computational pathology methods, from task-specific models [6, 7, 8, 9, 10] to large-scale foundation models [11, 12, 13, 14, 15], have substantially advanced both biomarker profiling and outcome prediction. However, these methods typically decouple biomarker profiling from outcome prediction, treating them as parallel tasks rather than biologically coupled processes. This decoupling overlooks the clinical reality that patient outcomes are fundamentally driven by multi-scale biological processes spanning disease extent, microenvironmental composition, and molecular programming [16, 17, 18], leaving outcome predictions detached from the biomarker context needed for clinical interpretation and decision-making.

Beyond this gap, three further barriers hinder the clinical translation of existing approaches. First, the pretraining objectives of foundation models remain morphology-centric without explicit alignment to the relative risk prior, constraining performance on outcome-related tasks. Second, existing approaches often prioritize model performance over trustworthiness, offering limited uncertainty quantification to flag cases that may warrant additional clinical scrutiny [19]. Finally, pathology foundation models are typically used as static prediction engines, with limited support for interpretable clinical reasoning or interactive human-in-the-loop refinement, restricting their clinical use to accept-or-reject decisions rather than enabling human-AI collaboration [20]. Collectively, these barriers highlight the need for a risk-aware, uncertainty-quantified, and interactive AI system for biomarker-driven outcome prediction.

Here, we present TEAM (Trustworthy intEractive risk-Aware Model), an interactive trustworthy AI pathology copilot that improves biomarker-driven outcome prediction in computational pathology. Pretrained on 55,648 pan-cancer WSIs and 1,750,648 regions of interest (ROIs), comprising 360 million patches (360,786,410; Fig. 1a), TEAM learns risk-aware pathology features by integrating clinical metadata (i.e., age, sex, race, and ethnicity) and aligning with the relative risk prior across cancer types. We then quantify both data (aleatoric) and model (epistemic) uncertainty of these features to ensure model reliability [21]. In outcome prediction, TEAM uses profiled biomarkers (cancer stage/grade, TME characterization, and gene expression) as intermediate features that contextualize downstream prognostic stratification and therapeutic response [22]. To translate model outputs into clinically actionable insights, we integrate large language models (LLMs) for knowledge-based clinical reasoning and further incorporate an agentic architecture that dynamically orchestrates biomarker profiling to deliver outcome-driven clinical recommendations. Additionally, a clinician-in-the-loop interface allows clinicians to adjust the predictions through region-specific feedback delivered via a web-based platform, enabling real-time oversight and iterative result refinement. TEAM is evaluated on 88 benchmarks (70 public and 18 in-house sets) across biomarker profiling, prognostic stratification, and therapeutic response prediction, spanning 51 multi-institutional cohorts with 30,313 WSIs and 848,655 ROIs (over 100 million patches; Fig. 1a). Results demonstrate performance advantages over existing methods and support further evaluation of TEAM as an AI-assisted pathology framework.

**Fig. 1.**
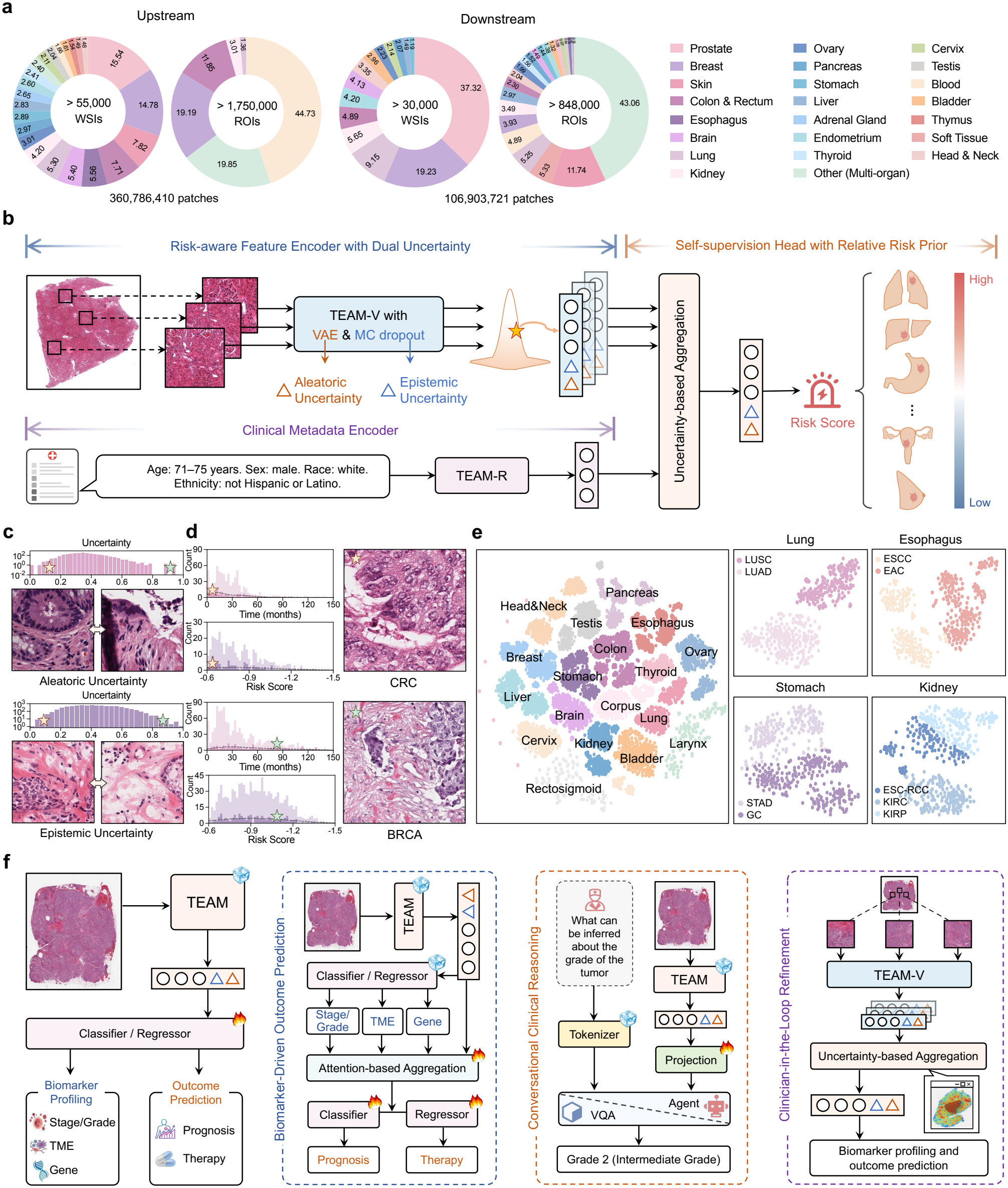
Overview of TEAM. TEAM is a trustworthy and risk-aware computational pathology copilot that couples large-scale pathology image pretraining with a flexible downstream pipeline for biomarker profiling and outcome prediction. **a**, Cohorts for pretraining and downstream analyses comprising WSIs and ROIs from multiple tissue systems and tumor types. **b**, Model architecture including (i) Risk-aware Feature Encoder with Dual Uncertainty, (ii) Clinical Metadata Encoder integrating demographic variables, and (iii) Self-supervision Head with Relative Risk Prior generating calibrated risk scores. **c**, Distributions of aleatoric and epistemic uncertainty across all WSI patches with representative examples from regions of low and high uncertainty. **d**, Distributions of overall survival (OS) time and TEAM-predicted risk scores for TCGA-CRC and TCGA-BRCA, with representative high- and low-risk patches illustrating corresponding morphological patterns. **e**, TEAM-derived embeddings of TCGA samples visualized by t-SNE showing clustering by cancer type and intra-organ subtype. **f**, Downstream use of TEAM embeddings in four settings. Biomarker profiling covers cancer staging/grading, tumor microenvironment (TME) characterization, and gene expression prediction, and outcome prediction covers prognostic stratification and therapeutic response prediction.

## Results

### Learning trustworthy risk-aware representations from whole-slide images

TEAM serves as an AI copilot for biomarker-driven outcome prediction, identifying risk-relevant patterns and providing trustworthy estimates. The overall architecture (Fig. 1b) consists of three core modules: Risk-aware Feature Encoder with Dual Uncertainty, Clinical Metadata Encoder, and Self-supervision Head with Relative Risk Prior. Each WSI is partitioned into non-overlapping patches, and the Risk-aware Feature Encoder extracts patch-level features using a DINOv2 vision transformer [23] initialized with UNI weights [11]. These features are then processed to quantify their uncertainty based on Bayesian principles [21], with a variational autoencoder (VAE) for data (aleatoric) uncertainty and Monte Carlo dropout for model (epistemic) uncertainty. In parallel, the Clinical Metadata Encoder (TEAM-R) encodes four demographic variables (i.e., age, sex, race, and ethnicity) using BioBERT [24] and is used only during pretraining, with no clinical metadata provided during downstream biomarker profiling or outcome prediction. Patch-level embeddings are then combined through uncertainty-weighted aggregation and fused with metadata embeddings to obtain a slide-level representation, which is projected into a risk score constrained by a relaxed ranking function to align with the relative risk prior while allowing for individual variability.

As visualized in Fig. 1c, TEAM captures uncertainty patterns associated with histopathological features: low data uncertainty corresponds to well-defined morphology, while high data uncertainty occurs where cells border open spaces; low model uncertainty corresponds to compact, densely nucleated regions, whereas high model uncertainty occurs in areas with sparse nuclei. TEAM-predicted risk scores capture the inter-cancer risk hierarchy (Fig. 1d), assigning significantly higher risk to CRC than to BRCA (CRC: ™0.58 ± 0.61 vs BRCA: −0.84 ± 0.41, *P*<0.001), consistent with the worse overall survival (OS) observed in CRC patients (*P*<0.001). Visualization of the pan-cancer landscape using t-distributed stochastic neighbor embedding (t-SNE) [25] (Fig. 1e) shows well-separated clusters by organ, with further segregation into histological subtypes (e.g., LUAD versus LUSC), suggesting that TEAM preserves biologically meaningful tissue architecture while encoding risk information. This organization is preserved when TEAM is pretrained without TEAM-R (TEAM w/o R) but absent when only the four demographic variables are used (Meta-only; Supplementary Fig. 1). Building on these representations, we evaluated TEAM across 88 downstream benchmarks (Fig. 1f), both as a direct predictor for outcome prediction and biomarker profiling, and as a clinical copilot for biomarker-driven outcome modeling, conversational clinical reasoning, and clinician-in-the-loop refinement.

### Accurate and robust outcome prediction via pan-cancer analysis

#### Prognostic stratification

We assessed TEAM’s prognostic performance across 31 cohorts spanning 13 cancer types (Supplementary Tables 11–12). TEAM outperforms baseline models across cohorts (Fig. 2a; Supplementary Tables 26–56), with a mean paired concordance index (C-index) improvement of 0.033 over the best-performing baseline in each cohort (95% CI, 0.024–0.042; *P*=1.17 × 10^−6^, two-sided Wilcoxon signed-rank test; Extended Data Fig. 9a). This advantage is retained when slide-level predictions are aggregated per patient (Extended Data Fig. 9b). Representative analysis of the full NFH-CESC cohort reveals significant risk stratification (*P*<0.001; Fig. 2b, top), with time-dependent receiver operating characteristic (ROC) analysis showing higher areas under the curve (AUCs) than the baselines at 1, 3, and 5 years (0.760, 0.831, and 0.944; Fig. 2b, bottom). Consistent results are observed across additional external cohorts (Extended Data Fig. 1). t-SNE visualization of TEAM embeddings from the NFH-CESC cohort (Fig. 2c) shows biologically coherent risk stratification. Among cases with observed events, predicted risk scores are negatively correlated with observed OS time (Spearman’s *ρ* = −0.75, *P*<0.001), and positions along the risk gradient are correlated with predicted risk scores (*ρ* = 0.95, *P*<0.001) and negatively with observed OS time (*ρ* = −0.70, *P*<0.001). Removing TEAM-R during pretraining lowers the mean C-index from 0.701 to 0.686 (TEAM w/o R), which remains higher than the baselines (0.622–0.645), whereas a model using only the four demographic variables reaches 0.525 (Meta-only; Extended Data Fig. 8). A decrease in C-index across the prognostic cohorts is also observed when the relative risk prior is removed (TEAM w/o prior; Supplementary Fig. 3).

**Fig. 2.**
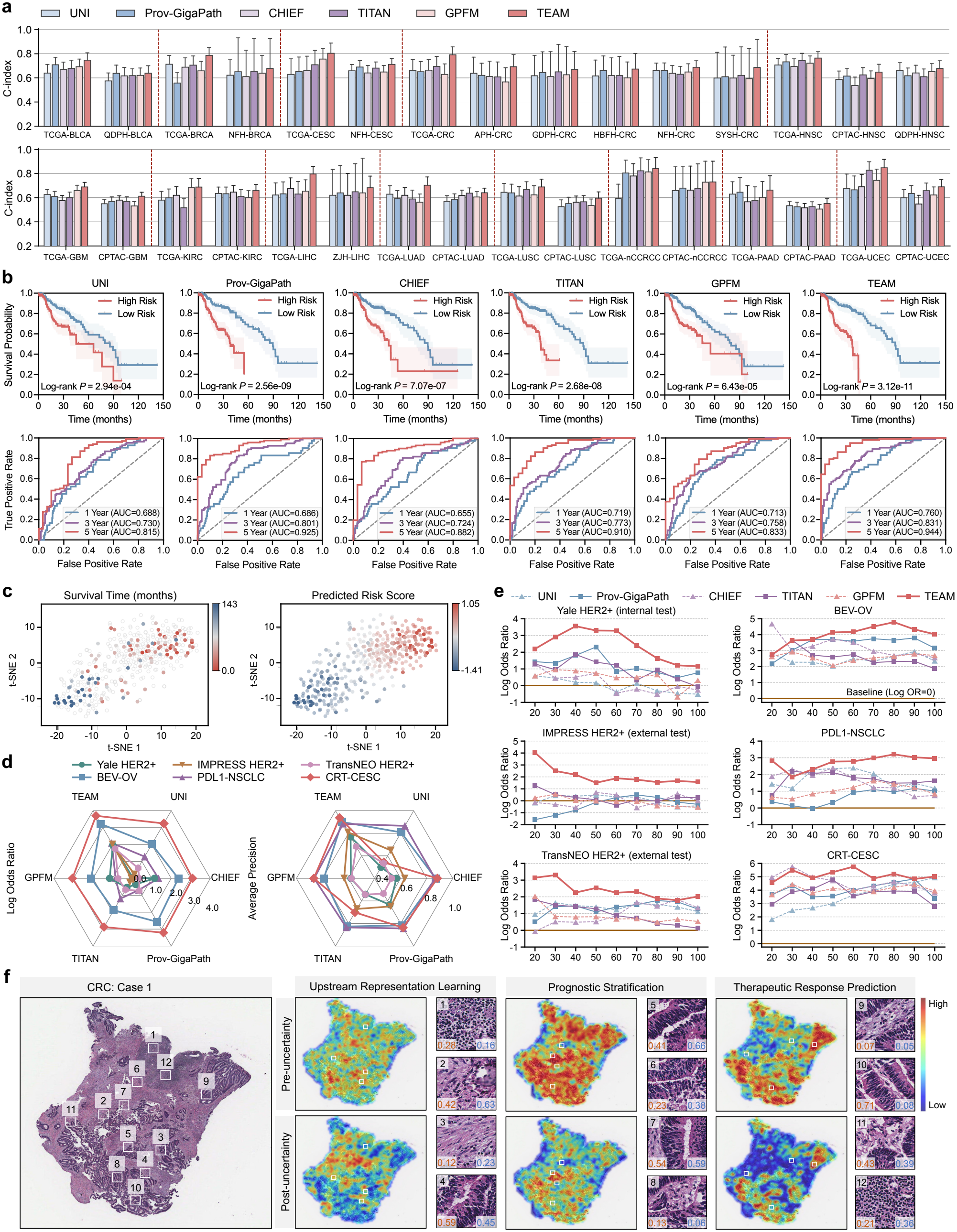
Prognostic stratification and therapeutic response prediction. **a**, C-index for slide-level prognostic stratification in 31 cohorts across 13 cancer types. Error bars indicate 95% confidence intervals (CIs). **b**, Prognostic analysis for the full NFH-CESC cohort. Top: Kaplan–Meier curves comparing low- and high-risk strata produced by each model (shaded area, 95% CI; *P* values from two-sided log-rank tests). Bottom: time-dependent ROC curves at 1, 3, and 5 years. **c**, t-SNE projection of NFH-CESC slide-level embeddings, colored by observed OS time in months (left; censored cases as open light-gray circles) and TEAM-predicted risk score (right; censored cases at reduced opacity). **d**, Therapeutic response prediction performance across six cohorts in four treatment settings, evaluated by log odds ratio (Log OR) and average precision (AP). **e**, Log OR–coverage curves for the six cohorts. **f**, Whole-slide attention heatmaps across three tasks. Four representative patches per task are annotated with aleatoric (orange) and epistemic (blue) uncertainty values.

**Fig. 3.**
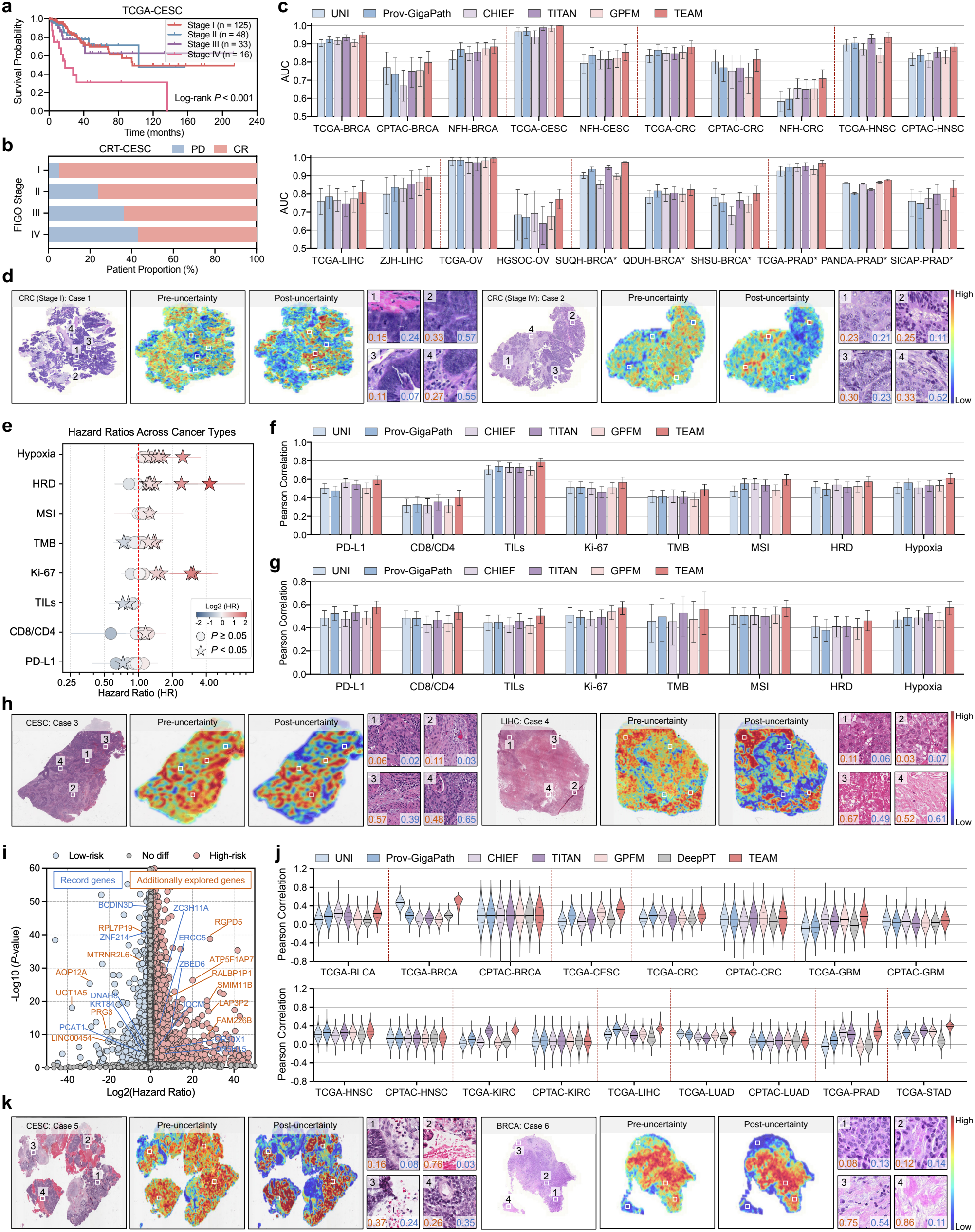
Multi-scale biomarker profiling across cancer staging/grading, TME characterization, and gene expression. **a**, Kaplan–Meier curves for CESC stratified by clinical stage (I–IV). **b**, 100% stacked bars showing CESC treatment outcomes by stage. **c**, AUCs with 95% CIs for cancer staging/grading across internal and external cohorts; within each cancer type, the leftmost cohort is the internal test set; * indicates grading tasks. **d**, Representative slides for staging/grading, with prediction heatmaps before and after uncertainty refinement and patch-level aleatoric (orange) and epistemic (blue) uncertainty. **e**, Forest plot of hazard ratios (HRs) with 95% CIs for eight TME biomarkers. **f**,**g**, PCC between model-predicted and measured biomarker values across six internal TCGA cohorts (f) and three external CPTAC cohorts (g). **h**, Representative slides for TME biomarkers. **i**, Volcano plot of gene-level associations with overall survival across 33 TCGA cancer types (Cox regression; FDR-adjusted *P* values). **j**, Gene-wise PCCs between predicted and measured expression for the seven models under the same regression head. **k**, Representative slides for gene expression.

#### Therapeutic response prediction

We evaluated TEAM for therapeutic response prediction in four treatment settings (Supplementary Table 13; Methods). For neoadjuvant therapy in HER2+ breast cancer (pathological complete response), the downstream model was developed and internally evaluated on Yale HER2+ [26], then externally evaluated on IMPRESS HER2+ [27] and TransNEO HER2+ [28]; bevacizumab treatment in ovarian cancer (BEV-OV) [29], PD-L1 blockade in non-small cell lung cancer (PDL1-NSCLC) [30], and chemoradiotherapy in cervical cancer (CRT-CESC) [31] were evaluated in exploratory within-cohort analyses. TEAM ranks first among the six models in the four settings (Fig. 2d), with a mean odds ratio (OR) of 7.19 on the internal Yale HER2+ test set, compared with 1.06–3.16 for the five baselines. This advantage is retained in the two external cohorts, with mean ORs of 7.52 on IMPRESS HER2+ and 5.74 on TransNEO HER2+, compared with 1.04–2.08 for the baselines (Supplementary Tables 57–59). TEAM’s OR in the exploratory cohorts is 1.6-to 2.1-fold that of the best-performing baseline (Supplementary Tables 60–62). Log OR–coverage curves (Fig. 2e) show that TEAM achieves competitive performance against the baselines across coverage thresholds.

#### Spatial attention patterns

Whole-slide attention heatmaps exhibit task-dependent spatial distributions (Fig. 2f). During upstream learning, attention is broadly distributed across densely nucleated (patch 1), glandular (patch 4), and fibrous regions (patches 2, 3). For prognostic stratification, attention extends to glandular regions (patches 5–7); for therapeutic response, attention concentrates on glandular and densely nucleated regions (patches 9, 12) with reduced weighting in peripheral regions (patches 10, 11). Uncertainty estimation refines these patterns by down-weighting high-uncertainty regions (patches 2, 5, 7, 10, 11) while preserving lower-uncertainty regions (patches 1, 8, 9, 12), suggesting interpretable spatial prioritization. Extended Data Fig. 2 provides additional examples of task-specific attention patterns and uncertainty-based refinement.

### Precise multi-scale biomarker profiling via pan-cancer analysis

#### Cancer staging and grading

Cancer stage and grade directly inform prognostic stratification and treatment planning [32], and higher stage is associated with worse outcomes (Fig. 3a,b). We evaluated TEAM on 20 cohorts encompassing seven cancer types (Supplementary Tables 14–15). Against five baseline models, TEAM achieves the best performance across the 20 cohorts (*P*<0.001), with a mean AUC of 0.873 ± 0.078 (Fig. 3c; Supplementary Tables 65–84). Cross-institutional evaluation further supports robust generalization. For example, TEAM maintains AUCs of 0.935 on TCGA and 0.883 on CPTAC for HNSC staging, with grading AUCs exceeding 0.95 internally and 0.80 externally. The t-SNE visualization (Extended Data Fig. 3a) reveals that samples are organized into different clusters corresponding to stages or grades, suggesting that the model captures clinically meaningful morphological differences. Attention heatmaps for stage prediction (Fig. 3d) show that at an early stage (stage I), attention localizes to glandular structures (patch 3) and regions bordering stroma (patch 1), while at an advanced stage (stage IV), attention concentrates on densely cellular clusters (patches 1–2) and regions with spindle-shaped cells (patch 4). Slides at stages II and III are shown with uncertainty overlays in Extended Data Fig. 3b.

#### TME characterization

TME composition shapes tumor progression and treatment sensitivity [33]. We targeted eight clinically validated biomarkers, whose hazard ratios across nine cancer types show patterns consistent with established biology (Fig. 3e): an immune surveillance profile comprising programmed death-ligand 1 (PD-L1), CD8+/CD4+ T-cell ratio (CD8/CD4), tumor-infiltrating lymphocyte density (TILs), and Ki-67 proliferation index (Ki-67) [2, 34]; and a microenvironmental stress profile encompassing tumor mutational burden (TMB), microsatellite instability (MSI), homologous recombination deficiency (HRD), and hypoxia metagene (Hypoxia) [2, 35]. We evaluated prediction performance for the eight biomarkers on six internal TCGA cohorts (TCGA-BLCA, TCGA-BRCA, TCGA-CESC, TCGA-CRC, TCGA-HNSC, and TCGA-LIHC) and three external CPTAC cohorts (CPTAC-BRCA, CPTAC-CRC, and CPTAC-HNSC), spanning six cancer types (Fig. 3f,g and Supplementary Tables 16–18). Across both internal and external cohorts, TEAM outperforms the competing models (*P*<0.01; Supplementary Tables 85–100), with improvements of +0.063 over the second-best method internally (0.577 vs 0.514 for CHIEF) and +0.055 externally (0.544 vs 0.489 for TITAN). TEAM also preserves inter-biomarker relationships consistent with known TME biology, with clustering on predicted biomarkers yielding prognostically distinct subgroups (Extended Data Fig. 3c–e). Attention heatmaps for TME biomarker prediction (Fig. 3h; Extended Data Fig. 3f) show that attention localizes to cellular regions with surrounding small round nuclei (CESC, patch 1) or intervening fibrous tissue (CESC, patch 2) and to sheets of large polygonal cells (LIHC, patches 1–2). Fibrous or sparsely nucleated regions (CESC, patch 3; LIHC, patches 3–4) have higher data and model uncertainty (0.39–0.67) than patches 1–2 in both examples (≤0.11) and therefore receive stronger uncertainty-based down-weighting.

#### Gene expression

Gene expression profiling reveals molecular programs governing cancer progression and therapeutic response [36], with prognostic relevance observed across cancer types [37] (Fig. 3i). TEAM’s gene expression prediction was evaluated across 17 cohorts (11 internal and 6 external; Supplementary Table 19). We benchmarked TEAM against the five pathology foundation models and DeepPT, a dedicated transcriptome-prediction model, using the same regression head and target genes. Across all genes, TEAM achieves a mean Pearson correlation coefficient (PCC) of 0.237 (pooled over all gene–cohort pairs), compared with 0.104–0.149 for the six baseline models (Fig. 3j; Supplementary Tables 101–118). The mean cohort-level paired PCC differences between TEAM and each baseline range from 0.085 to 0.130 (all *P*≤3.1 × 10^−5^, two-sided Wilcoxon signed-rank tests). For the top 1,000 prognostically associated genes (Methods), the mean PCC of TEAM is 0.247, compared with 0.114–0.150 for the six baseline models (Supplementary Fig. 5a). This advantage persists in sample-wise analyses of both gene sets (Supplementary Fig. 5b,c). When genes are stratified by phenotype category, TEAM achieves the highest median gene-wise PCC across the evaluated subgroups (Extended Data Fig. 3g). Attention heatmaps for gene expression predictions (Fig. 3k; Extended Data Fig. 3h) prioritize densely cellular regions over fibrous areas in the BRCA example (patches 1–2 versus patches 3–4). The fibrous patches show high data uncertainty (0.75 and 0.86) and are down-weighted more strongly by uncertainty estimation than the cellular patches, whose two uncertainty values are both ≤0.14.

### Improving biomarker-driven outcome prediction

TEAM uses multi-scale biomarkers (cancer stage/grade, TME, and gene expression) as complementary sources to improve prognostic stratification and therapeutic response prediction (Fig. 4a). Across 37 cohorts (31 prognostic and 6 therapeutic; Supplementary Tables 26–64), performance improves progressively with biomarker integration: for prognostic stratification, single-biomarker models achieve C-indices of 0.738–0.751, dual-biomarker models 0.767–0.770, and the fully integrated model (+Stage/Grade+TME+Gene) achieves the highest C-index of 0.783 (ΔC-index: +0.081 vs the pathology-only model of 0.701, *P*<0.001; Fig. 4b), improving the C-index by 0.114 (95% CI, 0.100–0.127) over the best-performing baseline in each cohort (Extended Data Fig. 9a); for therapeutic response prediction, the fully integrated model achieves a mean OR of 46.3, compared with 30.6–31.1 for single-biomarker and 32.0–35.9 for dual-biomarker models (Fig. 4c). Shapley values [38] (Fig. 4d) identify TME as the largest contributor to prognostic stratification and stage/grade as the largest contributor to therapeutic response prediction, with substantial heterogeneity across cancer types and treatment settings. Pathology nevertheless remains the largest single source of attention in both tasks, retaining 59–62% of the normalized weight in prognostic stratification and 42–54% in therapeutic response prediction (Fig. 4g,h). Within-stratum analyses suggest that TEAM captures additional prognostic information beyond individual biomarkers, enabling significant risk stratification within stage-homogeneous and TME-homogeneous subsets (all *P*<0.05; Extended Data Fig. 4a,b). In the 23 prognostic cohorts with recorded tumor stage, the TEAM-derived risk score remains prognostic after adjustment for stage (adjusted hazard ratios per standard deviation, 1.23–7.74; Supplementary Fig. 2). Gene set enrichment analysis (GSEA) [39] reveals that high-risk scores are associated with invasion-related pathways, while low-risk scores are associated with metabolic programs (Extended Data Fig. 4c–f).

**Fig. 4.**
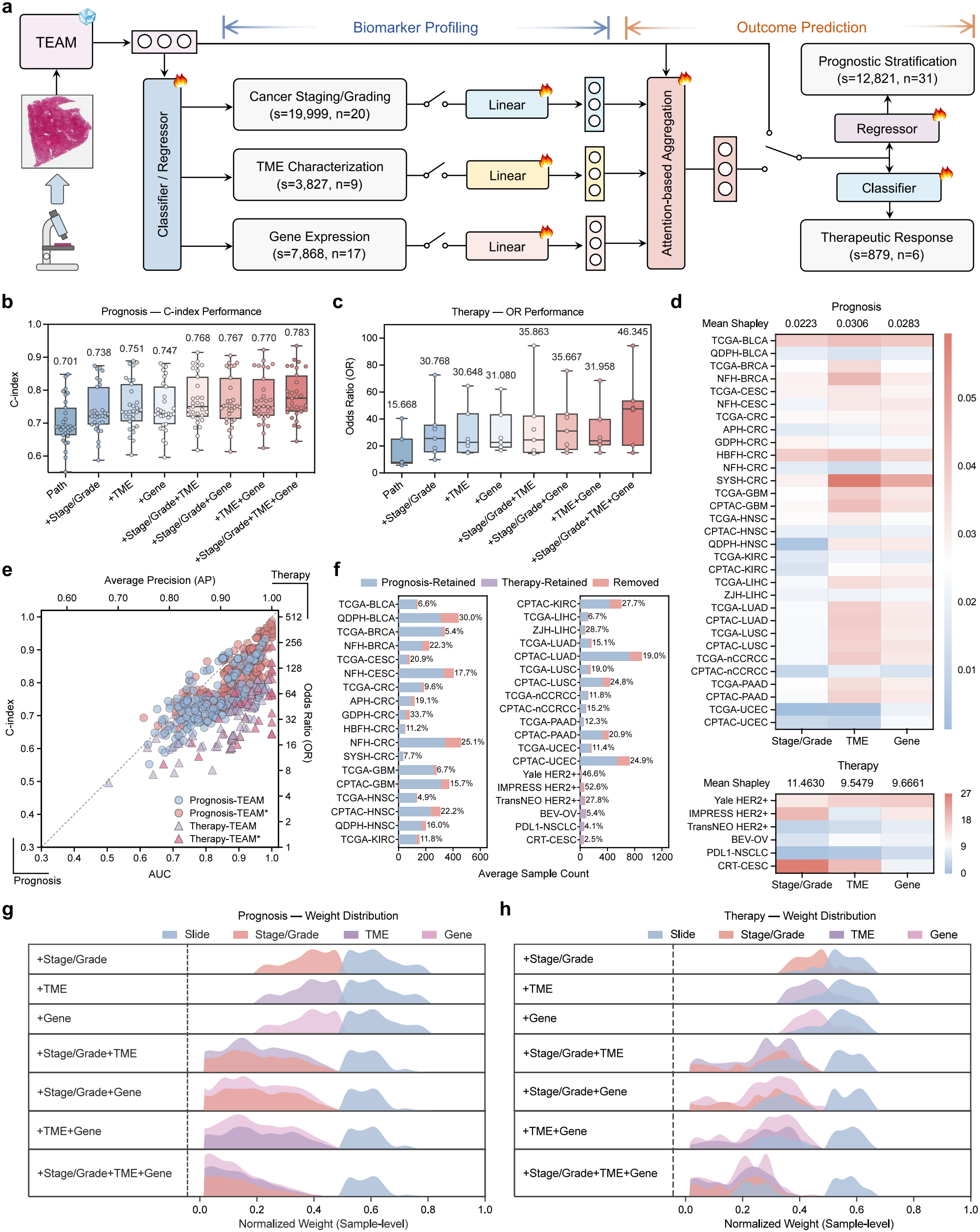
Biomarker-driven outcome prediction. Eight model variants were evaluated: pathology-only (Path), Path plus staging/grading (+Stage/Grade), TME (+TME), or gene expression (+Gene), and their combinations. **a**, Analytical pipeline for biomarker profiling and outcome prediction (s, samples; n, cohorts). **b**,**c**, C-index across 31 prognostic cohorts (b) and OR across 6 therapeutic response cohorts (c). Box plots show median and interquartile range (IQR); whiskers extend to 1.5 *×* IQR. Numbers above boxes indicate the mean across cohorts. **d**, Shapley values for prognosis (top) and therapeutic response (bottom). Values represent average marginal performance gains (ΔC-index for prognosis, ΔOR for therapeutic response). **e**, Performance of TEAM on all cases (TEAM) and on the cases retained by uncertainty-based case selection (TEAM*) across prognostic (C-index and AUC) and therapeutic (OR and AP) tasks; each point represents one cohort under one model configuration. **f**, Sample retention after uncertainty-based case selection (TEAM*) for prognostic and therapeutic tasks. **g**,**h**, Distributions of normalized attention weights for prognosis (g) and therapeutic response (h).

To translate uncertainty estimates into a practical indicator for clinical use [19], we introduced uncertainty-based case selection (TEAM*), which retains predictions with final uncertainty 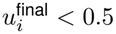 (Methods). For pathology-only prognostic stratification, TEAM* retains a mean of 74.0% ± 10.3% of cases (coverage) and reaches a mean C-index of 0.880 on the retained cases, compared with 0.701 on all cases, reflecting a trade-off between coverage and performance. In the evaluation on the cases retained by TEAM*, the five baselines also improve (mean C-index, 0.692–0.714) but still underperform TEAM* (all *P*<0.001, two-sided Wilcoxon signed-rank tests; Extended Data Fig. 10a and Supplementary Table 119), whereas the baselines do not substantially benefit from excluding their own most unstable cases at the same coverage (0.611–0.632, compared with 0.622–0.645 on all cases; Methods). Consistent results are observed in the other downstream tasks (Extended Data Fig. 10b–e and Supplementary Table 119). Across all biomarker configurations, TEAM* retains 82.1% of cases on average (range 47.4–97.5%; Fig. 4f), with performance on the retained cases exceeding that on all cases by 0.077 in C-index and 0.068 in AUC for prognostic stratification and by 22.2 in OR and 0.046 in AP for therapeutic response prediction (Fig. 4e and Supplementary Table 120).

### Agentic orchestration for facilitating outcome-related clinical reasoning

To enable clinical reasoning beyond static prediction, we integrated TEAM’s encoder with three LLMs to build TEAM-Llava (based on Llava [40, 41]), TEAM-DeepSeek (based on DeepSeek-R1 [42]), and TEAM-Agent (based on GPT-4o [43]), as shown in Fig. 5a. TEAM-Agent implements a supervisor architecture that coordinates biomarker profiling with outcome prediction through a six-step workflow (Fig. 5b). The complete workflow is shown in Extended Data Fig. 5b, with prompt specifications provided in the Prompt Sets of the Supplementary Information. We evaluated these models on standardized closed-ended benchmarks and pathologist-assessed open-ended scenarios (Supplementary Table 21).

**Fig. 5.**
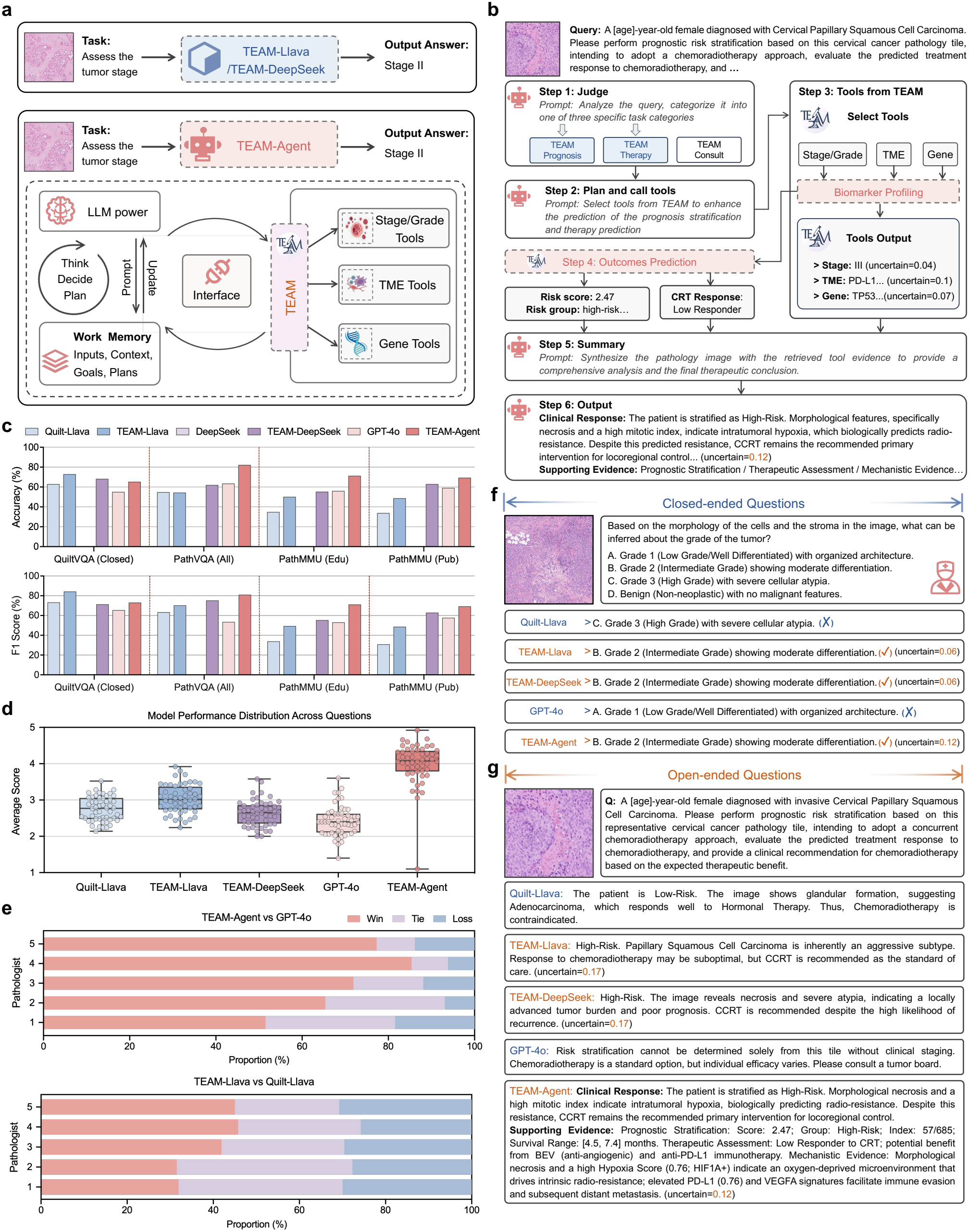
Agentic orchestration and conversational clinical reasoning. **a**, Architectural comparison between traditional multimodal large language models (MLLMs) and TEAM-Agent. **b**, TEAM-Agent workflow for patient-specific prognostic queries. **c**, Accuracy (top) and F1-score (bottom) on four closed-ended benchmarks (Supplementary Tables 123–126). DeepSeek-R1 accepts text input only and was not evaluated on these image-based benchmarks. **d**, Distribution of clinician rankings for 260 questions evaluated by five pathologists (1–5 scale; 5 = best). Rankings aggregated using Borda count. **e**, Win, tie, and loss proportions for each pathologist in head-to-head comparisons of TEAM-Agent vs GPT-4o (top) and TEAM-Llava vs Quilt-Llava (bottom). **f**, Closed-ended example: tumor grade classification integrating architectural and stromal patterns. Checkmarks indicate correct answers. **g**, Open-ended example: comprehensive prognostic assessment and treatment recommendations.

Closed-ended evaluation spanned 4,270 questions across QuiltVQA [41] (*n* = 343), PathVQA [44] (*n* = 3,391), and PathMMU [45] (EduContent: *n* = 255, PubMed: *n* = 281). TEAM-Agent achieves the highest performance (Fig. 5c; Supplementary Tables 123–126), with an accuracy of 72.1% and an F1-score of 73.7%, outperforming GPT-4o (accuracy 58.4%, F1-score 57.5%). TEAM-Llava outperforms Quilt-Llava, and TEAM-DeepSeek enables text-only DeepSeek-R1 to achieve performance comparable to multimodal baselines. Representative examples (Fig. 5f; Extended Data Fig. 5c,d) illustrate differences in reasoning quality: TEAM-Agent correctly identifies intermediate differentiation in a cancer grading task by integrating architectural organization with stromal characteristics, whereas GPT-4o and Quilt-Llava misclassify the case. Built on the TEAM encoder, all three TEAM-based LLMs inherit its uncertainty estimation and report a prediction uncertainty with each response.

For open-ended evaluation, five pathologists blindly ranked model responses for 260 PathQABench questions [46] on a 1–5 scale (5 = best; Fig. 5d). TEAM-Agent achieves the highest mean score (4.00 ± 1.25), compared with 2.42 ± 1.30 for GPT-4o and 2.75 ± 1.13 for Quilt-Llava, with a pairwise win rate of 86.0% against GPT-4o among non-tied evaluations (*P*<0.001; effect size *r* = 0.74; Extended Data Fig. 5a and Supplementary Tables 127–128). TEAM-Llava similarly outperforms Quilt-Llava (57.7% wins among non-tied evaluations; *P*<0.001; Fig. 5e and Supplementary Table 128). Representative outcome queries (Fig. 5g; Extended Data Fig. 5e,f) show that TEAM-Agent generates structured outputs comprising risk stratification with survival estimates, therapeutic response assessment, mechanistic evidence derived from predicted biomarkers, and aggregated prediction uncertainty.

### Clinician-in-the-loop refinement for improving prediction reliability

We developed an online interactive interface that enables pathologists to refine model predictions by adjusting region-specific attention weights on the generated heatmap via freehand scribbles or bounding boxes (Fig. 6a,b; Extended Data Fig. 6a–f; Supplementary Movie 1). Each annotation reweights patch contributions in incremental steps from −0.5 to +0.5 and updates slide-level predictions through uncertainty-weighted aggregation, with an integrated LLM module providing stepwise interpretation and a final synthesized report. We evaluated this interactive capability across three cohorts (TCGA-CESC, CPTAC-KIRC, and BEV-OV), comprising 40 cases per cohort (*n* = 120 total), each refined independently by five pathologists. Pathologist-guided refinement improves prediction performance for the refined cases across the downstream tasks (*P*<0.001; Fig. 6c and Supplementary Table 129). Mean C-index gains for prognostic stratification are +0.053 to +0.065, and therapeutic response prediction shows a 2.29-fold improvement in OR. These improvements are consistent across the five pathologists, with a median intraclass correlation coefficient (ICC) of 0.94 [47]. Refinements are typically completed within two interactions (median = 2 across all cohorts), with 94% of cases requiring ≤3 interactions (Fig. 6d and Supplementary Table 130). A representative case (Fig. 6e) illustrates this interactive process: as the pathologist progressively adjusts the model’s attention in selected regions, the estimated survival range increases from 13.8–27.4 months to 90.1–103.2 months (observed OS, 96.3 months), with the predicted stage changing from II to I and the estimated uncertainty decreasing from 0.37 to 0.16. Additional refinement examples for therapeutic response prediction are provided in Extended Data Fig. 6g,h.

**Fig. 6.**
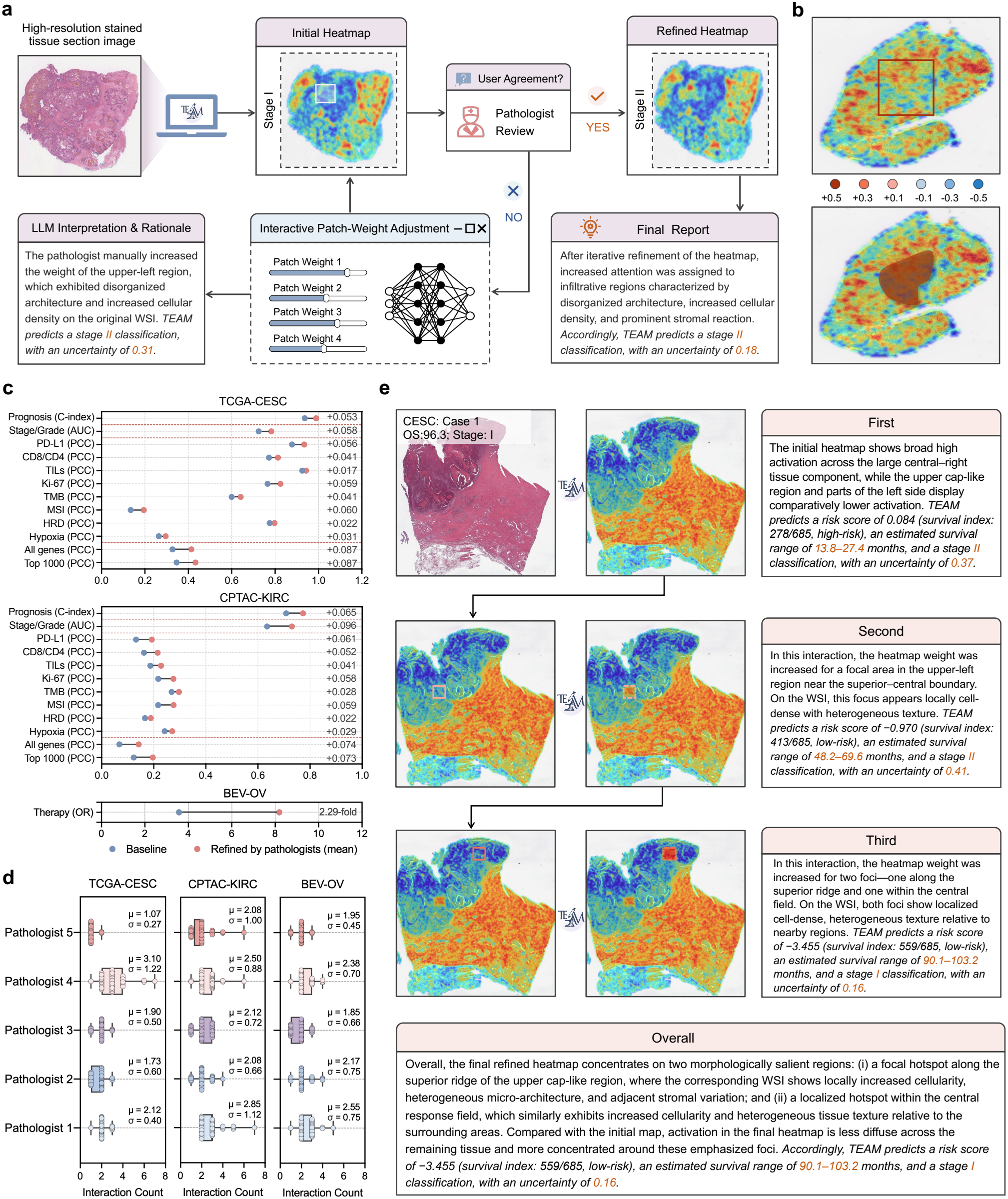
Clinician-in-the-loop interactive prediction refinement. **a**, Interactive workflow enabling pathologists to iteratively refine model predictions through attention heatmap annotation. Pathologists review initial predictions and either accept or adjust patch weights via an interactive interface; an integrated LLM module provides stepwise interpretation and rationale, culminating in a final report. **b**, Two annotation modalities (bounding box and freehand scribble) with graded weight adjustments ranging from *−*0.5 to +0.5 for decreasing or increasing patch contributions. **c**, Performance before and after pathologist-guided refinement in three cohorts (TCGA-CESC, CPTAC-KIRC, and BEV-OV; *n* = 40 cases per cohort), with each case refined independently by five pathologists and the refined performance averaged across pathologists. Metrics are the C-index for prognostic stratification, AUC for cancer staging/grading, PCC for TME characterization (PD-L1, CD8/CD4, TILs, Ki-67, TMB, MSI, HRD, and Hypoxia) and gene expression prediction (all genes and the top 1,000 prognostically associated genes), and OR for therapeutic response prediction. Numbers at right, change after refinement (fold change for OR). **d**, Distribution of interaction counts per pathologist across cohorts, with mean (*µ*) and standard deviation (*σ*) annotated. **e**, Representative refinement case illustrating progressive attention adjustment from the initial prediction through two rounds of refinement, with corresponding risk scores, survival estimates, staging/grading predictions, and uncertainty values at each step. Uncertainty values and ground-truth labels are shown for illustration and were not displayed to the pathologists during refinement.

## Discussion

In this study, we present TEAM, an interactive trustworthy AI pathology copilot for biomarker-driven outcome prediction. Unlike general-purpose pathology foundation models such as TITAN [14] and GPFM [15], TEAM integrates risk-aware pretraining with dual uncertainty quantification, uses profiled biomarkers as intermediate features to contextualize prognostic and therapeutic estimates, and embeds these capabilities within an agentic architecture that supports clinical reasoning and interactive prediction refinement. Across 51 multi-institutional cohorts and 88 benchmarks, TEAM outperforms existing models (Supplementary Table 121), and this integrated design improves not only predictive accuracy, but also the biological traceability and clinician oversight of pathology-based outcome assessment.

TEAM’s biomarker-driven design grounds outcome prediction in the multi-scale biological processes that shape cancer prognosis and therapeutic response. By using cancer stage/grade, TME characterization, and gene expression as intermediate features for prognostic and therapeutic prediction, TEAM integrates biomarker profiling with outcome prediction rather than treating them as parallel downstream tasks. Multi-factor fusion analyses show progressive performance gains as more biomarkers are incorporated. Shapley value analysis [38] and attention weight decomposition further reveal substantial heterogeneity in biomarker contributions across cancer types, linking outcome predictions to context-specific biomarker configurations and supporting both the biological plausibility and clinical interpretability of TEAM. Together, these findings provide a traceable link between TEAM’s outcome predictions and their underlying biological correlates, helping shift computational pathology from purely pattern-based prediction toward biologically informed outcome assessment.

Beyond biomarker integration, risk-aware pretraining extends morphology-centered pathology representation learning by aligning TEAM’s representations with the relative risk prior across cancer types. These riskaware representations support consistent performance across biomarker profiling, prognostic stratification, and therapeutic response prediction, suggesting that alignment with the relative risk prior helps capture morphological patterns associated with clinical risk. Complementing these representations, dual uncertainty quantification incorporates data (aleatoric) and model (epistemic) uncertainty into TEAM’s downstream predictions, an approach rarely integrated into computational pathology workflows [19, 48]. Restricting predictions to low-uncertainty cases improves performance on the retained cases and preserves TEAM’s advantage over the baselines at matched coverage. These uncertainty estimates also provide a practical indicator for clinical use, helping direct clinician attention toward cases warranting additional scrutiny and strengthening trustworthiness in AI-assisted pathology.

TEAM also functions as an interactive pathology copilot [46] by combining agentic reasoning with clinician-in-the-loop refinement. TEAM-Agent integrates the biomarker-driven outcome prediction pipeline into an agentic architecture [49] that grounds clinical reasoning in the model’s quantified outputs (i.e., risk estimates, biomarker profiles, and prediction uncertainty) rather than unconstrained language generation. In evaluations spanning standardized benchmarks and blinded pathologist assessment, this agentic architecture outperforms all compared models. The clinician-in-the-loop interface further moves beyond conventional accept-or-reject decisions [20] by enabling pathologists to iteratively refine predictions through region-specific attention adjustment. Such refinement improves prediction performance across evaluation tasks, with prediction uncertainty changing accordingly. TEAM thus points to a practical paradigm for collaborative decision-making in computational pathology, in which agentic reasoning is grounded in quantified outputs and clinical expertise is channeled into iterative prediction refinement.

Our study has several limitations that warrant consideration. Prospective validation in routine clinical workflows is needed to determine whether TEAM can meaningfully support prognostic assessment and treatment recommendation. Therapeutic response evaluation was limited to four treatment settings, and further validation across additional targeted, immune-based, and combination therapies is needed to reflect the diverse landscape of precision oncology. TEAM-Agent grounds clinical reasoning in quantified outputs, but the narrative responses should remain subject to expert clinical review before influencing patient management. TEAM identifies risk-relevant morphological features that inform biomarker profiling and outcome prediction, but the biological mechanisms underlying these associations remain to be fully explored. Future work integrating multimodal data such as medical imaging and genomics may help elucidate the underlying mechanisms from both macroscopic and molecular perspectives. The modular design allows incorporation of newly validated biomarkers, enabling the framework to evolve with clinical advances toward more comprehensive AI-assisted decision support in precision oncology.

## Methods

### Pretraining dataset curation and preprocessing

We assembled a pretraining corpus from 65 publicly available datasets across 23 anatomical sites and diverse tissue types, including large research consortia such as TCGA [50], GTEx [51], and PAIP [52]. The corpus comprised 55,648 WSIs and 1,750,648 ROIs, collectively yielding 360,786,410 tissue patches (Supplementary Table 1). For TCGA cohorts, samples were stratified by cancer type, with approximately 70% of patients allocated to pretraining and the remaining 30% reserved for downstream evaluation. Patient-level splits were applied before model development to prevent leakage, and pretraining was performed in a task-agnostic manner without individual-level outcome annotations.

All WSIs underwent standardized tissue segmentation and patch extraction. Tissue regions were segmented using threshold-based methods followed by morphological refinement. Slides with unreadable or unsupported formats were excluded, and low-quality regions were further removed by contour filtering (connected components *<*100 pixels or regions with excessive internal holes). The remaining tissue regions were tessellated into non-overlapping 512 × 512 pixel patches at native magnification (typically 20× or 40×), and ROI-only datasets were processed using the same patch size. To preserve morphology across imaging protocols, patches were extracted without magnification normalization and resized to 224 × 224 pixels for encoder compatibility.

### Risk-aware feature encoder with dual uncertainty

For a cohort of *N* WSIs, the *i*-th slide is tessellated into *M*_*i*_ tissue patches, forming the set 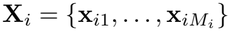. We adopted a DINOv2 vision transformer [23] initialized with UNI weights [11] as the patch encoder (denoted TEAM-V). During pretraining, we retained self-distillation and masked image modeling objectives while selectively updating the first and last two transformer blocks (detailed in Supplementary Tables 2–3). The encoder maps each patch **x**_*im*_ to a *d*-dimensional feature representation **z**_*im*_ ∈ ℝ^*d*^, where 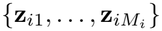 denotes the patch-level features for the *i*-th slide. These features are subsequently processed to quantify data (aleatoric) and model (epistemic) uncertainty.

#### Aleatoric uncertainty modeling

We modeled aleatoric uncertainty using a variational autoencoder (VAE) that maps patch features to a Gaussian latent distribution *q*_*θ*_(**z** | **x**) = *N* (***µ***_*θ*_(**x**), 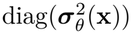, where the variance 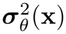 quantifies input-dependent uncertainty. The VAE was trained by maximizing the evidence lower bound (ELBO):

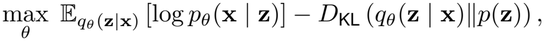

where *p*(**z**) = *N* (**0, I**) is the standard Gaussian prior and *D*_KL_ is the Kullback–Leibler divergence. The predicted variance 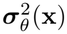 was normalized to [0, 1] and averaged to obtain the aleatoric uncertainty score *u*^ale^ ∈ [0, 1] for downstream aggregation.

#### Epistemic uncertainty modeling

We modeled epistemic uncertainty using Monte Carlo dropout, a Bayesian approximation that quantifies model uncertainty through stochastic forward passes [21]. For each patch **x**, we performed *T* forward passes with dropout masks **M**_*t*_ (dropout rate *π* = 0.3) applied to the encoder output features, producing outputs 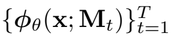. The predictive mean and variance are:

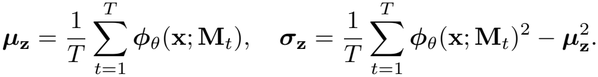

We normalized the variance element-wise by its maximum 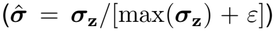 and averaged across dimensions to obtain the epistemic uncertainty score 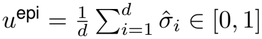 for patch-level weighting in slide aggregation. The number of forward passes was set to *T* = 3 as a computationally efficient default. For sensitivity analysis, we evaluated *T* = 5 and *T* = 20 with all other settings unchanged across the 31 prognostic cohorts (C-index) and the 6 therapeutic response cohorts (Log OR and AP; Extended Data Fig. 9c–e and Supplementary Table 122).

### Clinical metadata encoder

The Clinical Metadata Encoder (denoted TEAM-R) represents four demographic variables (i.e., age, sex, race, and ethnicity; Supplementary Table 6) as free-text descriptions, accommodating the missing or incomplete records typical of real-world clinical settings. TEAM-R was used only during pretraining, with no clinical metadata provided during downstream biomarker profiling or outcome prediction. Missing variables were omitted from the free-text input without imputation, and the text “None” was used when all four variables were missing. For the *i*-th slide, the clinical description *c*_*i*_ of the corresponding patient was encoded using frozen BioBERT [24], which processes the entire text sequence and outputs a *d*-dimensional embedding **v**_*i*_ ∈ R^*d*^ extracted from the [CLS] token:

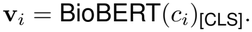

The embedding **v**_*i*_ is then aggregated with patch features in the self-supervision head. Availability of the four variables by pretraining dataset is given in Supplementary Table 7.

### Self-supervision head with relative risk prior

#### Uncertainty-weighted aggregation

Patch features 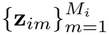 and clinical embedding **v**_*i*_ were aggregated using attention weights computed by:

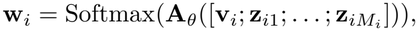

where **A**_*θ*_ is a learnable linear projection and [·] denotes concatenation. This yields 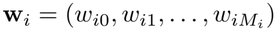, with *w*_*i*0_ for clinical features and 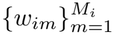 for patches. Patch weights were modulated by their uncertainty estimates (aleatoric 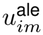 and epistemic 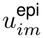, both in [0, 1]):

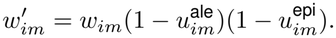

The slide-level representation was computed as the uncertainty-weighted average:

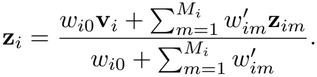

The final risk score *r*_*i*_ = *ψ*_*θ*_(**z**_*i*_) ∈ R was obtained through a multilayer perceptron (MLP), enabling pan-cancer risk ranking.

To propagate uncertainty alongside the slide representation, the two patch-level uncertainty components were combined as

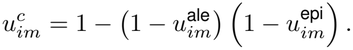

The slide-level pathology uncertainty was obtained by aggregating these patch-level estimates using the normalized attention weights:

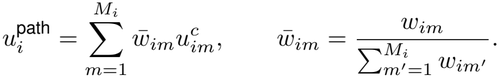

Here, *w*_*im*_ is the attention weight of the *m*-th patch before uncertainty modulation, and *M*_*i*_ is the number of patches in the *i*-th slide. For direct prediction tasks based on the slide representation (biomarker profiling and pathology-only outcome prediction), the final uncertainty was 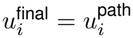.

#### Relaxed ranking function

To leverage population-level survival statistics while accounting for patient hetero-geneity, we developed a pairwise ranking function with registry-specific survival priors. For a sample pair (*p, q*), supervision was defined when their survival gap exceeded a clinical margin. If the *q*-th sample had substantially worse prognosis than the *p*-th sample based on population statistics, we enforced *r*_*q*_ *> r*_*p*_, where *r* denotes the model-predicted risk score. Formally, we defined the binary supervision signal as:

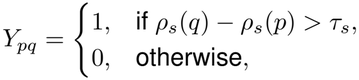

where *ρ*_*s*_(·) denotes the relative risk prior for a given cancer type in registry *s*, and *τ*_*s*_ is the clinical margin threshold. We adopted the thresholding rank-loss framework [53] with hinge relaxation and used a fixed hinge margin of 2 without further tuning. Following this framework, the loss reduces to a pairwise margin objective with the positive-part operator |*x*|_+_ = max(*x*, 0):

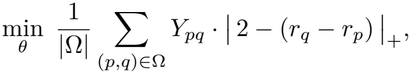

where Ω denotes the set of sampled within-registry training pairs (implemented via mini-batch pair sampling). In practice, population-level priors may not hold uniformly at the individual level, and clinical outcomes can be noisy. We therefore introduced an adaptive pairwise weight ϒ_*pq*_ ∈ [0, 1] denoting the strength of the prior ordering between the *p*-th and *q*-th samples, and penalized deviations from the prior with a regularization weight *δ >* 0:

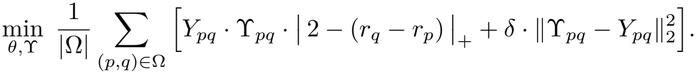

We further adapted supervision to the feature context by parameterizing ϒ_*pq*_ as a learnable function ϒ_*θ*_(**z**^*p*^, **z**^*q*^ ) ∈ [0, 1] that modulates supervision strength based on feature similarity, yielding the final objective:

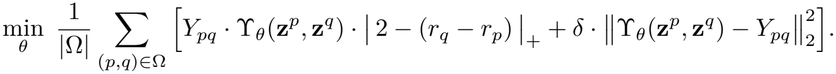

This formulation integrates registry-specific survival priors into optimization while allowing pairwise weights to adapt to latent feature similarity, thereby improving robustness to label noise and inter-patient heterogeneity.

#### Relative risk prior derivation

The relative risk prior *ρ*_*s*_(·) was derived from 5-year cancer-specific survival statistics from three population-based cancer registries: the National Central Cancer Registry of China [54], the EUROCARE-5 database [55], and the SEER Statistics Network of the United States (https://seer.cancer.gov/statistics-network/explorer/application.html). Within each registry, survival rates were stratified by ICD-10 site and histological subtype when available, with pairwise comparisons restricted to samples from the same registry to avoid cross-registry confounding. The survival statistics of the three registries come from different registry systems and reporting periods, so the clinical margin *τ*_*s*_ was set separately for each registry. Clinicians participating in this study first provided qualitative prognostic comparisons without being shown the 5-year survival rates. These judgments were then quantified using the survival statistics of each registry, and the clinicians confirmed the resulting margins of approximately 15% for China, 20% for Europe, and 5% for the United States. Supervision signals *Y*_*pq*_ were generated only when the within-registry survival gap exceeded *τ*_*s*_. These survival statistics were used as weak pairwise constraints during training, with registry identifiers excluded from model inputs and inference. The strength of these constraints was modulated through feature-adaptive weights ϒ_*θ*_(**z**^*p*^, **z**^*q*^ ) to preserve individual-level variability. Details of within-registry pairing are provided in Extended Data Fig. 7 and Supplementary Table 8.

### Biomarker-driven outcome prediction

We developed a multi-source fusion framework that integrates slide-level morphological features with three profiled biomarker sources: cancer staging/grading, TME characterization (eight markers), and gene expression (15,717 genes). Instead of operating on raw encoder features, the fusion framework uses profiling outputs from each biomarker source as intermediate representations, so that downstream outcome estimates are conditioned on profiled biomarkers. These profiling outputs (cancer stage/grade, TME characterization, and gene expression) were predicted from the WSI rather than supplied as clinical metadata. The fusion framework therefore required only outcome labels, not ground-truth biomarker labels. All branches share the same patch-level encoder while employing task-specific classifiers across cancer types. A learnable cancer type embedding was introduced to capture tissue-specific morphological patterns. The slide-level pathology representation was projected to a shared representation space, combined with the cancer type embedding, and normalized via layer normalization to yield the source representation 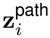 and scalar uncertainty 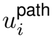. For each biomarker source, profiling outputs were similarly projected to yield a source representation 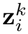 and uncertainty 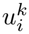. To integrate the pathology and biomarker sources, we employed an attention-based fusion mechanism in which source weights were computed through temperature-scaled attention with z-score normalization:

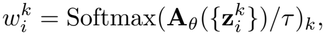

where *k* ∈ {path, stage/grade, TME, gene}, **A**_*θ*_ is a learnable scoring function, and *τ* is the temperature parameter. The same attention weights were then used to fuse both source representations and their uncertainties:

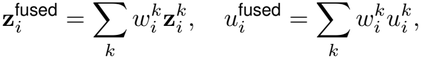

where 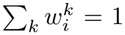 and 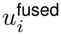 denotes the uncertainty obtained by multi-source fusion. This yields a unified representation for outcome prediction alongside an attention-weighted uncertainty estimate across sources. For biomarker-driven outcome prediction, the final uncertainty was 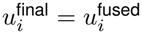.

### Agentic orchestration for clinical reasoning

To enable knowledge-based clinical reasoning, we implemented three TEAM-based language reasoning variants: TEAM-Llava and TEAM-DeepSeek rely on vision-language alignment, whereas TEAM-Agent performs dynamic tool orchestration. For TEAM-Llava, TEAM-V replaced the native Llava vision encoder [40]; for TEAM-DeepSeek, TEAM-V served as the visual encoder paired with DeepSeek-R1 [42]. Alignment training followed a two-stage protocol adapted from Quilt-Llava [41]: the first stage used Quilt-1M image-caption pairs [56] for feature alignment; the second applied Low-Rank Adaptation (LoRA) [57] on Quilt-Instruct question-answer pairs for visual instruction tuning, with TEAM-V kept frozen. Training configurations and datasets are detailed in Supplementary Tables 4 and 20. Uncertainty estimates from TEAM-V are propagated alongside visual features to inform language model reasoning, supporting interpretation and triage under uncertainty. TEAM-Agent implements a GPT-4o-based supervisor architecture [43] that dynamically orchestrates TEAM modules according to patient-specific queries. For each query, TEAM-Agent classifies the task as prognostic stratification (TEAM-Prognosis), therapeutic response prediction (TEAM-Therapy), or clinical consultation (TEAM-Consult), and invokes the relevant biomarker profiling and outcome prediction modules. The results are then synthesized into a structured clinical response covering risk stratification, treatment recommendation, mechanistic rationale, and aggregated uncertainty. Full prompts, tool-selection rules, and output formats are provided in the Prompt Sets of the Supplementary Information. At inference, TEAM-Agent receives the pathology image and the accompanying question from each benchmark, with no clinical metadata provided and without invoking TEAM-R.

### Clinician-in-the-loop refinement

To enable clinician-guided prediction refinement, we implemented an interface that allows manual adjustment of attention weights. Clinicians review WSIs with overlaid attention heatmaps visualizing patch-level weights 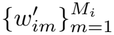 and can modify attention for specific regions through bounding boxes or freehand scribbles. Upon annotation completion, all patch weights are renormalized via softmax:

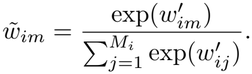

The updated weights 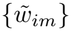 are then used in the standard aggregation pipeline (without the clinical embedding **v**_*i*_, as in all downstream tasks) to construct slide-level representations for generating refined predictions, enabling incorporation of domain expertise in ambiguous cases. Throughout the interactive refinement process, an integrated LLM module (i.e., GPT-4o) provides stepwise interpretation and rationale for each adjustment, synthesizing the refined predictions with morphological evidence into a final report. During each interaction, pathologists review the WSI, attention overlay, and current prediction, with uncertainty estimates shown only after refinement and ground truth used only for post hoc evaluation (Supplementary Fig. 4). After each region selection, the attention weights and prediction for the current case are updated in real time without changing any model parameters. These steps are repeated until the pathologist considers that the model has adequately attended to the relevant pathological regions, without a predefined number of interactions. Refinement is case-specific and does not affect predictions for other patients, so the reported gains describe the refined cases themselves rather than generalization to unannotated cases.

### Baseline models

We benchmarked TEAM against representative external pathology foundation models and visual reasoning baselines, with baseline implementations following published specifications where available. For prognostic stratification, therapeutic response prediction, cancer staging/grading, TME characterization, and gene expression prediction, we evaluated five pathology foundation models spanning distinct backbone designs: UNI [11], a ViT-Large encoder pretrained with DINOv2; Prov-GigaPath [12], a LongNet-based slide encoder; CHIEF [13], a tile-to-slide model combining unsupervised pretraining with weak supervision; TITAN [14], a slide-level visionlanguage foundation model; and GPFM [15], a knowledge-distilled pathology foundation model. We additionally compared against DeepPT [58], a ResNet-50-based slide-level transcriptomic regression model using matched WSI-transcriptome pairs, for gene expression prediction. Each model was implemented as published for the tasks covered by its original publication. In the remaining tasks, slide-level models (Prov-GigaPath, CHIEF, and TITAN) retained their native slide-level representations and only the task-specific prediction head was adapted, whereas patch-level models (UNI and GPFM) used attention-based multiple-instance learning (ABMIL) to obtain slide-level representations before the corresponding prediction head. The pretrained encoders of all five foundation models were kept frozen, and only the task-specific prediction heads and the ABMIL modules (where used) were trained. Task-specific implementations and detailed model settings are provided in Supplementary Tables 23–24. For histopathology visual reasoning, external baselines included Quilt-Llava (histopathology-specific) [41] and GPT-4o, while DeepSeek-R1 served as the language backbone for the TEAM-DeepSeek variant. To quantify the contribution of TEAM-R during pretraining, we evaluated Meta-only and TEAM w/o R. Meta-only used only the four demographic variables provided to TEAM-R, formatted as free-text descriptions and encoded with frozen BioBERT, with missing variables handled as for TEAM-R. The encoded metadata representation was passed to the corresponding task-specific prediction head, with a Cox prediction head used for prognostic stratification. Cohort-level metadata availability is provided in Supplementary Table 25. TEAM w/o R removed TEAM-R entirely during pretraining, with all other experimental settings unchanged.

### Downstream evaluation

All downstream experiments were conducted under a unified evaluation protocol. Feature extraction used the pretrained TEAM-V patch encoder with frozen weights, while uncertainty mapping layers remained trainable for task-specific calibration. Sample sizes and detailed dataset characteristics are provided in Supplementary Tables 10–21, with the 13 in-house validation cohorts (across six cancer types) and institutional details summarized in Supplementary Table 9. Cancer type abbreviations are listed in Supplementary Table 131.

For outcome prediction, overall survival (OS) served as the primary endpoint of prognostic models, with training using Cox proportional hazards loss [59] and assessment by C-index, Kaplan–Meier analysis, and time-dependent ROC curves. The AUC reported for the prognostic cohorts (Supplementary Tables 26–56 and Fig. 4e) is the area under the ROC curve at 36 months, with deaths within 36 months as cases, follow-up beyond 36 months as controls, and observations censored earlier excluded. For within-cohort analyses, the task-specific downstream head was fitted and evaluated using training and test sets from the same cohort. In external evaluation, the downstream head trained on the corresponding internal cohort or cohorts was transferred unchanged to the external cohort, without refitting or hyperparameter tuning on the external data. Therapeutic response was predicted in six cohorts. In HER2+ breast cancer, pathological complete response after neoadjuvant therapy was predicted from pretreatment biopsies, with Yale HER2+ used for model development and internal evaluation and IMPRESS HER2+ and TransNEO HER2+ for external evaluation. BEV-OV, PDL1-NSCLC, and CRT-CESC were evaluated in exploratory within-cohort analyses using 7:3 splits, with CR/PR as responders and SD/PD as non-responders. In all six cohorts, models were optimized using cross-entropy loss and evaluated using AP, the mean OR for responder enrichment across cut-offs classifying the top 20–90% of patients ranked by predicted probability *p* as responders (Fig. 2d) [58], and the OR at a threshold of 0.5 among the top 20–100% of patients ranked by |*p* − 0.5| (Fig. 2e).

For biomarker profiling, stage and grade were modeled as slide-level classification tasks with cross-entropy loss and evaluated by macro-averaged one-versus-rest AUC. TME biomarkers and gene expression were modeled as regression tasks with mean absolute error (MAE) loss and evaluated by PCC. The eight TME biomarkers were derived using standardized rules: PD-L1 from CD274; CD8/CD4 as mean(CD8A, CD8B) minus mean(CD4, IL7R); TILs from xCell deconvolution of bulk RNA-seq [60, 61]; Ki-67 from MKI67; TMB as nonsynonymous mutations per megabase [62, 63]; MSI from clinical records [64]; HRD from loss of heterozygosity, telomeric allelic imbalance, and large-scale transitions [65, 66]; and Hypoxia from a gene signature [67]. Expression values were mapped to log_2_(TPM+1), TMB was log_10_-transformed, and biomarkers were Z-standardized within each cohort. Gene expression prediction used 15,717 genes; in CPTAC cohorts, evaluation used the genes shared with TCGA. A prognostic-enriched panel of 1,000 genes was additionally selected from 57,214 genes using Cox-derived HRs and *P* values via the ranking score 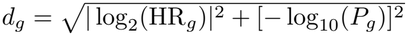, independent of model outputs (Fig. 3i).

For uncertainty-based case selection (TEAM*), we performed two complementary comparisons at the coverage defined by TEAM* in each cohort. In the identical-population comparison, all models were evaluated on the same cases retained by TEAM*. In the model-specific matched-coverage comparison, cases were ranked separately for each baseline within each task and cohort by prediction instability (defined below), and the most unstable cases were excluded until the baseline retained the same number of cases as TEAM*. For all baseline models and tasks, test-time dropout perturbation was applied to the output of the last hidden layer of the downstream MLP immediately before the final linear output layer, with a dropout rate of 0.3 and 20 stochastic forward passes per slide. Slide-level prediction instability was defined for each task: for prognostic stratification, the standard deviation of the risk score across stochastic passes; for therapeutic response prediction, the standard deviation across passes of the response probability, obtained after sigmoid or softmax transformation in each pass; for cancer staging/grading, the variance of each class probability across passes, averaged across classes; for TME characterization and gene expression prediction, the median across markers or genes of the standard deviation of each marker or gene across passes, normalized by the corresponding training-set target standard deviation. Performance on the retained cases was calculated using the original deterministic predictions.

For conversational clinical reasoning and clinician-in-the-loop refinement, the TEAM-V encoder replaced baseline vision encoders. Conversational reasoning was assessed on closed-ended benchmarks (PathVQA, QuiltVQA, PathMMU-EduContent, and PathMMU-PubMed) using accuracy and macro-F1. Open-ended PathQABench responses were anonymized, shuffled, and ranked by five expert pathologists on instruction adherence, clinical accuracy, completeness, brevity, and terminology usage, with rankings aggregated by Borda count [68].

### Statistical analysis

Unless otherwise specified, point estimates are reported with 95% confidence intervals (CIs) computed by nonparametric bootstrapping with 1,000 resamples [69]. Paired model comparisons were evaluated using two-sided Wilcoxon signed-rank tests, with Wilcoxon effect sizes reported where applicable. Risk scores of different cancer types (Fig. 1d) were compared using two-sided Wilcoxon rank-sum tests. Within-cohort 95% CIs were estimated from 1,000 paired bootstrap resamples of slides, preserving the pairing of predictions between models. Mean paired differences across evaluation units are reported with 95% CIs from 1,000 bootstrap resamples of evaluation units. The corresponding *P* values were obtained from two-sided Wilcoxon signed-rank tests on one paired estimate per unit. As a sensitivity analysis, slide-level predictions were averaged per patient and follow-up record in the 21 prognostic cohorts in which some patients contributed more than one slide with the same follow-up record (Extended Data Fig. 9b). For associations between model risk scores and censored survival outcomes, Harrell’s C-index and Cox proportional hazards models were used as the primary analyses. Spearman correlations between risk scores and observed OS time (Fig. 2c) were restricted to cases with observed events. For survival analyses, group differences were assessed using log-rank tests, and Cox proportional hazards models were used to evaluate biomarker-outcome associations in the TME and gene modules, with hazard ratios (HRs) reported. Gene set enrichment analyses reported normalized enrichment scores (NES) and Benjamini–Hochberg-adjusted false discovery rate (FDR) *q* values [70]. For open-ended clinical reasoning evaluation, pathologist rankings were aggregated using the Borda count method [68], and pairwise comparisons were summarized by win rates among non-tied rankings. Inter-rater consistency was assessed using ICC(3,1) (two-way mixed, single measures, consistency) [47].

## Data Availability

Access to in-house cohorts is governed by institutional data use agreements and may be requested from the corresponding authors subject to ethics approval.

https://portal.gdc.cancer.gov/

## Computing software and hardware

Experiments were implemented in PyTorch (v2.1.0) with CUDA 12.1, using OpenSlide (v1.2.0) and CLAM [71] for WSI processing, with Torchmetrics (v1.3.2) and scikit-learn (v1.2.2) for evaluation. TEAM was pretrained on five NVIDIA RTX 5880 Ada GPUs with mixed-precision distributed training. Full specifications are provided in Supplementary Tables 2–5.

## Ethics approval

This retrospective study of de-identified archival pathology slides and associated clinical data was conducted in accordance with the Declaration of Helsinki. Formal ethics approvals and waivers of informed consent for the retrospective use of archival histopathology slides and associated clinicopathological variables were obtained as follows: NFH-CRC (Ethics Committee and Institutional Review Board of Nanfang Hospital, Southern Medical University; approval nos. NFEC-2022-428 and NFEC-2025-092); QDUH-BRCA, SUQH-BRCA, and SHSU-BRCA (Ethics Committee of the School of Basic Medical Sciences, Shandong University; approval no. ECSBMSSDU2024-1-179); ZJH-LIHC (Medical Ethics Committee of Zhujiang Hospital, Southern Medical University; approval no. 2026-KY-135-01); QDPH-HNSC (Medical Ethics Committee of the Affiliated Hospital of Qingdao University; approval no. QYFYEC2023.56); and QDPH-BLCA (Medical Ethics Committee of the Affiliated Hospital of Qingdao University; approval no. ZYFYWZLL29745). All slides were de-identified at the source institution prior to transfer, and no patient identifiers were retained in the analysis dataset. Public datasets were used in accordance with their data use policies and ethical approvals.

## Data availability

Public datasets used for pretraining and downstream evaluation are listed in Supplementary Table 22. Access to in-house cohorts is governed by institutional data use agreements and may be requested from the corresponding authors subject to ethics approval.

## Code availability

Source code, pretrained model weights, and the interactive web platform are publicly available at https://github.com/juruoxcj/TEAM.

## Acknowledgements

We thank Yuting Chen for assistance with figure visualization. Generative AI tools were used to improve the language and readability of the manuscript. The authors reviewed and edited all text and take full responsibility for the content. This work was supported in part by the National Natural Science Foundation of China under Grant 82402363, Grant U22A20350, and Grant 82472056, in part by the Basic and Applied Basic Research Foundation of Guangdong Province under Grant 2025A1515011205 and Grant 2024A1515012004, in part by the Guangdong Provincial Key Laboratory of Medical Image Processing under Grant No. 2020B1212060039, in part by the Science and Technology Program of Guangzhou under Grant No. 2025A04J4019, in part by the Guangdong Association for Science and Technology Young Talents Training Program under Grant SKXRC2025372, in part by the Special Funds for the Cultivation of Guangdong College Students’ Scientific and Technological Innovation under Grant No. pdjh2023a0099 and Grant No. pdjh2024a087, and in part by the Research Support Fund for High-level Talents of Southern Medical University under Grant No. G624300273.

## Author contributions

Y.M., C.X., Y.Z., and Z.N. conceived and designed the study. Y.M., C.X., and F.L. developed and implemented the methodology. C.X. and F.L. performed model training and experiments. Y.M., C.X., F.L., and W.Z. conducted statistical analyses and generated visualizations. Y.M., C.X., Y.D.Z., H.T., J.Y., J.W., L.L., and C.L. curated the datasets. D.L., B.B.L., C.L.Z., Z.Y.Z., Y.T., and Z.C. performed expert pathological evaluation and contributed to interpretation of the findings. Y.M., D.L., W.Z., Y.Z., and Z.N. interpreted the results. Q.F., B.L., L.L., C.L., Y.Z., and Z.N. supervised the study. Y.Z. and Z.N. acquired funding. Y.M. and C.X. drafted the manuscript. B.L., L.L., C.L., Y.Z., and Z.N. revised the manuscript. All authors reviewed and approved the final manuscript.

## Supplementary information

Extended Data Figures and Supplementary Information (including Supplementary Figures, Tables, and Supplementary Movie 1) are provided with this manuscript.

## Extended Data for

An Interactive Trustworthy AI Pathology Copilot to Improve Biomarker-Driven Prognostic Stratification and Therapeutic Response Prediction

**Extended Data Fig. 1.**
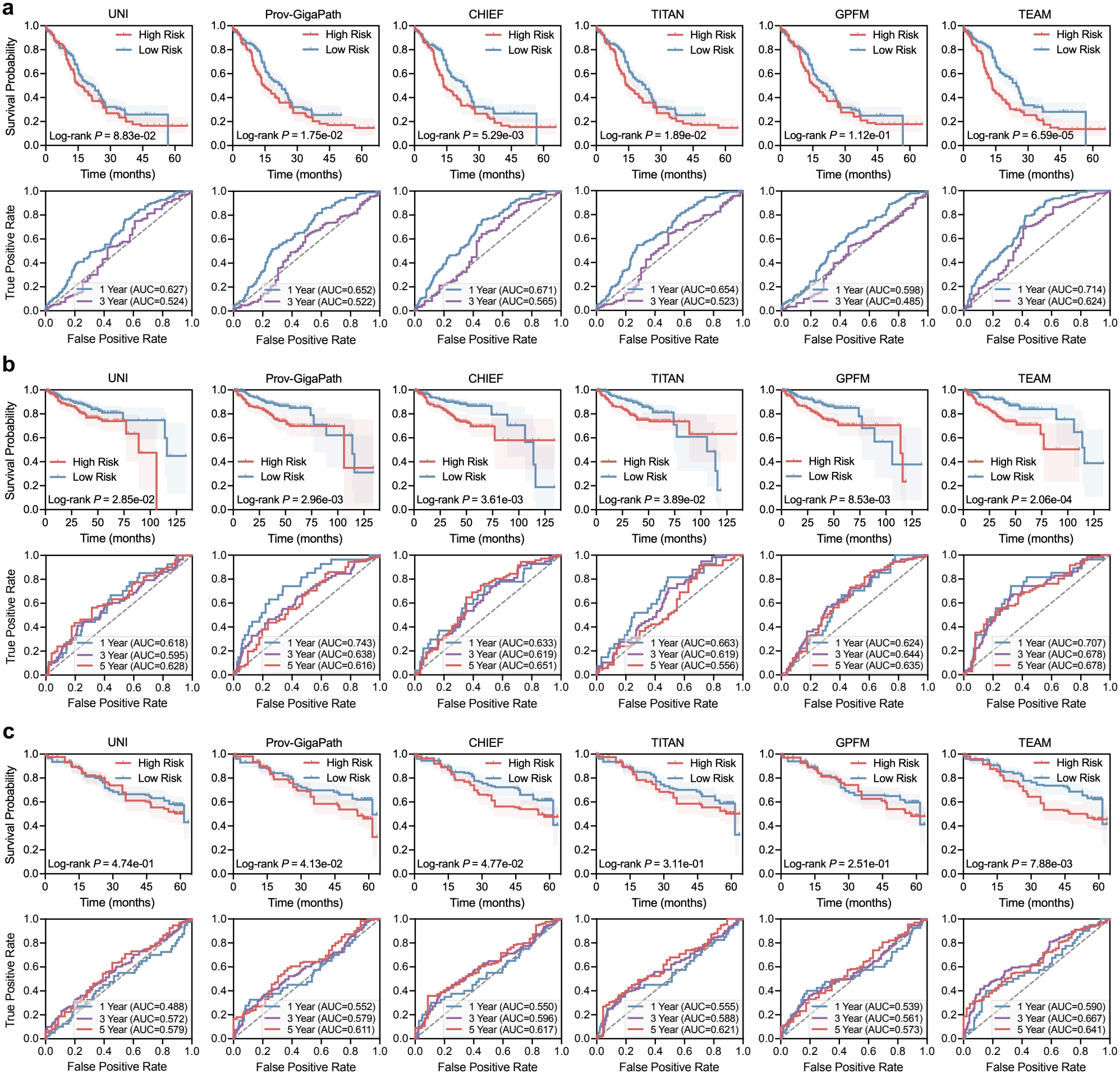
Extended prognostic validation across additional cancer cohorts. Prognostic analysis for the CPTAC-GBM (**a**), QDPH-BLCA (**b**), and CPTAC-LUSC (**c**) cohorts. Top: Kaplan–Meier curves comparing low- and high-risk strata produced by each model (shaded area, 95% CI; *P* values from two-sided log-rank tests). Bottom: time-dependent ROC curves at 1, 3, and 5 years. The 5-year curve is not shown for CPTAC-GBM, in which five patients had follow-up beyond 5 years.

**Extended Data Fig. 2.**
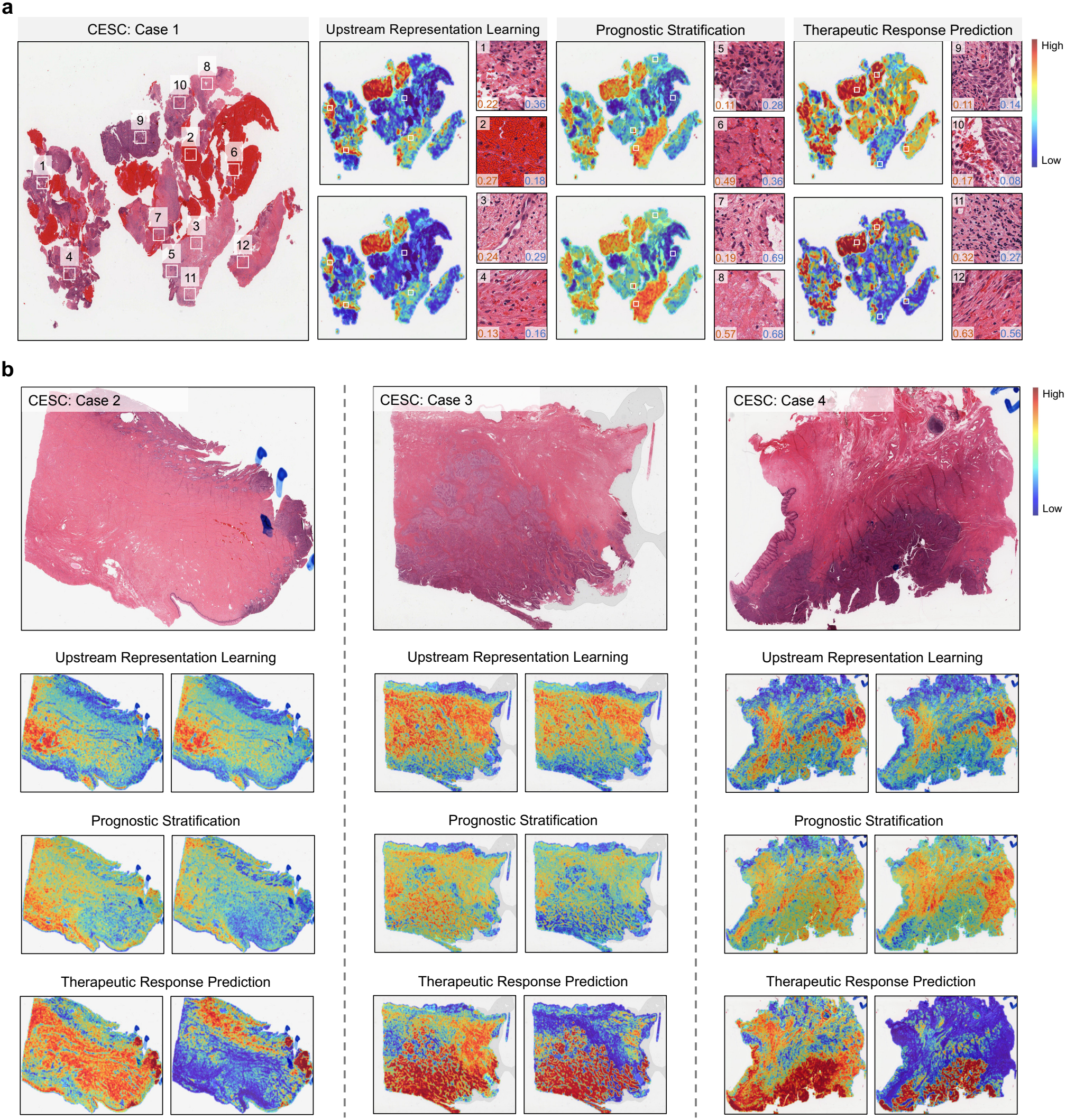
Extended visualization for patient outcome prediction. **a**, Whole-slide attention heatmaps across three tasks (left to right: upstream representation learning, prognostic stratification, therapeutic response prediction). For each task, the upper panel shows raw attention weights and the lower panel shows uncertainty-weighted attention. Four representative patches per task are annotated with aleatoric (orange) and epistemic (blue) uncertainty values. **b**, Three additional whole-slide examples with attention heatmaps.

**Extended Data Fig. 3.**
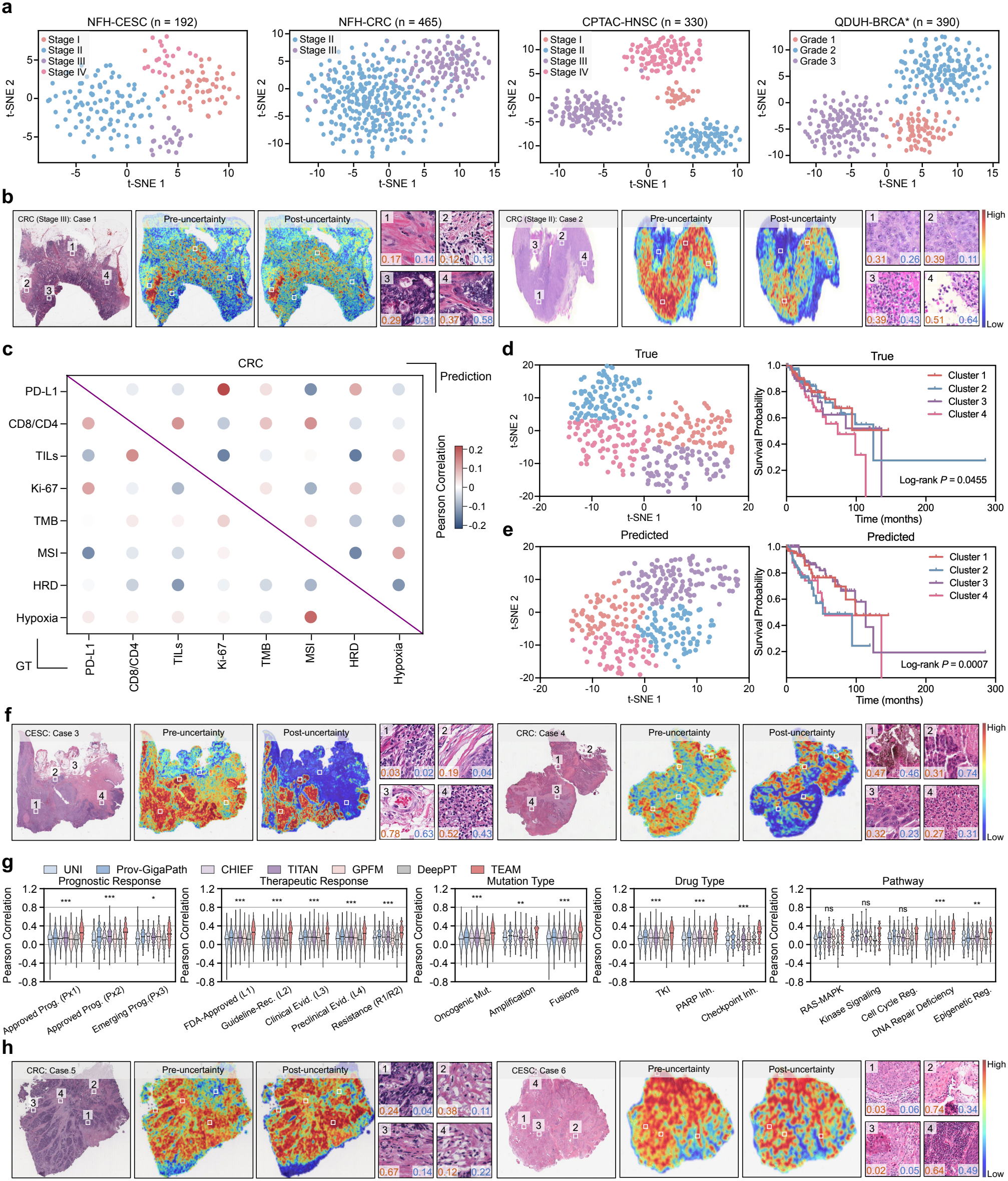
Extended visualization for multi-scale biomarker profiling. **a**, t-SNE of TEAM slide-level embeddings colored by stage/grade. **b**, Additional staging slides (stages II and III) with uncertainty overlay. **c**, Pairwise Pearson correlations among eight TME biomarkers in CRC (lower triangle, ground truth; upper triangle, TEAM-predicted values). **d**,**e**, t-SNE embeddings constructed from eight TME biomarkers using ground truth (d) and TEAM-predicted (e) values, with K-means clusters and corresponding Kaplan–Meier curves. **f**, Additional TME heatmap examples across cancer types. **g**, Gene-wise PCC for gene expression prediction stratified by phenotype category, for the seven models of Fig. 3j under the same regression head (17 cohorts pooled). TEAM was compared with the best-performing baseline in each subgroup using two-sided Welch’s t-tests (\**P*<0.05, \*\**P*<0.01, \*\*\**P*<0.001; ns, *P≥*0.05). **h**, Additional gene expression attention maps across cancer types.

**Extended Data Fig. 4.**
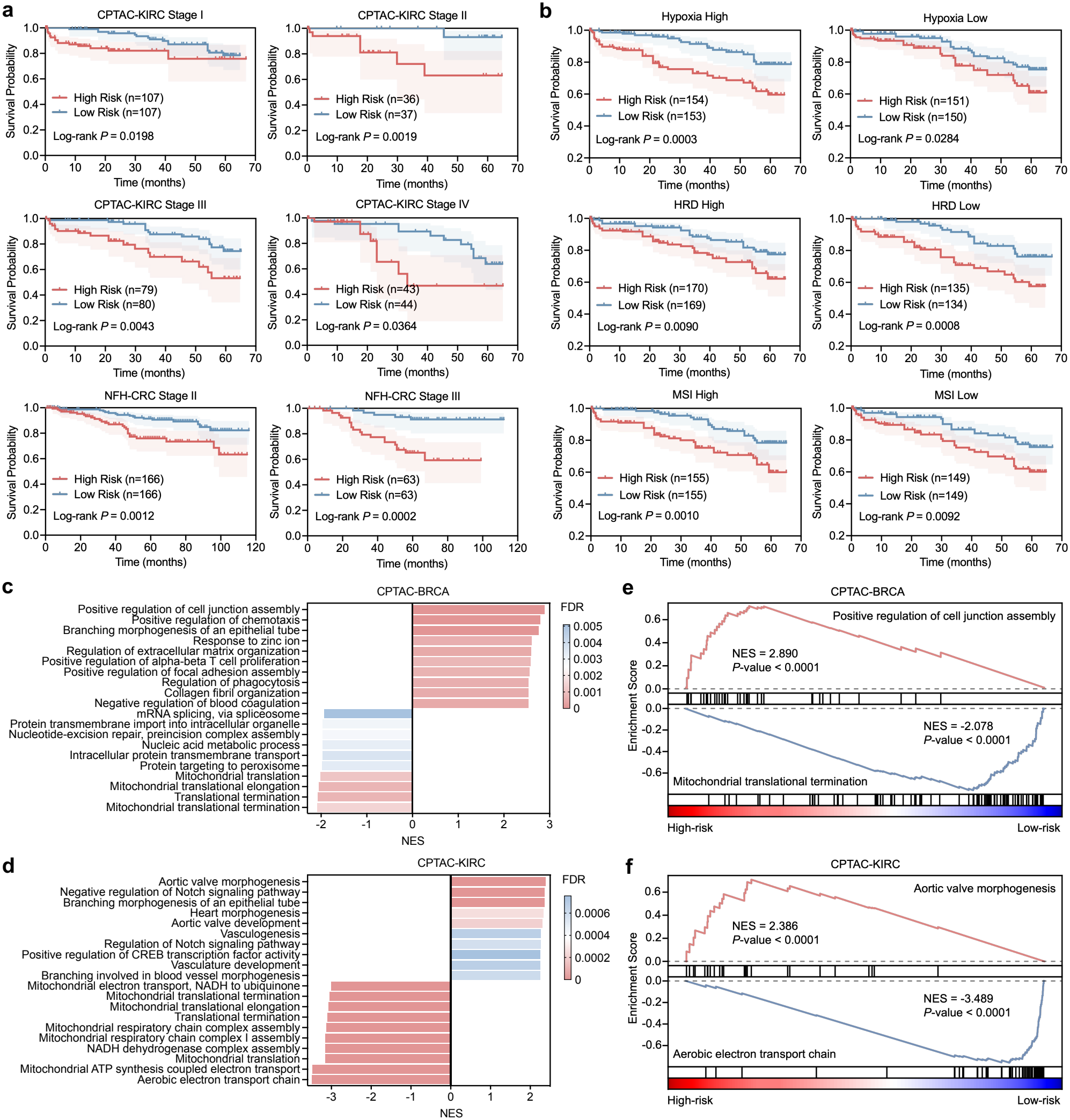
Extended validation of prognostic information beyond individual biomarkers. **a**, Within-stage risk stratification by TEAM in CPTAC-KIRC (stages I–IV) and NFH-CRC (stages II–III) cohorts. Kaplan–Meier curves show highversus low-risk groups stratified by TEAM-predicted risk scores within each clinical stage (*P* values from two-sided log-rank tests). **b**, Risk stratification by TEAM within TME biomarker subgroups in the CPTAC-KIRC cohort. Kaplan–Meier curves show high-versus low-risk groups stratified by TEAM-predicted risk scores within subgroups with high or low hypoxia, HRD, or MSI levels (median split). **c**,**d**, Gene set enrichment analysis (GSEA) of Gene Ontology (GO) Biological Process gene sets for CPTAC-BRCA (c) and CPTAC-KIRC (d), with genes ranked by their correlation with TEAM-predicted risk scores; the top enriched pathways are shown. **e**,**f**, Representative enrichment plots for CPTAC-BRCA (e) and CPTAC-KIRC (f), showing a pathway enriched among genes positively correlated with risk scores (top) and one enriched among genes negatively correlated with risk scores (bottom).

**Extended Data Fig. 5.**
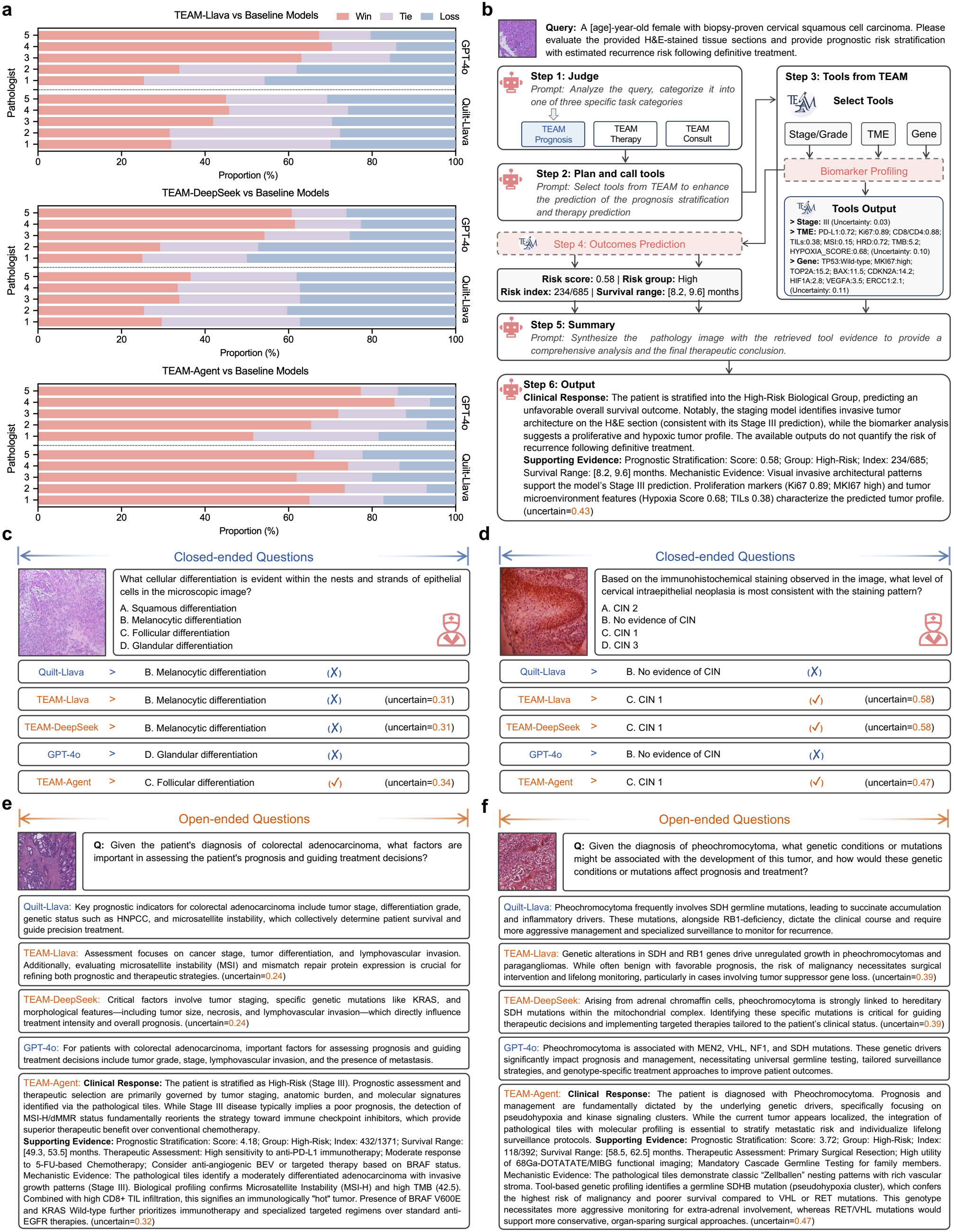
Extended evaluation of agentic orchestration and conversational clinical reasoning. **a**, Performance comparison of TEAM-Llava, TEAM-DeepSeek, and TEAM-Agent versus baselines (GPT-4o, Quilt-Llava) evaluated by five expert pathologists on open-ended PathQABench questions. Win, tie, and loss rates are shown as proportions. **b**, Complete agentic workflow of TEAM-Agent comprising six sequential steps from query classification and tool orchestration to outcome prediction and evidence synthesis. **c**,**d**, Representative closed-ended question-answering examples. **e**,**f**, Representative open-ended question-answering examples.

**Extended Data Fig. 6.**
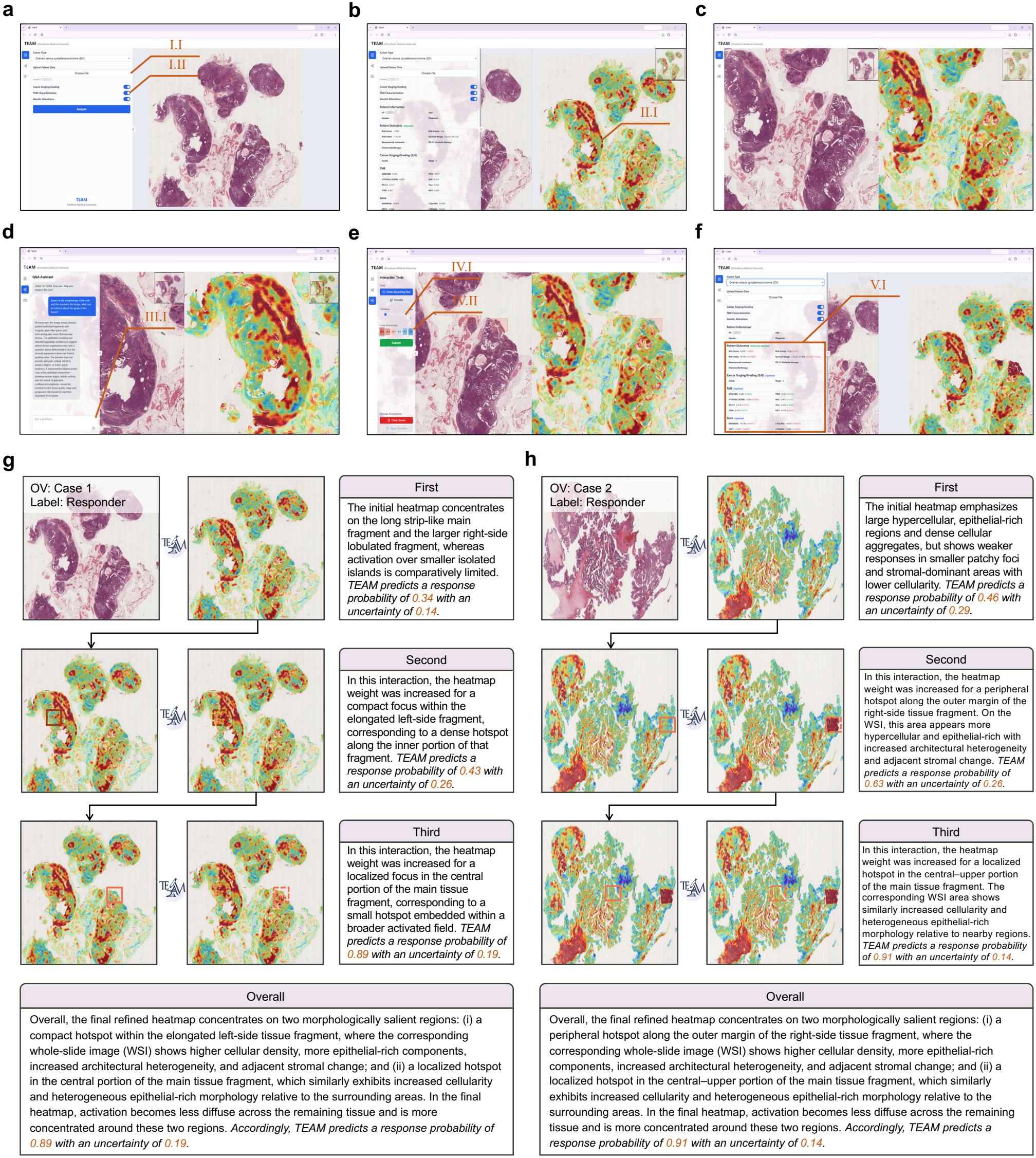
Extended demonstration of clinician-in-the-loop interactive prediction refinement. **a–f**, Step-by-step demonstration of the TEAM interactive interface: **(a)** cancer-type selection and whole-slide image upload (I.I) and biomarker orchestration (I.II); **(b)** initial model inference with attention heatmap visualization (II.I); **(c)** high-resolution whole-slide attention visualization; **(d)** TEAM-Agent chat interface for pathologist queries and feedback (III.I); **(e)** interaction tools for choosing the annotation mode (bounding box or freehand scribble; IV.I) and adjusting attention weights (IV.II); **(f)** refined results with updated predictions (V.I). **g**,**h**, Two representative examples illustrating iterative heatmap refinement for therapeutic response prediction. Each example shows the initial prediction (First) and two rounds of refinement (Second, Third) with corresponding descriptions of attention weight adjustments, response probabilities, and uncertainty values. Top row: original WSI (left) and initial attention heatmap (right). Middle and bottom rows: attention heatmap before (left, solid box marking the selected region) and after (right, dashed box) the weight adjustment. Bottom: overall summary describing how the final refined heatmap concentrates on morphologically salient regions identified through pathologist guidance. During the interaction, ground truth and uncertainty estimates were hidden from the pathologist, and the uncertainty values shown here were presented only after refinement (Supplementary Fig. 4).

**Extended Data Fig. 7.**
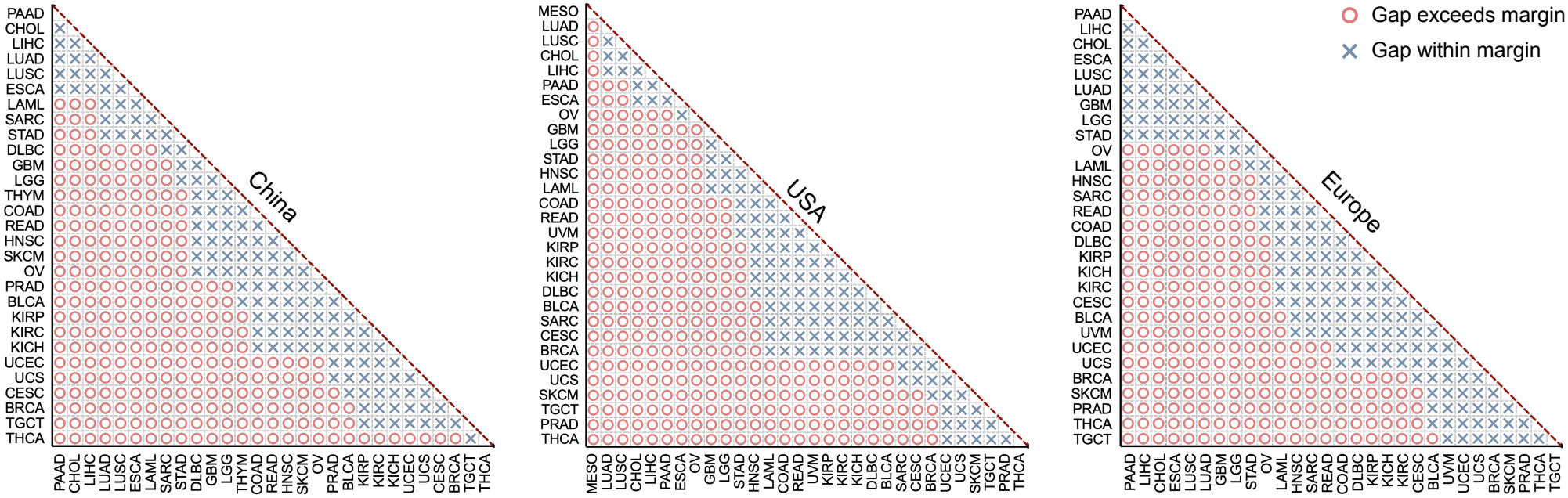
Within-registry pairwise survival gap analysis for registry-specific prior derivation. Triangular matrices showing pairwise within-registry survival gap comparisons across cancer types in China, the United States, and Europe. Red circles indicate pairs with survival gaps exceeding the registry-specific margin, used to derive weak supervision signals. Blue crosses indicate pairs with gaps within the margin.

**Extended Data Fig. 8.**
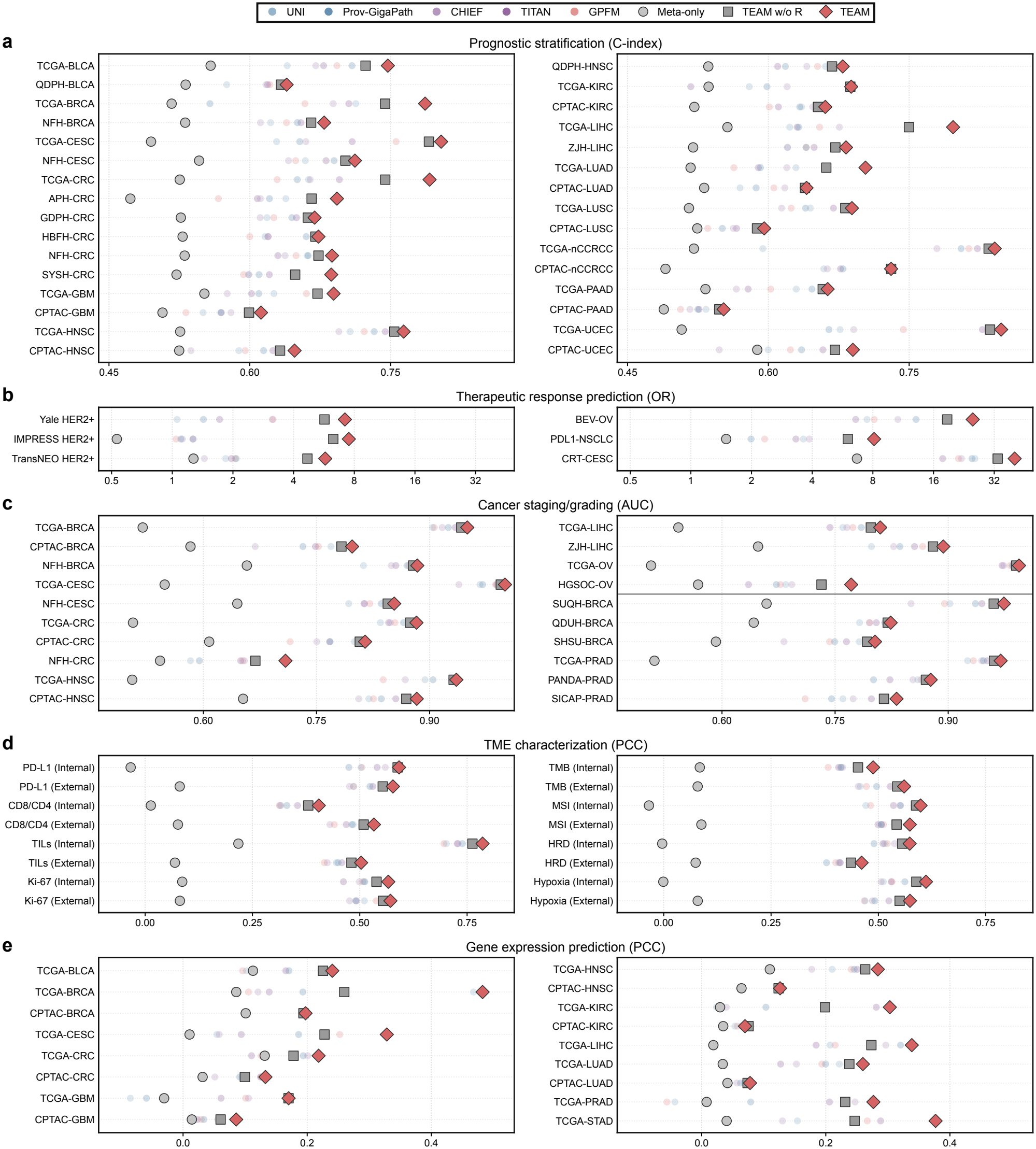
TEAM-R component and baseline comparisons across downstream tasks. Comparison of TEAM, TEAM w/o R, Meta-only, and the five image-only baselines in **a**, prognostic stratification (C-index); **b**, therapeutic response prediction (OR); **c**, cancer staging/grading (AUC); **d**, TME characterization (PCC); and **e**, gene expression prediction (PCC). Meta-only is not shown for cohorts without any of the four demographic variables (Yale HER2+, BEV-OV, PANDA-PRAD, and SICAP-PRAD).

**Extended Data Fig. 9.**
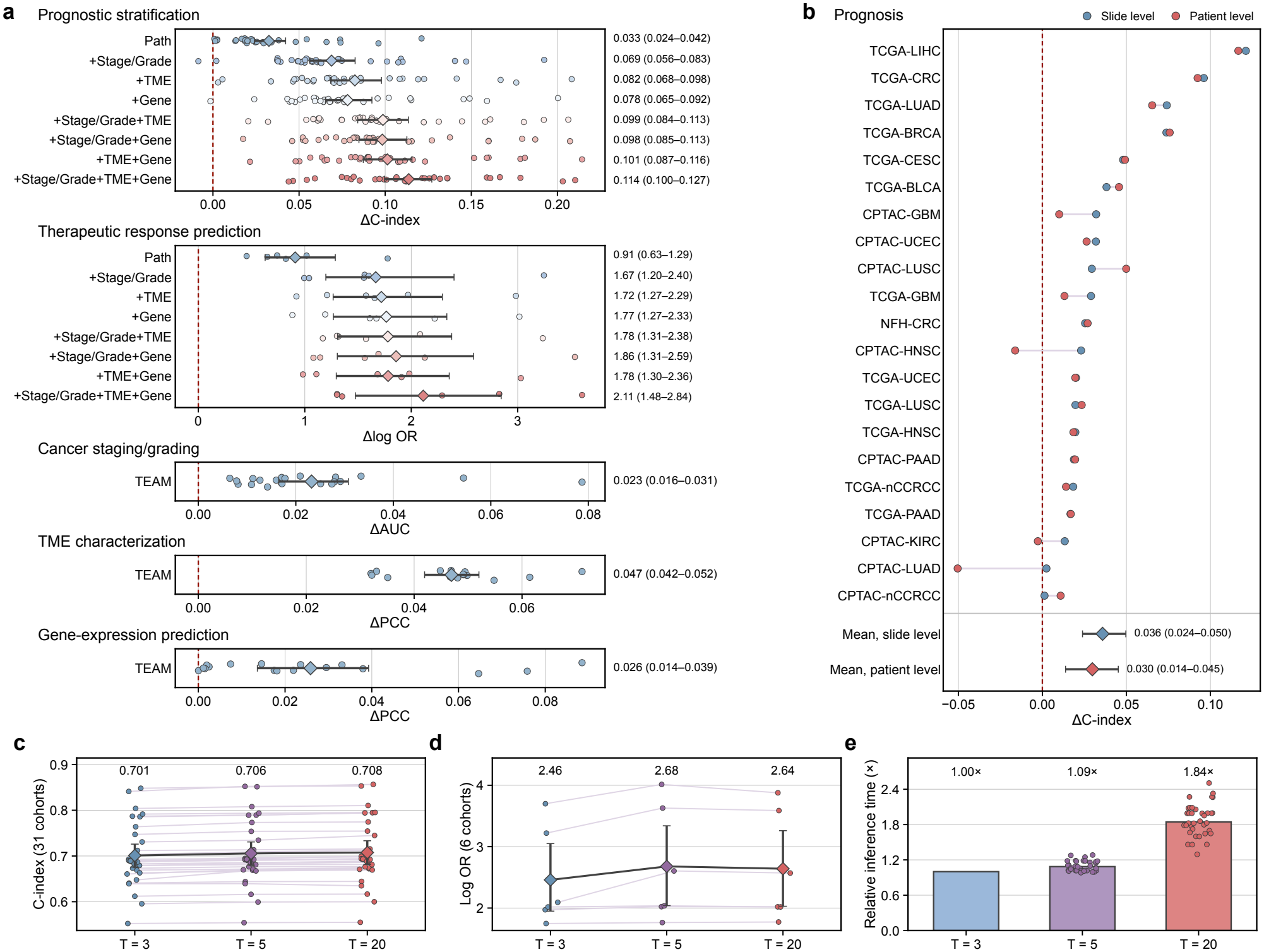
Effect size and sensitivity analyses. **a**, Paired differences between each model and the unit-wise best image-only baseline in each downstream task; points, evaluation units; diamonds, means with 95% CIs from 1,000 unit-level bootstrap resamples; right, mean (95% CI). Prognostic stratification and therapeutic response prediction show the eight biomarker-driven model configurations; the other tasks show TEAM. **b**, Difference in C-index between TEAM and the best-performing baseline at the slide level and after averaging slides per patient and follow-up record, in the 21 prognostic cohorts in which some patients contributed more than one slide with the same follow-up record (6,261 slides from 2,958 patients); points, cohorts (blue, slide level; red, patient level); diamonds, means with 95% CIs. **c**–**e**, Sensitivity to the number of Monte Carlo dropout passes *T* . **c**, C-index on 31 prognostic cohorts. **d**, Log OR on 6 therapeutic cohorts. Points and lines, cohorts; diamonds, means with 95% CIs; numbers above, means. **e**, Inference time relative to *T* = 3; bars and numbers above, total over all test sets; points, individual test sets.

**Extended Data Fig. 10.**
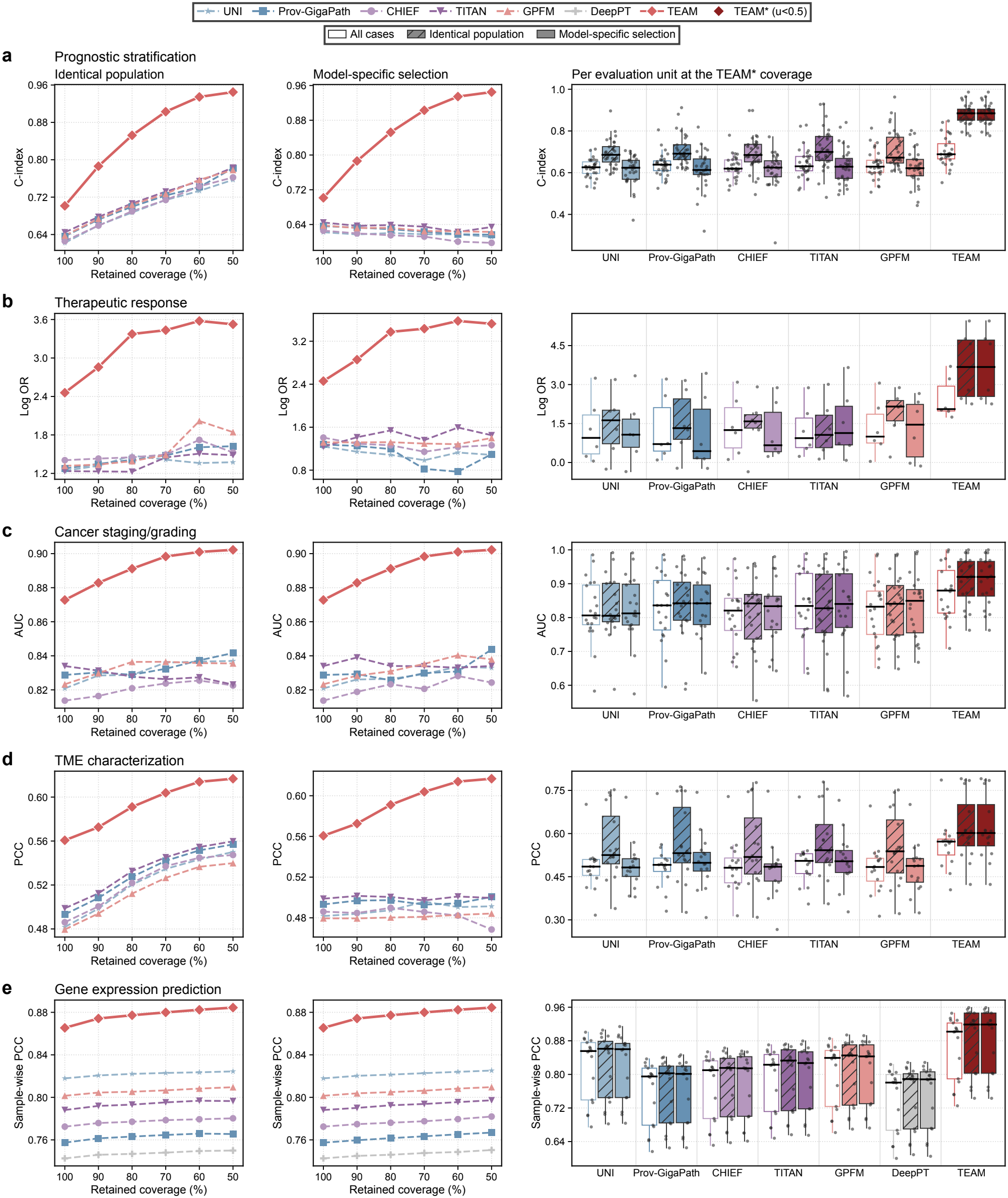
Two complementary comparisons at matched coverage. **a**, Prognostic stratification (31 cohorts); **b**, therapeutic response prediction (6 cohorts); **c**, cancer staging/grading (20 cohorts); **d**, TME characterization (eight markers, internal and external); **e**, gene expression prediction (17 cohorts, sample-wise PCC). Left, identical population (cases retained by TEAM*), and middle, model-specific selection, each from 100% to 50% retained coverage; right, per-unit distributions at the TEAM* coverage (boxes, median and IQR; whiskers, 1.5 *×* IQR).

