## Supplementary Information for "An Interactive Trustworthy AI Pathology Copilot to Improve Biomarker-Driven Prognostic Stratification and Therapeutic Response Prediction"

### Contents


**Supplementary Figures** **3**

**Prompt Sets for TEAM-Agent** **6**

**Supplementary Tables** **9**


### Supplementary Figures


| | | |
|---|---|---|
| 1 | t-SNE representation ablation. | 3 |
| 2 | Stage-adjusted prognostic analyses in the 23 cohorts with recorded tumor stage. | 3 |
| 3 | Relative-risk-prior ablation across downstream tasks. | 4 |
| 4 | Schematic of the clinician-in-the-loop workflow. | 4 |
| 5 | Gene expression prediction by the seven models of Fig. 3j. | 5 |


### Supplementary Tables


| | | |
|---|---|---|
| 1 | Organ-system distribution of pretraining data. | 10 |
| 2 | TEAM architecture specifications. | 11 |
| 3 | TEAM pretraining configuration. | 12 |
| 4 | Vision–language alignment training details. | 13 |
| 5 | Hyperparameters for ViT-L/16 pretraining with DINOv2. | 14 |
| 6 | TEAM-R input variables and representative input examples. | 15 |
| 7 | Availability of TEAM-R input variables during pretraining. | 16 |
| 8 | Cancer subtype risk rankings. | 16 |
| 9 | In-house validation cohorts. | 18 |
| 10 | Organ-system distribution of downstream evaluation data. | 19 |
| 11 | Distribution of prognostic stratification datasets. | 20 |
| 12 | Distribution of patient clinical metadata. | 21 |
| 13 | Distribution of therapeutic response datasets. | 22 |
| 14 | Distribution of cancer staging datasets. | 22 |
| 15 | Distribution of cancer grading datasets. | 23 |
| 16 | Distribution of tumor microenvironment (TME) datasets. | 23 |
| 17 | Distribution of immune surveillance profile samples. | 24 |
| 18 | Distribution of TME stress profile samples. | 24 |
| 19 | Distribution of gene expression datasets. | 25 |
| 20 | Distribution of question-answering model training datasets. | 25 |
| 21 | Distribution of question-answering model testing datasets. | 26 |
| 22 | Data availability and links to the datasets. | 26 |
| 23 | Task-specific implementation of the compared pathology models. | 29 |
| 24 | Implementation settings of the compared pathology models. | 30 |

| | | |
|---|---|---|
| 25 | Availability of TEAM-R variables for the Meta-only evaluation. | 30 |
| 26–56 | Performance of different modality combinations for prognostic stratification (31 cohorts). | 32 |
| 57–62 | Performance of different modality combinations for therapeutic response prediction (six cohorts). | 48 |
| 63 | Attention weight allocation for the prognostic stratification task. | 52 |
| 64 | Attention weight allocation for the therapeutic response prediction task. | 53 |
| 65–78 | Performance of different foundation models for cancer staging (14 cohorts). | 53 |
| 79–84 | Performance of different foundation models for cancer grading (six cohorts). | 58 |
| 85–100 | Performance of different foundation models for TME characterization (eight markers, internal and external validation). | 60 |
| 101–117 | Performance of different models for gene expression prediction (17 cohorts). | 65 |
| 118 | Cohort-level gene expression comparison under a common regression head. | 74 |
| 119 | Model comparison at matched coverage across downstream tasks. | 74 |
| 120 | TEAM on all cases and on TEAM*-retained cases. | 76 |
| 121 | Paired differences between TEAM and the best-performing baseline of each evaluation unit. | 77 |
| 122 | Sensitivity to $T$ by task. | 78 |
| 123–126 | Performance of different vision-language models for visual question answering (four datasets). | 78 |
| 127 | Overall performance of vision-language models evaluated by clinicians using Borda count method. | 80 |
| 128 | Head-to-head comparison of TEAM-based variants against baseline models. | 80 |
| 129 | Quantitative assessment of clinician-in-the-loop refinement across clinical tasks. | 81 |
| 130 | Pathologist interaction frequency during the model refinement process. | 82 |
| 131 | Cancer type abbreviations. | 83 |

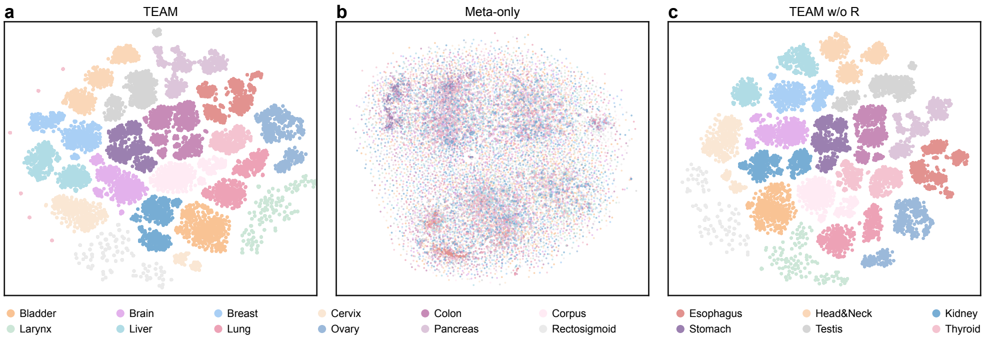


**Supplementary Fig. 1. t-SNE representation ablation.** t-SNE projections of the TCGA embeddings in Fig. 1e from TEAM (**a**), Meta-only (**b**), and TEAM w/o R (**c**), colored by organ site.

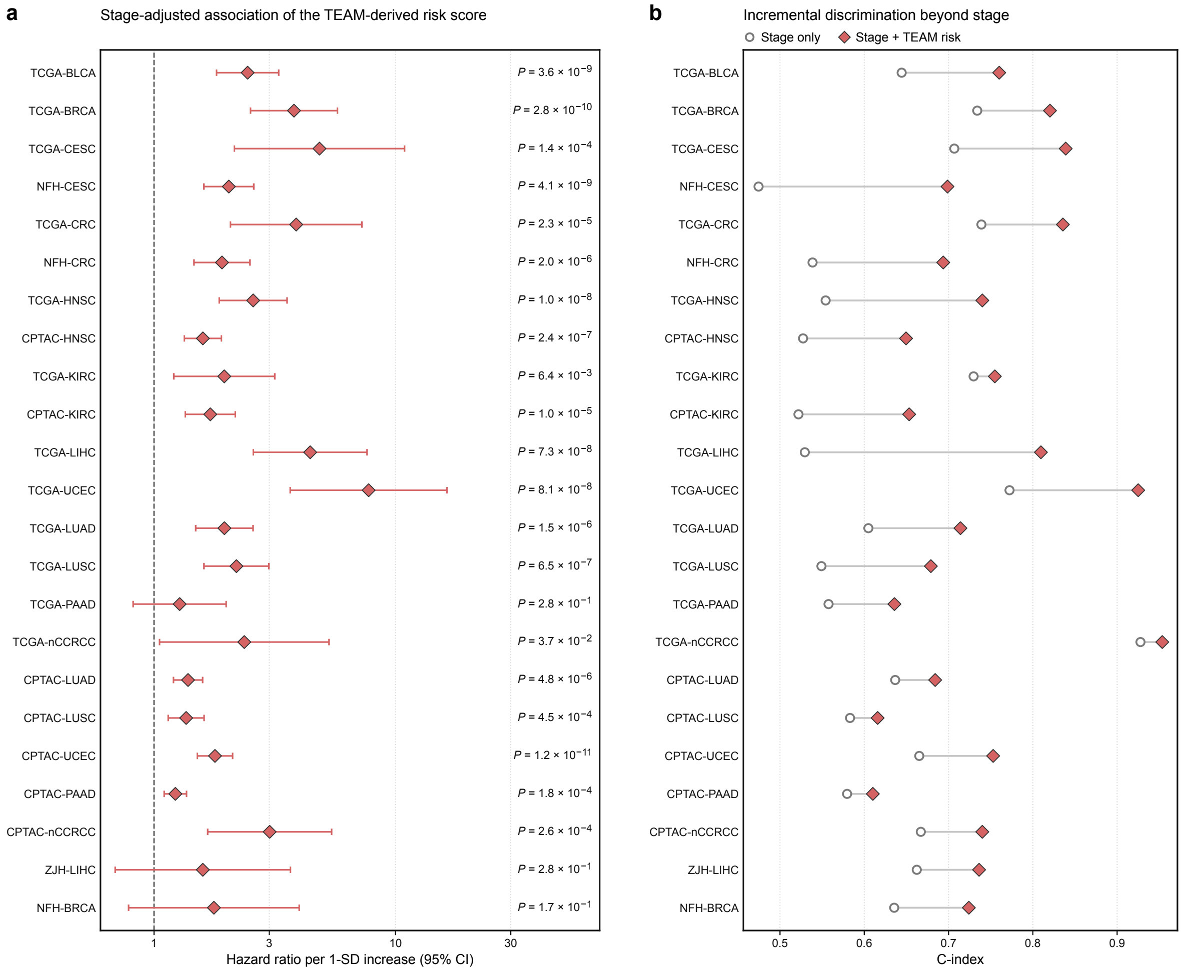


**Supplementary Fig. 2. Stage-adjusted prognostic analyses in the 23 cohorts with recorded tumor stage. a**, Hazard ratios per 1-s.d. increase in the TEAM-derived risk score from stage-adjusted Cox models (error bars, 95% CI; two-sided Wald $P$ values). **b**, C-indices of stage-only and stage-plus-risk-score Cox models; lines connect the same cohort.

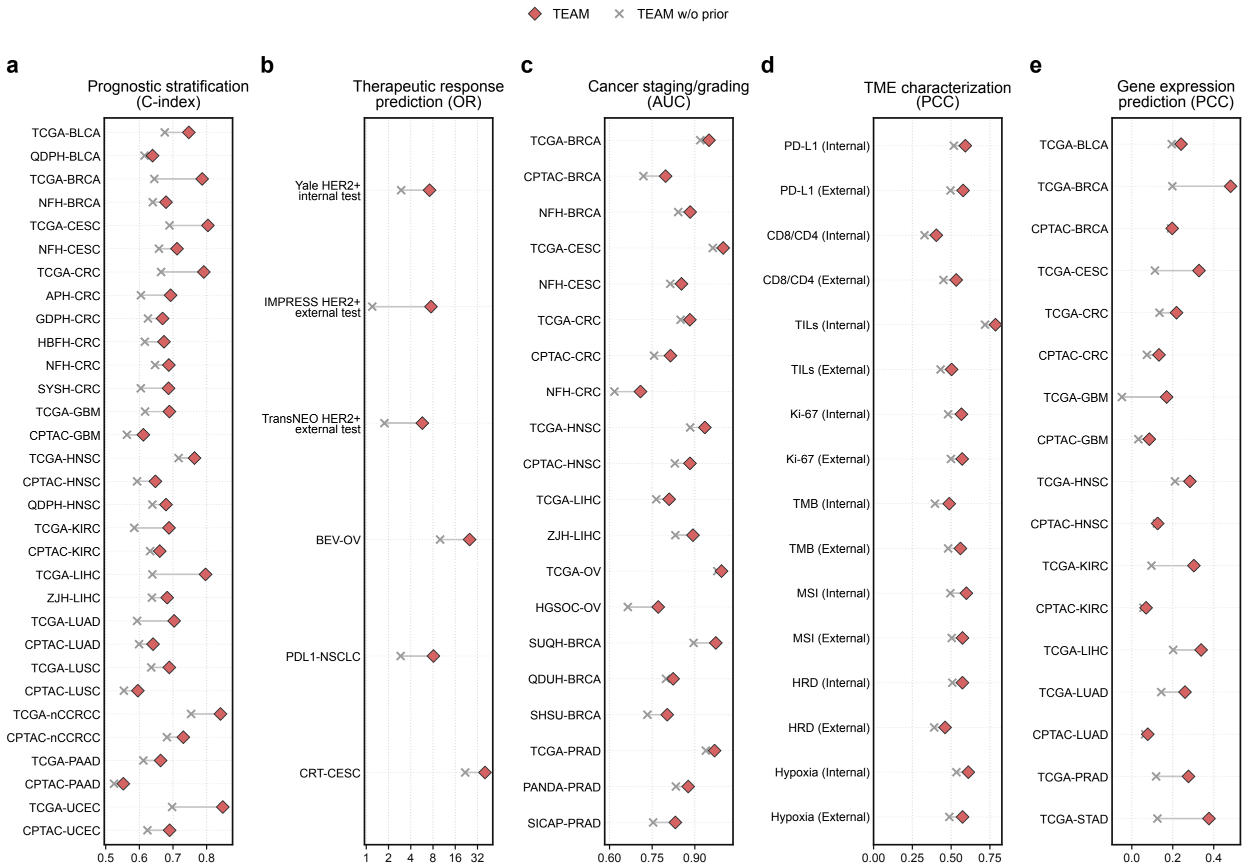


**Supplementary Fig. 3. Relative-risk-prior ablation across downstream tasks.** TEAM (red diamonds) and TEAM w/o prior (crosses) on each evaluation unit; gray lines connect the same unit. **a**, Prognostic stratification (C-index, 31 cohorts). **b**, Therapeutic response prediction (mean OR, log scale; 6 cohorts). **c**, Cancer staging/grading (AUC, 20 cohorts). **d**, TME characterization (PCC, eight markers, internal and external). **e**, Gene expression prediction (mean gene-wise PCC, 17 cohorts).

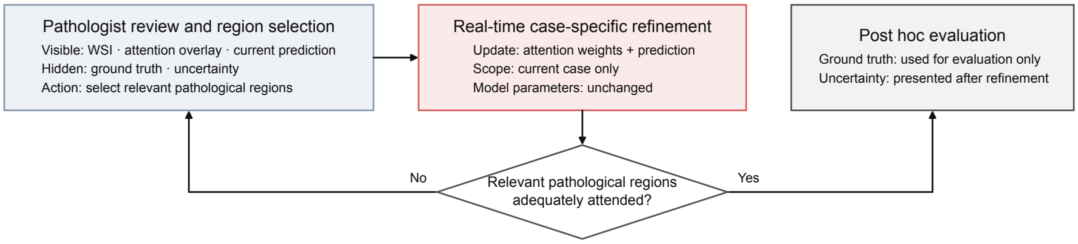


**Supplementary Fig. 4. Schematic of the clinician-in-the-loop workflow.**

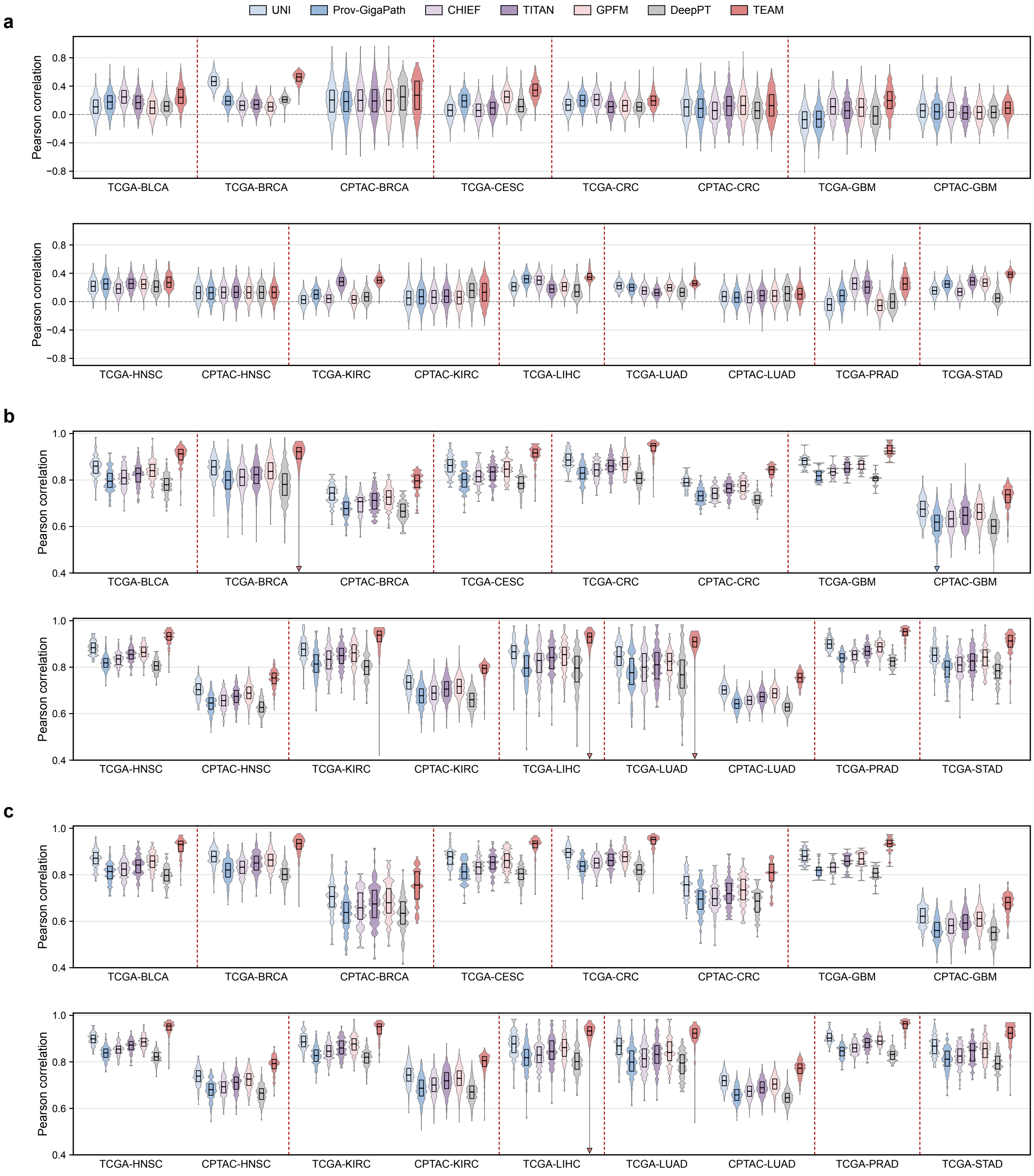


**Supplementary Fig. 5. Gene expression prediction by the seven models of Fig. 3j. a**, Gene-wise PCC for the top 1,000 prognostically associated genes. **b**, Sample-wise PCC across all genes. **c**, Sample-wise PCC for the top 1,000 prognostically associated genes. Violins, distributions across genes (**a**) or samples (**b**, **c**); boxes, interquartile range; center lines, medians; dashed lines separate cancer types; triangles, values below the axis range.

### Prompt Sets for TEAM-Agent

Prompt

**Prompt 1. Query Classification (Step 1: Judge)**
**Objective:** Analyze the user's natural language query and categorize it into one of three specific task categories for routing to the appropriate analysis team.
You are a Medical Triage Agent. Your sole purpose is to route the user's query to the correct specialized analysis team (TEAM_PROGNOSIS, TEAM_THERAPY, or TEAM_CONSULT).
**Task Categories:**

1. `TEAM_PROGNOSIS`: For queries regarding survival prediction, risk stratification, or disease progression.
2. `TEAM_THERAPY`: For queries regarding therapeutic response prediction (e.g., BEV, PD-L1, CRT).
3. `TEAM_CONSULT`: For general clinical guidance not requiring outcome prediction, with optional biomarker support.

**Input:**

- **User Query:** `{user_query}`
- **Pathology Image:** `{image}`

**Constraints:**

- Do not provide any conversational text.
- Output valid JSON only.

**Required Output Format (JSON):**

```
{
  "category": "String",  // One of: TEAM_PROGNOSIS, TEAM_THERAPY, TEAM_CONSULT
  "reasoning": "String"  // A concise sentence explaining the classification
}
```

Prompt

**Prompt 2. Tool Planning and Biomarker Profiling (Step 2-3: Plan and Call Tools)**
**Objective:** Based on the classification from Step 1, select tools from TEAM to enhance the prediction of prognosis stratification and therapy prediction.
You are a Clinical Strategy Planner. Based on the task category, you must select the

appropriate biomarker profiling tools and execute them to retrieve structured evidence.

**Context:**

- **User Query:** `{user_query}`
- **Task Category:** `{step1_category}`
- **Pathology Image:** `{image}`

**Available Tools:**

- **Stage Module:** Predicts cancer stage (I-IV) with uncertainty quantification.
- **TME Module:** Profiles tumor microenvironment (PD-L1, CD8/CD4, TILs, Ki-67, TMB, MSI, HRD, Hypoxia).
- **Gene Module:** Predicts gene expression signatures relevant to prognosis and therapy response.

**Instructions:**

1. Analyze which biomarker modules are required to answer the query.
2. Execute the selected tools on the pathology image.
3. Return structured biomarker evidence with uncertainty scores.
4. Pass the image embedding to TEAM for outcomes prediction.

**Required Output Format (JSON):**

```
{
  "plan_summary": "String",     // Brief description of the analysis strategy
  "selected_tools": {
    "stage_module": "Boolean",  // True/False
    "tme_module": "Boolean",    // True/False
    "gene_module": "Boolean"    // True/False
  },
  "tools_output": {
    // Only include outputs for modules where selected_tools is True
    "stage": {"prediction": "String", "uncertainty": "Float"},
    "tme": {"biomarker_name": "Float", ..., "uncertainty": "Float"},
    "gene": {"gene_name": "Float/String", ..., "uncertainty": "Float"}
  },
  "embedding": "Tensor"         // Image embedding for TEAM outcomes prediction
}
```

**Note:** The `tools_output` field only contains results from modules where `selected_tools` is set to `True`. The `embedding` is passed to TEAM for downstream outcomes prediction (prognosis and therapy response).

Prompt

**Prompt 3. Clinical Report Synthesis (Step 5-6: Summary and Output)**
**Objective:** Synthesize the pathology image analysis with the retrieved tool evidence to provide a comprehensive analysis and the final therapeutic conclusion.
You are a Senior Consultant Oncologist. You must synthesize a final clinical report that integrates biomarker evidence with outcome predictions to provide actionable clinical insights.

**Input Data (JSON):**

```
{
  "outcomes": {
    "prognosis": {prognosis_json_object},
    // risk_score, risk_group, risk_index, survival_range
    "therapy": {therapy_json_object}
    // BEV, PD-L1, CRT
  },
  "biomarkers": {
    "stage": {stage_json_object},     // Cancer stage with uncertainty
    "tme": {tme_json_object},         // TME biomarker values with uncertainty
    "gene": {gene_json_object}        // Gene signatures with uncertainty
  }
}
```

**User Query:** `{user_query}`
**Instructions:**

1. **Integrate:** Combine outcome predictions with biomarker evidence.
2. **Validate:** Ensure predictions are mechanistically supported by biomarkers.
3. **Synthesize:** Generate a coherent clinical narrative with therapeutic implications.

**Constraints:**

- Output a structured clinical text report (NOT JSON).
- Use professional medical terminology.
- Provide mechanistic explanations linking biomarkers to outcomes.

**Output Format:**

- Close-ended queries → categorical label only;
- Open-ended queries → structured report below.

**Report Structure** (open-ended):

**Clinical Response:**
[Directly answer the user's query in natural clinical language]

**Supporting Evidence:** (based on task category and selected modules)

- **Prognostic Stratification:** (if applicable)
  [Risk group, risk score, survival estimates]
- **Therapeutic Assessment:** (if applicable)
  [Treatment response prediction with probability]
- **Mechanistic Evidence:** (based on selected modules)
  [Stage / TME / Gene analysis supporting the response]

**Supplementary Table 1. Organ-system distribution of pretraining data.** Counts of whole-slide images (WSIs), regions of interest (ROIs), and tissue patches (in millions, M) across organ systems used for pretraining, with corresponding dataset sources.

| Organ | WSI Count | ROI Count | Patch Count (M) | Datasets |
|---|---|---|---|---|
| Brain | 3,004 | – | 14.79 | TCGA, GTEx |
| Head & Neck | 826 | – | 9.52 | TCGA, GTEx |
| Thyroid | 1,339 | – | 10.55 | TCGA, GTEx |
| Lung | 2,948 | 23,779 | 13.78 | TCGA, GTEx, CMB |
| Breast | 8,225 | 335,989 | 42.75 | TCGA, GTEx, ACROBAT, DROID-Breast, CMB, CAMELYON16, CAMELYON17, BRACS, TIGER2021, Post-NAT-BRCA |
| Esophagus | 3,094 | – | 14.07 | TCGA, GTEx, ESCA |
| Stomach | 1,609 | – | 11.09 | TCGA, GTEx, CMB |
| Colon & Rectum | 4,288 | 207,538 | 35.47 | TCGA, GTEx, PAIP, CMB, HunCRC, UniToPatho, DigestPath2019, NCT-CRC-HE-100K |
| Liver | 1,574 | – | 11.02 | TCGA, GTEx, PAIP |
| Pancreas | 1,654 | – | 11.18 | TCGA, GTEx, PAIP, CMB |
| Kidney | 2,337 | 52,713 | 22.55 | TCGA, GTEx, PAIP, DHMC |
| Bladder | 896 | – | 9.66 | TCGA, GTEx |
| Prostate | 8,646 | – | 25.22 | TCGA, GTEx, PAIP, Diagest, AGGC2022 |
| Testis | 1,137 | – | 10.14 | TCGA, GTEx |
| Uterus (Endometrium) | 1,447 | – | 10.77 | TCGA, GTEx |
| Uterus (Cervix) | 1,176 | – | 10.22 | TCGA, GTEx, TissueNet |
| Ovary | 1,674 | – | 11.22 | TCGA, GTEx, CMB, UBC-OCEAN |
| Skin | 4,354 | – | 16.60 | TCGA, GTEx, BCC, CMB |
| Soft Tissue (Sarcoma) | 828 | – | 9.52 | TCGA, GTEx |
| Blood (Lymphoma & Leukemia) | 926 | 783,108 | 19.72 | TCGA, GTEx, CMB |
| Thymus | 859 | – | 9.58 | TCGA, GTEx |
| Adrenal Gland | 1,472 | – | 10.82 | TCGA, GTEx |
| Other (Multi-organ) | 1,335 | 347,521 | 20.54 | TCGA, GTEx, CMB, DLBCL-Morph |
| **Total** | **55,648** | **1,750,648** | **360.79** | – |

**Supplementary Table 2. TEAM architecture specifications.** Component specifications and parameter counts for TEAM. The patch encoder uses DINOv2-L/16 initialized with UNI pretrained weights; uncertainty modeling uses a VAE and Monte Carlo dropout; the Clinical Metadata Encoder (TEAM-R) uses frozen BioBERT-base; and the interactive module combines an MLP projector with LoRA fine-tuning.

| Component | Specification | Parameters |
|---|---|---|
| *Patch Encoder* | | |
| Architecture | DINOv2-L/16 (ViT-Large) | 304M |
| Initialization | UNI pretrained weights | – |
| Fine-tuning strategy | Block 1 + Blocks 23–24 (others frozen) | ∼38M trainable |
| Input resolution | 224 × 224 pixels | – |
| Output dimension | 1024-dimensional embeddings | – |
| *Uncertainty Modeling* | | |
| VAE encoder | 6× MLP heads (1024 → 512 → 512) | 4.7M |
| VAE latent dim | 512-dimensional Gaussian | – |
| MC Dropout rate ($\pi$) | 0.3 | – |
| MC Dropout passes (T) | 3 | – |
| *Clinical Metadata Encoder (TEAM-R)* | | |
| Architecture | BioBERT-base | 110M (frozen) |
| Input | Free-text age, sex, race, and ethnicity (Supplementary Table 6) | – |
| Output dimension | 768 → projected to 1024 | 0.8M |
| *Aggregation Module* | | |
| Attention mechanism | Linear projection: 512 → 512 → 256 → 1, followed by softmax | 0.4M |
| Uncertainty modulation | Uncertainty-derived reweighting using normalized predictive variance and inter-head disagreement to modulate attention scores | – |
| Risk predictor ($\psi$) | MLP: 512 → 256 → 1 with tanh activation | 0.13M |
| *Interactive Module* | | |
| MLP projector (VLM) | 1024 → 4096 → 4096 + GELU | 21M |
| LoRA modules (LLM) | Rank 256, $\alpha$ 128, dropout 0.05 | ∼2B (5% of LLM) |
| **Total Parameters** | **TEAM-V (frozen) + trainable components** | **∼65M trainable** |

**Supplementary Table 3. TEAM pretraining configuration.** Hyperparameters and training configuration for TEAM pretraining. Training was performed on 5 × NVIDIA RTX 5880 Ada GPUs (48 GB each). The relative risk prior incorporates population-level survival statistics from China, Europe, and the United States. NCCR, National Central Cancer Registry of China.

| **Hyperparameter** | **Value** | **Notes** |
|---|---|---|
| *Dataset* | | |
| Training patches | 360,786,410 | After quality filtering |
| *Training Setup* | | |
| Batch size | 2048 patches | |
| Epochs | 100 | |
| Precision | Mixed (FP16/BF16) | |
| *Optimization* | | |
| Optimizer | AdamW | |
| Base learning rate | 1e-4 | With linear warmup |
| Warmup iterations | 10,000 | |
| LR scheduler | Cosine decay | Final LR = 1e-6 |
| Weight decay | 0.05 | Applied to non-bias params |
| Gradient clipping | 1.0 (global norm) | |
| *Loss Weights* | | |
| Self-distillation (DINO) | 1.0 | |
| Masked image modeling | 1.0 | |
| KL divergence (VAE) | 0.1 | Uncertainty regularization |
| Ranking objective weight ($\delta$) | 0.01 | Relative risk prior regularization |
| *Registry-specific prior* | | |
| Registries | China, Europe, United States | |
| Survival margins ($\tau_s$) | 15%, 20%, 5% respectively | Set by clinicians |
| ICD-10 categories | 33 cancer types | With subtype stratification |
| Data source | NCCR China, EUROCARE-5, SEER | Population-based registries |
| *Data Augmentation* | | |
| Random horizontal flip | p = 0.5 | |
| Random vertical flip | p = 0.5 | |
| Random rotation | ±90° with p = 0.3 | |
| Color jitter | Brightness ±0.2, contrast ±0.2 | |
| Stain normalization | Macenko method | Optional, not always used |

**Supplementary Table 4. Vision–language alignment training details.** Training comprised feature alignment using Quilt-1M image–caption pairs followed by instruction tuning using Quilt-Instruct pathology Q&A data.

| **Hyperparameter** | **Stage 1 (Feature Alignment)** | **Stage 2 (Instruction Tuning)** |
|---|---|---|
| *Dataset* | | |
| Training data | Quilt-1M (723K image-caption pairs) | Quilt-Instruct (107K Q&A pairs) |
| Data source | PubMed articles + TCGA | Pathology instruction dataset |
| *Model Configuration* | | |
| Vision encoder | TEAM-V (frozen) | TEAM-V (frozen) |
| MLP projector | 1024 → 4096 → 4096 + GELU (trainable) | Trainable |
| Language model | Frozen | LoRA fine-tuning |
| LoRA hyperparameters | – | rank 256, $\alpha$ 128, dropout 0.05 |
| LoRA target layers | – | Q, K, V, O, Gate, Up, Down |
| LoRA % parameters | – | ∼4–6% of LLM params |
| *Training Setup* | | |
| Batch size (global) | 64 | 64 |
| Number of GPUs | 5 × RTX 5880 Ada | 5 × RTX 5880 Ada |
| Epochs | 1 | 2 |
| Max sequence length | 2048 tokens | 2048 tokens |
| Precision | BF16 | BF16 |
| DeepSpeed | ZeRO-2 | ZeRO-3 |
| Wall-clock time | ∼8 hours | ∼6 hours |
| GPU-hours | ∼40 | ∼30 |
| *Optimization* | | |
| Optimizer | AdamW | AdamW |
| Learning rate | 1e-3 | 2e-4 |
| Weight decay | 0.0 | 0.0 |
| LR scheduler | Cosine decay | Cosine decay |
| Warmup ratio | 3% | 3% |
| Gradient clipping | 1.0 (global norm) | 1.0 (global norm) |
| Gradient accumulation | 1 step | 1 step |
| *Evaluation* | | |
| Validation metric | Image-text retrieval Recall@10 | Instruction-following accuracy |
| Validation frequency | Every 1,000 steps | Every 500 steps |
| Early stopping | Not used | Best checkpoint by val accuracy |

**Supplementary Table 5. Hyperparameters for ViT-L/16 pretraining with DINOv2.** Hyperparameters used for ViT-L/16 pretraining. Batch size refers to the total batch size across all GPUs.

| **Hyperparameter** | **Value** | **Hyperparameter** | **Value** |
|---|---|---|---|
| Layers | 24 | Global crop scale | 0.48, 1.0 |
| Heads | 16 | Global crop number & size | 2, 224 |
| Patch size | 16 | Local crop scale | 0.16, 0.48 |
| FFN layer | MLP | Local crop number & size | 8, 96 |
| Head activation | GELU | Max masking ratio | 0.5 |
| Embedding dimension | 1024 | Min masking ratio | 0.1 |
| Normalize last layer | ✓ | Stochastic dropout rate | 0.1 |
| Shared head | ✗ | Gradient clipping max norm | 3.0 |
| AdamW $\beta$ | (0.9, 0.999) | Learning rate (start) | 0 |
| Batch size | 3072 | Learning rate (post warmup) | 2e-3 |
| Freeze last layer iterations | 1250 | Learning rate (final) | 1e-6 |
| Warmup iterations | 12500 | Teacher temperature (start) | 0.04 |
| Warmup teacher temp. iterations | 37500 | Teacher temperature (final) | 0.4 |
| High-res finetuning iterations | 12500 | Teacher momentum (start) | 0.992 |
| Max iterations | 125000 | Teacher momentum (final) | 1.000 |
| Learning rate schedule | Cosine | Weight decay (start) | 0.04 |
| Weight decay (end) | 0.4 | Automatic mixed precision | FP16 |

**Supplementary Table 6. TEAM-R input variables and representative input examples.**

| Variable | Used as TEAM-R input |
|---|---|
| **Demographic variables (provided during pretraining only)** | |
| Age | ✓ |
| Sex | ✓ |
| Race | ✓ |
| Ethnicity | ✓ |
| **Organ and diagnosis** | |
| Organ | ✗ |
| Tissue type | ✗ |
| Cancer type | ✗ |
| Pathological diagnosis | ✗ |
| Histologic subtype | ✗ |
| **Outcome prediction** | |
| Overall survival (time and event) | ✗ |
| Therapeutic response | ✗ |
| **Biomarker profiling** | |
| Tumor stage | ✗ |
| Tumor grade | ✗ |
| TME biomarkers (PD-L1, CD8/CD4, TILs, Ki-67, TMB, MSI, HRD, hypoxia) | ✗ |
| Gene expression | ✗ |
| Molecular testing results | ✗ |
| **Other clinical records** | |
| Pathology report text | ✗ |
| Laboratory values | ✗ |

| Case | Age | Sex | Race | Ethnicity | TEAM-R input text (verbatim) |
|---|---|---|---|---|---|
| Case 1 | ✓ | ✓ | ✓ | ✓ | Age: 71–75 years. Sex: male. Race: white. Ethnicity: not Hispanic or Latino. |
| Case 2 | ✓ | ✓ | ✓ | – | Age: 61–65 years. Sex: male. Race: white. |
| Case 3 | ✓ | ✓ | – | – | Age: 66–70 years. Sex: female. |
| Case 4 | – | ✓ | – | – | Sex: male. |
| Case 5 | – | – | – | – | None |

**Supplementary Table 7. Availability of TEAM-R input variables during pre-training.** Only datasets with at least one of the four variables recorded are listed.

| Dataset | WSIs | WSIs with the variable available | | | | WSIs by number of missing variables | | | | |
|---|---|---|---|---|---|---|---|---|---|---|
| | | Age | Sex | Race | Ethnicity | 0 | 1 | 2 | 3 | 4 |
| GTEx | 25,713 | 25,713 | 25,713 | 0 | 0 | 0 | 0 | 25,713 | 0 | 0 |
| TCGA | 7,784 | 7,710 | 7,758 | 7,048 | 6,090 | 6,014 | 1,099 | 608 | 37 | 26 |
| ACROBAT | 4,212 | 0 | 4,212 | 0 | 0 | 0 | 0 | 0 | 4,212 | 0 |
| CMB | 2,141 | 2,137 | 2,137 | 2,061 | 2,094 | 2,027 | 101 | 9 | 0 | 4 |
| BCNB | 1,058 | 1,058 | 0 | 0 | 0 | 0 | 0 | 0 | 1,058 | 0 |
| TIGER2021 | 370 | 151 | 151 | 144 | 132 | 131 | 14 | 6 | 0 | 219 |
| DROID-Breast | 361 | 0 | 296 | 0 | 0 | 0 | 0 | 0 | 296 | 65 |
| DLBCL-Morph | 246 | 246 | 0 | 0 | 0 | 0 | 0 | 0 | 246 | 0 |
| HunCRC | 200 | 200 | 200 | 0 | 0 | 0 | 0 | 200 | 0 | 0 |
| Post-NAT-BRCA | 96 | 96 | 0 | 0 | 0 | 0 | 0 | 0 | 96 | 0 |
| **Overall** | **42,181** | **37,311** | **40,467** | **9,253** | **8,316** | **8,172** | **1,214** | **26,536** | **5,945** | **314** |

**Supplementary Table 8. Cancer subtype risk rankings.** Risk rankings of cancer subtypes within each cancer type. Rank 1 indicates the highest-risk subtype. Covers 55 subtypes across 20 cancer types, listed in alphabetical order by abbreviation. Cancer types are given as TCGA study abbreviations.

| Cancer | Subtype | Rank |
|---|---|---|
| ACC | Adrenal cortical carcinoma | 1 |
| | Paraganglioma | 2 |
| | Pheochromocytoma, NOS | 3 |
| BRCA | Infiltrating duct and lobular carcinoma | 1 |
| | Infiltrating duct carcinoma | 2 |
| | Lobular carcinoma | 3 |
| CESC | Adenosquamous carcinoma | 1 |
| | Adenocarcinoma | 2 |
| | Squamous cell carcinoma | 3 |
| COAD | Mucinous adenocarcinoma | 1 |
| | Adenocarcinoma, NOS | 2 |
| ESCA | Adenocarcinoma | 1 |
| | Squamous cell carcinoma | 2 |
| KICH | Renal clear cell carcinoma | 1 |
| | Renal chromophobe carcinoma | 2 |
| | Renal papillary cell carcinoma | 3 |
| LGG | Glioblastoma | 1 |
| | Mixed glioma | 2 |
| | Astrocytoma | 3 |

*Continued on next page*

| Cancer | Subtype | Rank |
|---|---|---|
| | Oligodendroglioma | 4 |
| LIHC | Combined hepatocellular carcinoma and cholangiocarcinoma | 1 |
| | Cholangiocarcinoma | 2 |
| | Hepatocellular carcinoma, NOS | 3 |
| LUAD | Lung adenocarcinoma | 1 |
| | Lung squamous cell carcinoma | 2 |
| PAAD | Neuroendocrine carcinoma | 1 |
| | Infiltrating duct carcinoma, NOS | 2 |
| | Adenocarcinoma, NOS | 3 |
| READ | Mucinous adenocarcinoma | 1 |
| | Adenocarcinoma | 2 |
| SARC | Undifferentiated sarcoma | 1 |
| | Fibromyxosarcoma | 2 |
| | Synovial sarcoma | 3 |
| | Leiomyosarcoma | 4 |
| | Liposarcoma | 5 |
| SKCM | Invasive melanoma | 1 |
| | Non-invasive or specific subtype melanoma | 2 |
| STAD | Carcinoma, diffuse type | 1 |
| | Adenocarcinoma | 2 |
| TGCT | Non-seminoma | 1 |
| | Seminoma, NOS | 2 |
| THCA | Papillary carcinoma, follicular variant | 1 |
| | Papillary adenocarcinoma, NOS | 2 |
| THYM | Thymoma, type B3, malignant | 1 |
| | Thymoma, type B2, malignant | 2 |
| | Thymoma, type B1, malignant | 3 |
| | Thymoma, type AB, malignant | 4 |
| | Thymoma, type A, malignant | 5 |
| UCEC | Serous cystadenocarcinoma | 1 |
| | Endometrioid adenocarcinoma | 2 |
| UCS | Carcinosarcoma, NOS | 1 |
| | Müllerian mixed tumor | 2 |
| UVM | Mixed epithelioid and spindle cell melanoma | 1 |
| | Epithelioid cell melanoma | 2 |
| | Spindle cell melanoma | 3 |

**Supplementary Table 9. In-house validation cohorts.** Thirteen institutional cohorts spanning six cancer types were assembled for downstream evaluation. Cohort abbreviations and institutional sources are listed below.

| **Cancer Type** | **Institution** | **Abbreviation** |
|---|---|---|
| Colorectal | The Fourth Hospital of Hebei Medical University | HBFH-CRC |
| | Anhui Provincial Hospital | APH-CRC |
| | The Sixth Affiliated Hospital of Sun Yat-sen University | SYSH-CRC |
| | Guangdong Provincial People's Hospital | GDPH-CRC |
| | Nanfang Hospital | NFH-CRC |
| Breast | Nanfang Hospital | NFH-BRCA |
| | Qingdao Center Hospital | QDUH-BRCA |
| | Shandong University Qilu Hospital | SUQH-BRCA |
| | The Second Hospital of Shandong University | SHSU-BRCA |
| Liver | Zhujiang Hospital | ZJH-LIHC |
| Head and Neck | The Affiliated Hospital of Qingdao University | QDPH-HNSC |
| Bladder | The Affiliated Hospital of Qingdao University | QDPH-BLCA |
| Cervical | Nanfang Hospital | NFH-CESC |

**Supplementary Table 10. Organ-system distribution of downstream evaluation data.** WSI and ROI counts across 23 organ-system categories in the downstream datasets, comprising a total of 30,313 WSIs and 848,655 ROIs. WSI counts are the union of slides used across the downstream tasks; per-task counts are given in Supplementary Tables 11–19.

| Organ | WSI Count | ROI Count |
|---|---|---|
| Brain | 1,251 | 12,679 |
| Head & Neck | 1,016 | 17,296 |
| Thyroid | – | 13,220 |
| Lung | 2,774 | 44,561 |
| Breast | 5,828 | 33,384 |
| Esophagus | – | 6,282 |
| Stomach | 360 | 12,227 |
| Colon & Rectum | 1,481 | 19,527 |
| Liver | 453 | 25,187 |
| Pancreas | 627 | 12,871 |
| Kidney | 1,712 | 29,584 |
| Bladder | 898 | 11,219 |
| Prostate | 11,314 | 9,206 |
| Testis | – | 7,543 |
| Uterus (Endometrium) | 1,274 | 11,739 |
| Uterus (Cervix) | 648 | 8,264 |
| Ovary | 677 | 16,900 |
| Skin | – | 99,602 |
| Soft Tissue (Sarcoma) | – | 45,270 |
| Blood (Lymphoma & Leukemia) | – | 41,498 |
| Thymus | – | 1,543 |
| Adrenal Gland | – | 3,594 |
| Other (Multi-organ) | – | 365,459 |
| **Total** | **30,313** | **848,655** |

**Supplementary Table 11. Distribution of prognostic stratification datasets.** Patient and slide counts for training and test splits across cancer types. Gray-shaded rows indicate datasets used for training and internal testing.

| Dataset | Patients/Slides | Train Patients/Slides | Test Patients/Slides |
|---|---|---|---|
| TCGA-BLCA | 399/457 | 283/316 | 116/141 |
| QDPH-BLCA | 441/441 | – | 441/441 |
| TCGA-BRCA | 1,007/1,129 | 690/791 | 317/338 |
| NFH-BRCA | 225/225 | – | 225/225 |
| TCGA-CESC | 272/279 | 191/197 | 81/82 |
| NFH-CESC | 192/405 | – | 192/405 |
| TCGA-CRC | 457/623 | 273/436 | 184/187 |
| HBFH-CRC | 49/49 | – | 49/49 |
| GDPH-CRC | 82/82 | – | 82/82 |
| APH-CRC | 118/118 | – | 118/118 |
| NFH-CRC | 145/458 | – | 145/458 |
| SYSH-CRC | 34/34 | – | 34/34 |
| TCGA-GBM | 349/811 | 231/533 | 118/278 |
| CPTAC-GBM | 160/373 | – | 160/373 |
| TCGA-HNSC | 450/471 | 315/334 | 135/137 |
| QDPH-HNSC | 201/201 | – | 201/201 |
| CPTAC-HNSC | 101/308 | – | 101/308 |
| TCGA-KIRC | 352/518 | 198/364 | 154/154 |
| CPTAC-KIRC | 196/608 | – | 196/608 |
| TCGA-LIHC | 348/378 | 239/266 | 109/112 |
| ZJH-LIHC | 75/75 | – | 75/75 |
| TCGA-LUAD | 502/531 | 365/366 | 137/165 |
| CPTAC-LUAD | 202/909 | – | 202/909 |
| TCGA-nCCRCC | 336/409 | 223/289 | 113/120 |
| CPTAC-nCCRCC | 39/89 | – | 39/89 |
| TCGA-UCEC | 507/547 | 360/380 | 147/167 |
| CPTAC-UCEC | 220/727 | – | 220/727 |
| TCGA-LUSC | 478/504 | 336/351 | 142/153 |
| CPTAC-LUSC | 102/435 | – | 102/435 |
| TCGA-PAAD | 183/209 | 128/153 | 55/56 |
| CPTAC-PAAD | 139/418 | – | 139/418 |
| **Total** | **8,361 / 12,821** | **3,832 / 4,776** | **4,529 / 8,045** |

**Supplementary Table 12. Distribution of patient clinical metadata.** Patient demographics including age and sex. M: male; F: female.

| Dataset | Age (mean ± s.d.) | Sex (M/F) |
|---|---|---|
| TCGA-BLCA | 68.2 ± 10.6 | 295/104 |
| QDPH-BLCA | 67.1 ± 11.2 | 362/79 |
| TCGA-BRCA | 58.4 ± 13.2 | 0/1,007 |
| NFH-BRCA | 57.3 ± 14.3 | 0/225 |
| TCGA-CESC | 48.2 ± 13.8 | 0/272 |
| NFH-CESC | 51.7 ± 11.3 | 0/192 |
| TCGA-CRC | 65.4 ± 13.1 | 232/225 |
| HBFH-CRC | 63.5 ± 14.3 | 30/19 |
| GDPH-CRC | 66.5 ± 12.5 | 49/33 |
| APH-CRC | 63.4 ± 11.5 | 79/39 |
| NFH-CRC | 65.5 ± 12.5 | 93/52 |
| SYSH-CRC | 63.4 ± 11.2 | 12/22 |
| TCGA-GBM | 57.3 ± 14.5 | 223/126 |
| CPTAC-GBM | 58.7 ± 13.6 | 98/62 |
| TCGA-HNSC | 61.1 ± 12.0 | 328/122 |
| QDPH-HNSC | 66.4 ± 13.4 | 144/57 |
| CPTAC-HNSC | 61.8 ± 8.9 | 88/13 |
| TCGA-KIRC | 60.1 ± 11.9 | 219/133 |
| CPTAC-KIRC | 60.7 ± 11.8 | 140/56 |
| TCGA-LIHC | 59.3 ± 13.3 | 234/114 |
| ZJH-LIHC | 56.9 ± 9.1 | 60/15 |
| TCGA-LUAD | 65.3 ± 10.0 | 233/269 |
| CPTAC-LUAD | 63.5 ± 9.4 | 129/73 |
| TCGA-nCCRCC | 59.3 ± 13.1 | 240/96 |
| CPTAC-nCCRCC | 64.4 ± 9.0 | 23/16 |
| TCGA-UCEC | 63.8 ± 11.1 | 0/507 |
| CPTAC-UCEC | 64.6 ± 9.5 | 0/220 |
| TCGA-LUSC | 67.3 ± 8.6 | 357/121 |
| CPTAC-LUSC | 66.4 ± 8.4 | 83/19 |
| TCGA-PAAD | 64.9 ± 11.1 | 102/81 |
| CPTAC-PAAD | 64.4 ± 11.0 | 71/68 |

**Supplementary Table 13. Distribution of therapeutic response datasets.** Counts are shown as total (responders/non-responders). BEV-OV: bevacizumab-treated ovarian cancer; PDL1-NSCLC: PD-L1 blockade in non-small cell lung cancer; CRT-CESC: chemoradiotherapy in cervical squamous cell carcinoma. Yale HER2+, IMPRESS HER2+, and TransNEO HER2+: neoadjuvant therapy in HER2+ breast cancer, with pathological complete response counted as response.

| **Dataset** | **Yale HER2+** | **IMPRESS HER2+** | **TransNEO HER2+** | **BEV-OV** | **PDL1-NSCLC** | **CRT-CESC** |
|---|---|---|---|---|---|---|
| Training | 59 (25/34) | – | – | 200 (112/88) | 135 (101/34) | 123 (87/36) |
| Testing | 26 (11/15) | 62 (38/24) | 76 (30/46) | 86 (48/38) | 58 (44/14) | 54 (38/16) |
| **Total** | **85 (36/49)** | **62 (38/24)** | **76 (30/46)** | **286 (160/126)** | **193 (145/48)** | **177 (125/52)** |

**Supplementary Table 14. Distribution of cancer staging datasets.** Sample counts (whole-slide images) across different cancer stages (I–IV) for training and testing datasets. Numbers in parentheses indicate train/test split where available. HGSOC-OV, high-grade serous ovarian carcinoma.

| **Dataset** | **I** | **II** | **III** | **IV** | **Total (Train/Test)** |
|---|---|---|---|---|---|
| TCGA-BRCA | 180 (71/109) | 644 (458/186) | 262 (111/151) | 32 (12/20) | 1,118 (652/466) |
| CPTAC-BRCA | 9 | 80 | 39 | 0 | 128 |
| NFH-BRCA | 72 | 115 | 13 | 14 | 214 |
| TCGA-CESC | 140 (103/37) | 61 (34/27) | 43 (26/17) | 21 (11/10) | 265 (174/91) |
| NFH-CESC | 50 | 101 | 22 | 19 | 192 |
| TCGA-CRC | 106 (57/49) | 222 (156/66) | 176 (110/66) | 87 (39/48) | 591 (362/229) |
| CPTAC-CRC | 13 | 44 | 46 | 7 | 110 |
| NFH-CRC | 0 | 339 | 126 | 0 | 465 |
| TCGA-HNSC | 53 (28/25) | 71 (25/46) | 73 (21/52) | 249 (176/73) | 446 (250/196) |
| CPTAC-HNSC | 31 | 91 | 104 | 104 | 330 |
| TCGA-LIHC | 84 (60/24) | 74 (52/22) | 71 (49/22) | 10 (5/5) | 239 (166/73) |
| ZJH-LIHC | 37 | 16 | 6 | 14 | 73 |
| TCGA-OV | 2 (1/1) | 8 (4/4) | 22 (14/8) | 13 (7/6) | 45 (26/19) |
| HGSOC-OV | 2 | 4 | 294 | 46 | 346 |
| **Total** | **779** | **1,870** | **1,297** | **616** | **4,562** |

**Supplementary Table 15. Distribution of cancer grading datasets.** Sample counts (whole-slide images) across histological grading categories for training and testing datasets. Numbers in parentheses indicate train/test splits where available.

| Dataset | G1 | G2 | G3 | G4 | G5 | Total (Train/Test) |
|---|---|---|---|---|---|---|
| SUQH-BRCA | 602 (522/80) | 1,717 (1,156/561) | 1,130 (848/282) | 0 | 0 | 3,449 (2,526/923) |
| QDUH-BRCA | 80 | 169 | 141 | 0 | 0 | 390 |
| SHSU-BRCA | 43 | 128 | 113 | 0 | 0 | 284 |
| TCGA-PRAD | 74 (42/32) | 209 (146/63) | 107 (61/46) | 143 (96/47) | 10 (5/5) | 543 (350/193) |
| PANDA-PRAD | 5,558 | 1,343 | 1,242 | 1,249 | 1,224 | 10,616 |
| SICAP-PRAD | 50 | 45 | 18 | 35 | 7 | 155 |
| **Total** | **6,407** | **3,611** | **2,751** | **1,427** | **1,241** | **15,437** |

**Supplementary Table 16. Distribution of tumor microenvironment (TME) datasets.** Sample counts (whole-slide images) for TME characterization across six cancer types from TCGA and CPTAC datasets. Gray-shaded rows indicate TCGA datasets used for training and internal testing.

| Dataset | Train | Test | Total |
|---|---|---|---|
| TCGA-BLCA | 289 | 110 | 399 |
| TCGA-BRCA | 810 | 319 | 1,129 |
| CPTAC-BRCA | – | 114 | 114 |
| TCGA-CESC | 196 | 83 | 279 |
| TCGA-CRC | 443 | 180 | 623 |
| CPTAC-CRC | – | 108 | 108 |
| TCGA-HNSC | 345 | 126 | 471 |
| CPTAC-HNSC | – | 326 | 326 |
| TCGA-LIHC | 267 | 111 | 378 |
| **Total** | **2,350** | **1,477** | **3,827** |

**Supplementary Table 17. Distribution of immune surveillance profile samples.** Sample counts for immune surveillance biomarkers including programmed death-ligand 1 (PD-L1), intratumoral CD8+/CD4+ T-cell ratio, tumor-infiltrating lymphocyte density (TILs), and Ki-67 proliferation index.

| Dataset | PD-L1 | CD8/CD4 | TILs | Ki-67 |
|---|---|---|---|---|
| TCGA-BLCA | 108 | 106 | 74 | 107 |
| TCGA-BRCA | 310 | 281 | 250 | 313 |
| CPTAC-BRCA | 105 | 105 | 105 | 105 |
| TCGA-CESC | 82 | 76 | 66 | 82 |
| TCGA-CRC | 177 | 162 | 158 | 175 |
| CPTAC-CRC | 104 | 104 | 104 | 104 |
| TCGA-HNSC | 126 | 0 | 0 | 0 |
| CPTAC-HNSC | 326 | 326 | 326 | 326 |
| TCGA-LIHC | 107 | 99 | 0 | 110 |
| **Total** | **1,445** | **1,259** | **1,083** | **1,322** |

**Supplementary Table 18. Distribution of TME stress profile samples.** Sample counts for TME stress biomarkers including tumor mutational burden (TMB), microsatellite instability (MSI), homologous recombination deficiency (HRD), and Winter hypoxia metagene.

| Dataset | TMB | MSI | HRD | Hypoxia |
|---|---|---|---|---|
| TCGA-BLCA | 101 | 105 | 107 | 110 |
| TCGA-BRCA | 252 | 291 | 271 | 312 |
| CPTAC-BRCA | 99 | 99 | 0 | 105 |
| TCGA-CESC | 72 | 78 | 73 | 78 |
| TCGA-CRC | 143 | 140 | 133 | 100 |
| CPTAC-CRC | 103 | 103 | 0 | 104 |
| TCGA-HNSC | 114 | 0 | 0 | 0 |
| CPTAC-HNSC | 326 | 326 | 326 | 326 |
| TCGA-LIHC | 106 | 99 | 105 | 106 |
| **Total** | **1,316** | **1,241** | **1,015** | **1,241** |

**Supplementary Table 19. Distribution of gene expression datasets.** Sample counts (whole-slide images) for gene expression prediction across 11 cancer types from TCGA and CPTAC datasets. Gray-shaded rows indicate TCGA datasets used for training and internal testing.

| Dataset | Train | Test | Total |
|---|---|---|---|
| TCGA-BLCA | 291 | 125 | 416 |
| TCGA-BRCA | 773 | 356 | 1,129 |
| CPTAC-BRCA | – | 96 | 96 |
| TCGA-CESC | 187 | 92 | 279 |
| TCGA-CRC | 420 | 180 | 600 |
| CPTAC-CRC | – | 100 | 100 |
| TCGA-GBM | 65 | 28 | 93 |
| CPTAC-GBM | – | 440 | 440 |
| TCGA-HNSC | 323 | 148 | 471 |
| CPTAC-HNSC | – | 313 | 313 |
| TCGA-KIRC | 353 | 165 | 518 |
| CPTAC-KIRC | – | 696 | 696 |
| TCGA-LIHC | 251 | 127 | 378 |
| TCGA-LUAD | 338 | 145 | 483 |
| CPTAC-LUAD | – | 1,063 | 1,063 |
| TCGA-PRAD | 303 | 130 | 433 |
| TCGA-STAD | 252 | 108 | 360 |
| **Total** | **3,556** | **4,312** | **7,868** |

**Supplementary Table 20. Distribution of question-answering model training datasets.** Two-stage training approach for question-answering model development. Stage 1 focuses on projection learning, while Stage 2 performs instruction-based fine-tuning.

| Attribute | Stage 1 | Stage 2 |
|---|---|---|
| Dataset | Quilt-1M | Quilt-Instruct |
| Data Volume | 723,328 | 107,131 |
| Purpose | Projection | Fine-tuning |

**Supplementary Table 21. Distribution of question-answering model testing datasets.** Comprehensive evaluation across multiple pathology-specific Q&A datasets. PathMMU includes both educational and literature-based subsets. QuiltVQA evaluation is restricted to closed-ended questions only. PathQABench represents open-ended evaluation.

| Test Dataset | Data Volume | Description |
|---|---|---|
| | *Closed-ended Evaluation* | |
| PathMMU-EduContent | 255 | Educational content subset |
| PathMMU-PubMed | 281 | PubMed literature subset |
| QuiltVQA | 343 | Closed-ended visual Q&A |
| PathVQA | 3,391 | Closed-ended (yes/no) visual Q&A |
| **Closed-ended Total** | **4,270** | |
| | *Open-ended Evaluation* | |
| PathQABench | 260 | Pathology Q&A benchmark |
| **Overall Total** | **4,530** | |

**Supplementary Table 22. Data availability and links to the datasets.** Pathology and multimodal datasets used in this study, with links to their repositories, project pages, or access instructions.

| Dataset | Link |
|---|---|
| *Upstream and downstream datasets* | |
| TCGA | `https://portal.gdc.cancer.gov` |
| *Upstream pretraining datasets* | |
| ACROBAT | `https://doi.org/10.48723/w728-p041` |
| AGGC2022 | `https://aggc22.grand-challenge.org/Data/` |
| ANHIR | `https://anhir.grand-challenge.org/` |
| BACH | `https://zenodo.org/records/3632035` |
| BCC | `https://datahub.aida.scilifelab.se/10.23698/aida/bccc` |
| BCNB | `https://bcnb.grand-challenge.org/` |
| BRACS | `https://www.bracs.icar.cnr.it/download/` |
| BreakHis | `https://web.inf.ufpr.br/vri/databases/breast-cancer-histopathological-database-BreakHis/` |
| BreastPathQ | `https://breastpathq.grand-challenge.org/` |
| CAMELYON16 | `https://camelyon16.grand-challenge.org/Data/` |
| CAMELYON17 | `https://camelyon17.grand-challenge.org/Data/` |

*Continued on next page*

| Dataset | Link |
|---|---|
| Camelyon17-WILDS | https://wilds.stanford.edu/datasets/ |
| CCRCC-TCGA-HEL | https://zenodo.org/records/7898308 |
| Chaoyang | https://bupt-ai-cz.github.io/HSA-NRL/ |
| CMB | https://pathdb.cancerimagingarchive.net/index.php/collections |
| CoCaHis | https://cocahis.irb.hr/ |
| CoNIC2022 | https://conic-challenge.grand-challenge.org/ |
| CRC-MSI | https://zenodo.org/records/3832231 |
| Dataset-PT | https://github.com/CSU-BME/pathology_SSL |
| DHMC | https://bmirds.github.io/KidneyCancer/ |
| DiagSet | https://github.com/michalkoziarski/DiagSet |
| DiagSet-B | https://github.com/michalkoziarski/DiagSet |
| DigestPath2019 (colonoscopy tissue segmentation and classification) | https://medicalimaging.ai/challenges/digestpath2019 |
| DigestPath2019 (signet-ring cell detection) | https://medicalimaging.ai/challenges/digestpath2019 |
| DLBCL-Morph | https://github.com/stanfordmlgroup/DLBCL-Morph |
| DROID-Breast | https://datahub.aida.scilifelab.se/10.23698/aida/drbr |
| EBHI-Seg | https://doi.org/10.6084/m9.figshare.21540159 |
| ESCA | https://zenodo.org/records/7548828 |
| FocusPath | https://github.com/mahdihosseini/FoucsPath |
| GlaS | https://academictorrents.com/details/208814dd113c2b0a242e74e83 2ccac28fcff74e5 |
| Gleason2019 | https://gleason2019.grand-challenge.org/Register/ |
| GTEx | https://www.gtexportal.org/home/ |
| HunCRC | https://www.cancerimagingarchive.net/collection/hungarian-col orectal-screening/ |
| ICDC-Glioma | https://portal.imaging.datacommons.cancer.gov/collections/icdc _glioma/ |
| KPIs | https://sites.google.com/view/kpis2024 |
| LC25000 | https://github.com/tampapath/lung_colon_image_set |
| Malignant Lymphoma | https://www.kaggle.com/datasets/andrewmvd/malignant-lymphoma-c lassification |
| MedFM2023 (ColonPath) | https://github.com/openmedlab/dataset/blob/main/MedFM.md |
| MHIST | https://bmirds.github.io/MHIST/ |
| MIDOG2021 | https://midog.deepmicroscopy.org/2021/getting-the-data |
| MIDOG2022 | https://midog.deepmicroscopy.org/2022/download-dataset |
| MIDOG++ | https://doi.org/10.6084/m9.figshare.c.6615571.v1 |
| MSK MIND (NSCLC) | https://www.synapse.org/Synapse:syn26642505/datasets/ |
| NCT-CRC-HE-100K | https://zenodo.org/records/1214456 |
| NADT-Prostate | https://www.cancerimagingarchive.net/collection/nadt-prostate/ |
| NLST | https://www.cancerimagingarchive.net/collection/nlst/ |
| NuInsSeg | https://zenodo.org/records/10518968 |
| Osteosarcoma | https://www.cancerimagingarchive.net/collection/osteosarcoma-t umor-assessment/ |

*Continued on next page*

| Dataset | Link |
|---|---|
| PAIP | https://wisepaip.org/ |
| PAIP2019 | https://paip2019.grand-challenge.org/ |
| PAIP2020 | https://paip2020.grand-challenge.org/ |
| PAIP2021 | https://paip2021.grand-challenge.org/ |
| PanCancer-TIL | https://zenodo.org/records/6604094 |
| PanNuke | https://warwick.ac.uk/fac/cross_fac/tia/data/pannuke/ |
| PatchCamelyon | https://github.com/basveeling/pcam |
| Post-NAT-BRCA | https://www.cancerimagingarchive.net/collection/post-nat-brca/ |
| SegPath | https://dakomura.github.io/SegPath/ |
| SLN-Breast | https://www.cancerimagingarchive.net/collection/sln-breast/ |
| TIGER2021 | https://tiger.grand-challenge.org/ |
| TissueNet | https://www.drivendata.org/competitions/67/competition-cervical-biopsy/ |
| TUPAC | https://tupac.grand-challenge.org/ |
| UBC-OCEAN | https://www.kaggle.com/competitions/UBC-OCEAN/data |
| UniToPatho | https://zenodo.org/record/4643645 |
| WSSS4LUAD | https://wsss4luad.grand-challenge.org/ |
| *Downstream evaluation datasets* | |
| BEV-OV | https://doi.org/10.7937/TCIA.985G-EY35 |
| CPTAC | https://proteomic.datacommons.cancer.gov/pdc/ |
| HGSOC | https://www.cancerimagingarchive.net/collection/ptrc-hgsoc/ |
| IMPRESS | https://tinyurl.com/IMPRESS-DATA |
| PANDA | https://www.kaggle.com/c/prostate-cancer-grade-assessment/data |
| PathMMU | https://pathmmu-benchmark.github.io/ |
| PathVQA | https://pathvqachallenge.grand-challenge.org/Data_Info/ |
| PDL1-NSCLC (IGNITE) | https://zenodo.org/records/17735903 |
| QuiltVQA | https://huggingface.co/datasets/wisdomik/Quilt_VQA |
| SICAPv2 | https://data.mendeley.com/datasets/9xxm58dvs3/1 |
| TransNEO | https://zenodo.org/records/6337925 |
| Yale HER2 | https://doi.org/10.7937/E65C-AM96 |

**Supplementary Table 23. Task-specific implementation of the compared pathology models.** ABMIL, attention-based multiple-instance learning.

| Model | Model type | Prognostic stratification | Therapeutic response prediction | Cancer staging | Cancer grading | TME characterization | Gene expression prediction |
|---|---|---|---|---|---|---|---|
| **UNI** | Patch-level | UNI patch encoder → ABMIL → survival prediction head | UNI patch encoder → ABMIL → classification head | UNI patch encoder → ABMIL → classification head | UNI patch encoder → ABMIL → multiclass classification head | UNI patch encoder → ABMIL → regression head | UNI patch encoder → ABMIL → multi-output regression head |
| **Prov-GigaPath** | Slide-level | Prov-GigaPath slide model → survival prediction head | Prov-GigaPath slide model → classification head | Prov-GigaPath slide model → classification head | Prov-GigaPath slide model → classification head | Prov-GigaPath slide model → regression head | Prov-GigaPath slide model → multi-output regression head |
| **CHIEF** | Slide-level | CHIEF WSI model → regression head → single-neuron mortality risk score | CHIEF slide model → classification head | CHIEF slide model → classification head | CHIEF slide model → classification head | CHIEF slide model → regression head | CHIEF slide model → multi-output regression head |
| **TITAN** | Slide-level | TITAN slide model → slide embedding → linear Cox proportional hazards model | TITAN slide model → classification head | TITAN slide model → slide embedding → logistic-regression linear probe | TITAN slide model → slide embedding → logistic-regression linear probe | TITAN slide model → regression head | TITAN slide model → multi-output regression head |
| **GPFM** | Patch-level | GPFM patch encoder → ABMIL → 4-bin discrete-time survival head → negative log-likelihood (NLL) survival loss | GPFM patch encoder → ABMIL → classification head | GPFM patch encoder → ABMIL → classification head | GPFM patch encoder → ABMIL → classification head | GPFM patch encoder → ABMIL → regression head | GPFM patch encoder → ABMIL → multi-output regression head |
| **DeepPT** | Slide-level | – | – | – | – | – | DeepPT slide representation → common multi-output regression head |
| **TEAM** | Slide-level | TEAM-V patch encoder → uncertainty-weighted aggregation → Cox risk head | TEAM-V patch encoder → uncertainty-weighted aggregation → classification head | TEAM-V patch encoder → uncertainty-weighted aggregation → classification head | TEAM-V patch encoder → uncertainty-weighted aggregation → classification head | TEAM-V patch encoder → uncertainty-weighted aggregation → regression head | TEAM-V patch encoder → uncertainty-weighted aggregation → multi-output regression head |

**Supplementary Table 24. Implementation settings of the compared pathology models.**

| Model | Checkpoint and source | Input magnification and patch size | Feature dimension |
|---|---|---|---|
| UNI | UNI (ViT-L/16), Hugging Face `MahmoodLab/UNI` | Native magnification (20× or 40×); 512 × 512-pixel patches, resized to 224 × 224 | 1024 (patch); slide-level via ABMIL |
| Prov-GigaPath | Tile and slide encoders, Hugging Face `prov-gigapath/prov-gigapath` | Native magnification (20× or 40×); 512 × 512-pixel patches, resized to 224 × 224 | 1536 (tile); 768 (slide) |
| CHIEF | CHIEF patch encoder (CTransPath backbone) and WSI aggregator, GitHub `hms-dbmi/CHIEF` | Native magnification (20× or 40×); 512 × 512-pixel patches, resized to 224 × 224 | 768 (patch, CTransPath); 768 (slide) |
| TITAN | TITAN slide encoder with CONCH v1.5 patch encoder, Hugging Face `MahmoodLab/TITAN` | Native magnification (20× or 40×); 512 × 512-pixel patches, resized to 224 × 224 | 768 (patch, CONCH v1.5); 768 (slide) |
| GPFM | GPFM (ViT-L/14), GitHub release `birkhoffkiki/GPFM` | Native magnification (20× or 40×); 512 × 512-pixel patches, resized to 224 × 224 | 1024 (patch); slide-level via ABMIL |
| DeepPT | DeepPT framework v1.1 (Zenodo, doi:10.5281/zenodo.11125591) with ImageNet-pretrained ResNet-50, trained on the study cohorts | Native magnification (20× or 40×); 512 × 512-pixel patches, resized to 224 × 224 | 2048 (tile, ResNet-50), compressed to 512 by an autoencoder |
| TEAM | TEAM-V checkpoint initialized from UNI pretrained weights, GitHub `juruoxcj/TEAM` | Native magnification (20× or 40×); 512 × 512-pixel patches, resized to 224 × 224 | 1024 (patch); 512 (slide) |

**Supplementary Table 25. Availability of TEAM-R variables for the Meta-only evaluation.** For each cohort, WSIs are the union of the WSIs used in any of the five downstream tasks; Tasks gives these tasks by Extended Data Fig. 8 panel (a, prognostic stratification; b, therapeutic response prediction; c, cancer staging/grading; d, TME characterization; e, gene expression prediction).

| Cohort | Tasks | WSIs | WSIs with the variable available | | | | WSIs by number of missing variables | | | | |
|---|---|---|---|---|---|---|---|---|---|---|---|
| | | | Age | Sex | Race | Ethnicity | 0 | 1 | 2 | 3 | 4 |
| **BLCA** | | | | | | | | | | | |
| TCGA-BLCA | a, d, e | 457 | 412 | 457 | 394 | 380 | 374 | 26 | 12 | 45 | 0 |
| QDPH-BLCA | a | 441 | 441 | 441 | 0 | 0 | 0 | 0 | 441 | 0 | 0 |
| **BRCA** | | | | | | | | | | | |
| TCGA-BRCA | a, c, d, e | 1,129 | 1,097 | 1,128 | 1,002 | 923 | 919 | 87 | 91 | 31 | 1 |
| NFH-BRCA | a, c | 225 | 225 | 225 | 0 | 0 | 0 | 0 | 225 | 0 | 0 |
| CPTAC-BRCA | c, d, e | 128 | 127 | 126 | 121 | 120 | 115 | 9 | 3 | 1 | 0 |
| SUQH-BRCA | c | 3,449 | 0 | 3,449 | 0 | 0 | 0 | 0 | 0 | 3,449 | 0 |
| QDUH-BRCA | c | 390 | 0 | 390 | 0 | 0 | 0 | 0 | 0 | 390 | 0 |
| SHSU-BRCA | c | 284 | 0 | 284 | 0 | 0 | 0 | 0 | 0 | 284 | 0 |
| Yale HER2+ | b | 85 | 0 | 0 | 0 | 0 | 0 | 0 | 0 | 0 | 85 |
| IMPRESS HER2+ | b | 62 | 62 | 62 | 0 | 0 | 0 | 0 | 62 | 0 | 0 |
| TransNEO HER2+ | b | 76 | 76 | 76 | 0 | 0 | 0 | 0 | 76 | 0 | 0 |
| **CESC** | | | | | | | | | | | |
| TCGA-CESC | a, c, d, e | 279 | 279 | 279 | 243 | 167 | 164 | 82 | 33 | 0 | 0 |

*Continued on next page*

| Cohort | Tasks | WSIs | WSIs with the variable available | | | | WSIs by number of missing variables | | | | |
|---|---|---|---|---|---|---|---|---|---|---|---|
| | | | Age | Sex | Race | Ethnicity | 0 | 1 | 2 | 3 | 4 |
| NFH-CESC | a, c | 192 | 192 | 192 | 0 | 0 | 0 | 0 | 192 | 0 | 0 |
| CRT-CESC | b | 177 | 177 | 177 | 0 | 0 | 0 | 0 | 177 | 0 | 0 |
| **CRC** | | | | | | | | | | | |
| TCGA-CRC | a, c, d, e | 623 | 619 | 619 | 364 | 349 | 348 | 17 | 254 | 0 | 4 |
| HBFH-CRC | a | 49 | 49 | 49 | 0 | 0 | 0 | 0 | 49 | 0 | 0 |
| GDPH-CRC | a | 82 | 82 | 82 | 0 | 0 | 0 | 0 | 82 | 0 | 0 |
| APH-CRC | a | 118 | 118 | 118 | 0 | 0 | 0 | 0 | 118 | 0 | 0 |
| NFH-CRC | a, c | 465 | 458 | 465 | 0 | 0 | 0 | 0 | 458 | 7 | 0 |
| SYSH-CRC | a | 34 | 34 | 34 | 0 | 0 | 0 | 0 | 34 | 0 | 0 |
| CPTAC-CRC | c, d, e | 110 | 0 | 110 | 108 | 109 | 0 | 107 | 3 | 0 | 0 |
| **GBM** | | | | | | | | | | | |
| TCGA-GBM | a, e | 811 | 811 | 811 | 789 | 703 | 693 | 106 | 12 | 0 | 0 |
| CPTAC-GBM | a, e | 440 | 373 | 440 | 387 | 86 | 80 | 291 | 24 | 45 | 0 |
| **HNSC** | | | | | | | | | | | |
| TCGA-HNSC | a, c, d, e | 471 | 470 | 471 | 456 | 434 | 429 | 32 | 9 | 1 | 0 |
| QDPH-HNSC | a | 201 | 200 | 201 | 0 | 0 | 0 | 0 | 200 | 1 | 0 |
| CPTAC-HNSC | a, c, d, e | 344 | 308 | 344 | 255 | 0 | 0 | 255 | 53 | 36 | 0 |
| **KIRC** | | | | | | | | | | | |
| TCGA-KIRC | a, e | 518 | 518 | 518 | 511 | 366 | 365 | 147 | 6 | 0 | 0 |
| CPTAC-KIRC | a, e | 696 | 609 | 696 | 686 | 242 | 233 | 375 | 88 | 0 | 0 |
| **LIHC** | | | | | | | | | | | |
| TCGA-LIHC | a, c, d, e | 378 | 376 | 378 | 367 | 358 | 354 | 16 | 7 | 1 | 0 |
| ZJH-LIHC | a, c | 75 | 75 | 75 | 0 | 0 | 0 | 0 | 75 | 0 | 0 |
| **LUAD** | | | | | | | | | | | |
| TCGA-LUAD | a, e | 531 | 449 | 468 | 401 | 342 | 342 | 59 | 48 | 19 | 63 |
| CPTAC-LUAD | a, e | 1,111 | 909 | 1,111 | 456 | 142 | 137 | 324 | 448 | 202 | 0 |
| **nCCRCC** | | | | | | | | | | | |
| TCGA-nCCRCC | a | 409 | 401 | 409 | 387 | 330 | 327 | 63 | 11 | 8 | 0 |
| CPTAC-nCCRCC | a | 89 | 89 | 89 | 57 | 13 | 13 | 44 | 32 | 0 | 0 |
| **UCEC** | | | | | | | | | | | |
| TCGA-UCEC | a | 547 | 532 | 535 | 503 | 379 | 372 | 137 | 24 | 2 | 12 |
| CPTAC-UCEC | a | 727 | 727 | 727 | 462 | 218 | 218 | 244 | 265 | 0 | 0 |
| **LUSC** | | | | | | | | | | | |
| TCGA-LUSC | a | 504 | 495 | 504 | 391 | 327 | 324 | 70 | 101 | 9 | 0 |
| CPTAC-LUSC | a | 435 | 435 | 435 | 220 | 62 | 57 | 168 | 210 | 0 | 0 |
| **PAAD** | | | | | | | | | | | |
| TCGA-PAAD | a | 209 | 185 | 209 | 180 | 142 | 142 | 38 | 5 | 24 | 0 |
| CPTAC-PAAD | a | 418 | 418 | 418 | 280 | 60 | 60 | 220 | 138 | 0 | 0 |
| **NSCLC** | | | | | | | | | | | |
| PDL1-NSCLC | b | 193 | 193 | 193 | 0 | 0 | 0 | 0 | 193 | 0 | 0 |
| **OV** | | | | | | | | | | | |
| TCGA-OV | c | 45 | 45 | 45 | 45 | 41 | 41 | 4 | 0 | 0 | 0 |
| HGSOC-OV | c | 346 | 346 | 0 | 252 | 232 | 0 | 162 | 160 | 24 | 0 |
| BEV-OV | b | 286 | 0 | 0 | 0 | 0 | 0 | 0 | 0 | 0 | 286 |
| **PRAD** | | | | | | | | | | | |
| TCGA-PRAD | c, e | 543 | 543 | 543 | 393 | 328 | 325 | 71 | 147 | 0 | 0 |
| PANDA-PRAD | c | 10,616 | 0 | 0 | 0 | 0 | 0 | 0 | 0 | 0 | 10,616 |
| SICAP-PRAD | c | 155 | 0 | 0 | 0 | 0 | 0 | 0 | 0 | 0 | 155 |
| **STAD** | | | | | | | | | | | |
| TCGA-STAD | e | 360 | 357 | 360 | 308 | 259 | 259 | 49 | 49 | 3 | 0 |

**Supplementary Table 26. Performance of different modality combinations on the TCGA-BLCA dataset for prognostic stratification.** C-index: Concordance Index; AUC: area under the ROC curve at 36 months (Methods); Δ: difference from the pathology-only model (Path). N: number of samples. Values are reported with 95% CIs.

| Model | N | C-index | ΔC-index | AUC | ΔAUC |
|---|---|---|---|---|---|
| UNI | 141 | 0.640 (0.557–0.712) | −0.107 | 0.682 (0.572–0.784) | −0.151 |
| Prov-GigaPath | 141 | 0.709 (0.636–0.771) | −0.038 | 0.781 (0.669–0.871) | −0.053 |
| CHIEF | 141 | 0.670 (0.597–0.731) | −0.077 | 0.726 (0.621–0.822) | −0.107 |
| TITAN | 141 | 0.679 (0.598–0.747) | −0.068 | 0.751 (0.644–0.852) | −0.082 |
| GPFM | 141 | 0.693 (0.618–0.756) | −0.054 | 0.759 (0.648–0.856) | −0.074 |
| *TEAM* | | | | | |
| Path | 141 | 0.747 (0.674–0.808) | +0.000 | 0.833 (0.739–0.912) | +0.000 |
| +Stage/Grade | 141 | 0.810 (0.746–0.860) | +0.063 | 0.874 (0.797–0.937) | +0.041 |
| +TME | 141 | 0.819 (0.755–0.867) | +0.072 | 0.882 (0.809–0.941) | +0.049 |
| +Gene | 141 | 0.811 (0.748–0.861) | +0.064 | 0.867 (0.788–0.931) | +0.034 |
| +Stage/Grade+TME | 141 | 0.867 (0.821–0.905) | +0.120 | 0.946 (0.901–0.982) | +0.113 |
| +Stage/Grade+Gene | 141 | 0.869 (0.823–0.908) | +0.122 | 0.948 (0.904–0.983) | +0.115 |
| +TME+Gene | 141 | 0.869 (0.823–0.907) | +0.122 | 0.947 (0.902–0.982) | +0.114 |
| +Stage/Grade +TME+Gene | 141 | 0.868 (0.822–0.907) | +0.121 | 0.946 (0.901–0.982) | +0.113 |

**Supplementary Table 27. Performance of different modality combinations on the QDPH-BLCA dataset for prognostic stratification.** C-index: Concordance Index; AUC: area under the ROC curve at 36 months (Methods); Δ: difference from the pathology-only model (Path). N: number of samples. Values are reported with 95% CIs.

| Model | N | C-index | ΔC-index | AUC | ΔAUC |
|---|---|---|---|---|---|
| UNI | 441 | 0.575 (0.507–0.641) | −0.064 | 0.595 (0.509–0.683) | −0.083 |
| Prov-GigaPath | 441 | 0.638 (0.568–0.706) | −0.001 | 0.638 (0.551–0.717) | −0.040 |
| CHIEF | 441 | 0.617 (0.551–0.687) | −0.022 | 0.619 (0.544–0.692) | −0.059 |
| TITAN | 441 | 0.619 (0.558–0.675) | −0.020 | 0.619 (0.546–0.686) | −0.059 |
| GPFM | 441 | 0.621 (0.555–0.682) | −0.018 | 0.644 (0.569–0.719) | −0.034 |
| *TEAM* | | | | | |
| Path | 441 | 0.639 (0.570–0.701) | +0.000 | 0.678 (0.596–0.749) | +0.000 |
| +Stage/Grade | 441 | 0.676 (0.608–0.739) | +0.037 | 0.815 (0.738–0.879) | +0.137 |
| +TME | 441 | 0.685 (0.619–0.750) | +0.046 | 0.803 (0.729–0.872) | +0.125 |
| +Gene | 441 | 0.683 (0.619–0.747) | +0.044 | 0.806 (0.740–0.868) | +0.128 |
| +Stage/Grade+TME | 441 | 0.701 (0.644–0.757) | +0.062 | 0.849 (0.794–0.898) | +0.171 |
| +Stage/Grade+Gene | 441 | 0.710 (0.649–0.769) | +0.071 | 0.873 (0.816–0.924) | +0.194 |
| +TME+Gene | 441 | 0.695 (0.639–0.748) | +0.055 | 0.828 (0.770–0.880) | +0.150 |
| +Stage/Grade +TME+Gene | 441 | 0.693 (0.637–0.747) | +0.054 | 0.832 (0.779–0.881) | +0.154 |

**Supplementary Table 28. Performance of different modality combinations on the TCGA-BRCA dataset for prognostic stratification.** C-index: Concordance Index; AUC: area under the ROC curve at 36 months (Methods); Δ: difference from the pathology-only model (Path). N: number of samples. Values are reported with 95% CIs.

| Model | N | C-index | ΔC-index | AUC | ΔAUC |
|---|---|---|---|---|---|
| UNI | 338 | 0.713 (0.626–0.786) | −0.074 | 0.812 (0.710–0.897) | −0.090 |
| Prov-GigaPath | 338 | 0.558 (0.463–0.643) | −0.229 | 0.648 (0.536–0.750) | −0.254 |
| CHIEF | 338 | 0.689 (0.601–0.766) | −0.098 | 0.785 (0.681–0.874) | −0.117 |
| TITAN | 338 | 0.706 (0.620–0.782) | −0.081 | 0.807 (0.715–0.886) | −0.095 |
| GPFM | 338 | 0.659 (0.569–0.738) | −0.128 | 0.753 (0.643–0.845) | −0.149 |
| *TEAM* | | | | | |
| Path | 338 | 0.787 (0.716–0.851) | +0.000 | 0.902 (0.837–0.956) | +0.000 |
| +Stage/Grade | 338 | 0.860 (0.792–0.913) | +0.073 | 0.884 (0.807–0.943) | −0.018 |
| +TME | 338 | 0.880 (0.823–0.925) | +0.094 | 0.916 (0.853–0.960) | +0.014 |
| +Gene | 338 | 0.862 (0.797–0.914) | +0.075 | 0.889 (0.814–0.946) | −0.013 |
| +Stage/Grade+TME | 338 | 0.874 (0.812–0.924) | +0.088 | 0.898 (0.822–0.953) | −0.004 |
| +Stage/Grade+Gene | 338 | 0.873 (0.810–0.922) | +0.086 | 0.896 (0.819–0.953) | −0.006 |
| +TME+Gene | 338 | 0.872 (0.809–0.921) | +0.085 | 0.896 (0.819–0.950) | −0.006 |
| +Stage/Grade +TME+Gene | 338 | 0.874 (0.810–0.923) | +0.087 | 0.897 (0.820–0.952) | −0.005 |

**Supplementary Table 29. Performance of different modality combinations on the NFH-BRCA dataset for prognostic stratification.** C-index: Concordance Index; AUC: area under the ROC curve at 36 months (Methods); Δ: difference from the pathology-only model (Path). N: number of samples. Values are reported with 95% CIs.

| Model | N | C-index | ΔC-index | AUC | ΔAUC |
|---|---|---|---|---|---|
| UNI | 225 | 0.622 (0.473–0.750) | −0.057 | 0.667 (0.476–0.857) | +0.000 |
| Prov-GigaPath | 225 | 0.651 (0.346–0.932) | −0.029 | 0.719 (0.413–0.979) | +0.052 |
| CHIEF | 225 | 0.611 (0.335–0.830) | −0.068 | 0.643 (0.396–0.862) | −0.024 |
| TITAN | 225 | 0.655 (0.334–0.925) | −0.025 | 0.757 (0.435–0.974) | +0.090 |
| GPFM | 225 | 0.639 (0.450–0.787) | −0.040 | 0.757 (0.550–0.919) | +0.090 |
| *TEAM* | | | | | |
| Path | 225 | 0.679 (0.437–0.927) | +0.000 | 0.667 (0.412–0.923) | +0.000 |
| +Stage/Grade | 225 | 0.728 (0.501–0.933) | +0.049 | 0.876 (0.647–1.000) | +0.210 |
| +TME | 225 | 0.763 (0.591–0.882) | +0.083 | 0.948 (0.841–1.000) | +0.281 |
| +Gene | 225 | 0.742 (0.558–0.863) | +0.062 | 0.933 (0.783–1.000) | +0.267 |
| +Stage/Grade+TME | 225 | 0.777 (0.690–0.856) | +0.098 | 0.967 (0.906–1.000) | +0.300 |
| +Stage/Grade+Gene | 225 | 0.770 (0.708–0.831) | +0.091 | 0.990 (0.953–1.000) | +0.324 |
| +TME+Gene | 225 | 0.766 (0.564–0.929) | +0.087 | 0.905 (0.763–1.000) | +0.238 |
| +Stage/Grade +TME+Gene | 225 | 0.783 (0.624–0.912) | +0.104 | 0.981 (0.915–1.000) | +0.314 |

**Supplementary Table 30. Performance of different modality combinations on the TCGA-CESC dataset for prognostic stratification.** C-index: Concordance Index; AUC: area under the ROC curve at 36 months (Methods); Δ: difference from the pathology-only model (Path). N: number of samples. Values are reported with 95% CIs.

| Model | N | C-index | ΔC-index | AUC | ΔAUC |
|---|---|---|---|---|---|
| UNI | 82 | 0.629 (0.487–0.758) | −0.175 | 0.739 (0.545–0.907) | −0.140 |
| Prov-GigaPath | 82 | 0.653 (0.530–0.773) | −0.151 | 0.761 (0.575–0.914) | −0.118 |
| CHIEF | 82 | 0.660 (0.533–0.780) | −0.144 | 0.764 (0.569–0.926) | −0.115 |
| TITAN | 82 | 0.709 (0.578–0.829) | −0.095 | 0.811 (0.651–0.942) | −0.068 |
| GPFM | 82 | 0.756 (0.653–0.842) | −0.048 | 0.863 (0.722–0.979) | −0.016 |
| *TEAM* | | | | | |
| Path | 82 | 0.804 (0.698–0.889) | +0.000 | 0.879 (0.744–0.984) | +0.000 |
| +Stage/Grade | 82 | 0.865 (0.750–0.959) | +0.061 | 0.839 (0.657–0.972) | −0.040 |
| +TME | 82 | 0.875 (0.762–0.959) | +0.071 | 0.848 (0.684–0.970) | −0.031 |
| +Gene | 82 | 0.871 (0.767–0.954) | +0.068 | 0.854 (0.698–0.973) | −0.025 |
| +Stage/Grade+TME | 82 | 0.887 (0.780–0.966) | +0.083 | 0.870 (0.714–0.986) | −0.009 |
| +Stage/Grade+Gene | 82 | 0.883 (0.781–0.965) | +0.079 | 0.854 (0.681–0.981) | −0.025 |
| +TME+Gene | 82 | 0.879 (0.776–0.961) | +0.075 | 0.857 (0.684–0.981) | −0.022 |
| +Stage/Grade +TME+Gene | 82 | 0.892 (0.797–0.964) | +0.088 | 0.870 (0.719–0.984) | −0.009 |

**Supplementary Table 31. Performance of different modality combinations on the NFH-CESC dataset for prognostic stratification.** C-index: Concordance Index; AUC: area under the ROC curve at 36 months (Methods); Δ: difference from the pathology-only model (Path). N: number of samples. Values are reported with 95% CIs.

| Model | N | C-index | ΔC-index | AUC | ΔAUC |
|---|---|---|---|---|---|
| UNI | 405 | 0.659 (0.609–0.711) | −0.053 | 0.730 (0.661–0.796) | −0.101 |
| Prov-GigaPath | 405 | 0.690 (0.637–0.744) | −0.022 | 0.801 (0.743–0.859) | −0.030 |
| CHIEF | 405 | 0.641 (0.582–0.703) | −0.071 | 0.724 (0.653–0.797) | −0.107 |
| TITAN | 405 | 0.681 (0.628–0.732) | −0.031 | 0.773 (0.707–0.830) | −0.058 |
| GPFM | 405 | 0.649 (0.600–0.703) | −0.063 | 0.758 (0.690–0.826) | −0.073 |
| *TEAM* | | | | | |
| Path | 405 | 0.712 (0.659–0.762) | +0.000 | 0.831 (0.776–0.886) | +0.000 |
| +Stage/Grade | 405 | 0.739 (0.694–0.783) | +0.027 | 0.848 (0.789–0.901) | +0.017 |
| +TME | 405 | 0.756 (0.717–0.795) | +0.044 | 0.901 (0.856–0.942) | +0.070 |
| +Gene | 405 | 0.766 (0.730–0.797) | +0.054 | 0.911 (0.871–0.947) | +0.080 |
| +Stage/Grade+TME | 405 | 0.777 (0.739–0.815) | +0.065 | 0.912 (0.865–0.946) | +0.081 |
| +Stage/Grade+Gene | 405 | 0.766 (0.728–0.805) | +0.054 | 0.898 (0.847–0.940) | +0.067 |
| +TME+Gene | 405 | 0.770 (0.731–0.808) | +0.058 | 0.885 (0.830–0.931) | +0.054 |
| +Stage/Grade +TME+Gene | 405 | 0.796 (0.756–0.837) | +0.084 | 0.934 (0.898–0.964) | +0.103 |

**Supplementary Table 32. Performance of different modality combinations on the TCGA-CRC dataset for prognostic stratification.** C-index: Concordance Index; AUC: area under the ROC curve at 36 months (Methods); Δ: difference from the pathology-only model (Path). N: number of samples. Values are reported with 95% CIs.

| Model | N | C-index | ΔC-index | AUC | ΔAUC |
|---|---|---|---|---|---|
| UNI | 187 | 0.663 (0.563–0.752) | −0.129 | 0.745 (0.634–0.851) | −0.207 |
| Prov-GigaPath | 187 | 0.651 (0.551–0.736) | −0.141 | 0.727 (0.605–0.834) | −0.225 |
| CHIEF | 187 | 0.664 (0.567–0.753) | −0.127 | 0.747 (0.634–0.851) | −0.206 |
| TITAN | 187 | 0.695 (0.607–0.778) | −0.096 | 0.819 (0.719–0.905) | −0.134 |
| GPFM | 187 | 0.629 (0.525–0.716) | −0.162 | 0.682 (0.553–0.796) | −0.271 |
| *TEAM* | | | | | |
| Path | 187 | 0.792 (0.717–0.857) | +0.000 | 0.952 (0.895–0.996) | +0.000 |
| +Stage/Grade | 187 | 0.836 (0.763–0.892) | +0.045 | 0.794 (0.680–0.897) | −0.158 |
| +TME | 187 | 0.837 (0.765–0.892) | +0.046 | 0.804 (0.691–0.905) | −0.148 |
| +Gene | 187 | 0.841 (0.770–0.895) | +0.049 | 0.812 (0.699–0.910) | −0.140 |
| +Stage/Grade+TME | 187 | 0.876 (0.812–0.926) | +0.084 | 0.857 (0.756–0.940) | −0.096 |
| +Stage/Grade+Gene | 187 | 0.875 (0.811–0.925) | +0.083 | 0.853 (0.750–0.937) | −0.100 |
| +TME+Gene | 187 | 0.874 (0.810–0.925) | +0.083 | 0.853 (0.751–0.936) | −0.100 |
| +Stage/Grade +TME+Gene | 187 | 0.874 (0.811–0.924) | +0.083 | 0.851 (0.748–0.933) | −0.101 |

**Supplementary Table 33. Performance of different modality combinations on the APH-CRC dataset for prognostic stratification.** C-index: Concordance Index; AUC: area under the ROC curve at 36 months (Methods); Δ: difference from the pathology-only model (Path). N: number of samples. Values are reported with 95% CIs.

| Model | N | C-index | ΔC-index | AUC | ΔAUC |
|---|---|---|---|---|---|
| UNI | 118 | 0.638 (0.515–0.772) | −0.054 | 0.639 (0.479–0.783) | −0.076 |
| Prov-GigaPath | 118 | 0.621 (0.501–0.722) | −0.072 | 0.639 (0.494–0.773) | −0.076 |
| CHIEF | 118 | 0.611 (0.485–0.739) | −0.081 | 0.706 (0.574–0.828) | −0.008 |
| TITAN | 118 | 0.608 (0.484–0.732) | −0.085 | 0.632 (0.444–0.807) | −0.082 |
| GPFM | 118 | 0.567 (0.431–0.684) | −0.126 | 0.655 (0.561–0.752) | −0.060 |
| *TEAM* | | | | | |
| Path | 118 | 0.693 (0.600–0.793) | +0.000 | 0.715 (0.584–0.835) | +0.000 |
| +Stage/Grade | 118 | 0.730 (0.631–0.826) | +0.037 | 0.793 (0.690–0.891) | +0.078 |
| +TME | 118 | 0.741 (0.643–0.837) | +0.048 | 0.782 (0.640–0.894) | +0.068 |
| +Gene | 118 | 0.734 (0.638–0.824) | +0.041 | 0.749 (0.629–0.862) | +0.034 |
| +Stage/Grade+TME | 118 | 0.727 (0.633–0.822) | +0.034 | 0.719 (0.579–0.851) | +0.004 |
| +Stage/Grade+Gene | 118 | 0.752 (0.658–0.840) | +0.059 | 0.712 (0.572–0.832) | −0.003 |
| +TME+Gene | 118 | 0.746 (0.642–0.851) | +0.053 | 0.736 (0.604–0.865) | +0.021 |
| +Stage/Grade +TME+Gene | 118 | 0.760 (0.663–0.843) | +0.067 | 0.805 (0.709–0.888) | +0.090 |

**Supplementary Table 34. Performance of different modality combinations on the GDPH-CRC dataset for prognostic stratification.** C-index: Concordance Index; AUC: area under the ROC curve at 36 months (Methods); Δ: difference from the pathology-only model (Path). N: number of samples. Values are reported with 95% CIs.

| Model | N | C-index | ΔC-index | AUC | ΔAUC |
|---|---|---|---|---|---|
| UNI | 82 | 0.618 (0.442–0.787) | −0.051 | 0.689 (0.477–0.890) | +0.000 |
| Prov-GigaPath | 82 | 0.645 (0.464–0.816) | −0.024 | 0.593 (0.369–0.789) | −0.096 |
| CHIEF | 82 | 0.611 (0.408–0.817) | −0.058 | 0.552 (0.248–0.835) | −0.137 |
| TITAN | 82 | 0.650 (0.374–0.878) | −0.019 | 0.785 (0.420–0.984) | +0.096 |
| GPFM | 82 | 0.626 (0.385–0.859) | −0.043 | 0.700 (0.385–1.000) | +0.011 |
| *TEAM* | | | | | |
| Path | 82 | 0.669 (0.516–0.819) | +0.000 | 0.689 (0.509–0.841) | +0.000 |
| +Stage/Grade | 82 | 0.703 (0.449–0.912) | +0.034 | 0.741 (0.444–0.947) | +0.052 |
| +TME | 82 | 0.720 (0.537–0.879) | +0.051 | 0.652 (0.406–0.865) | −0.037 |
| +Gene | 82 | 0.722 (0.551–0.873) | +0.053 | 0.741 (0.504–0.901) | +0.052 |
| +Stage/Grade+TME | 82 | 0.742 (0.530–0.903) | +0.072 | 0.715 (0.396–0.933) | +0.026 |
| +Stage/Grade+Gene | 82 | 0.739 (0.605–0.858) | +0.070 | 0.730 (0.562–0.868) | +0.041 |
| +TME+Gene | 82 | 0.713 (0.491–0.903) | +0.043 | 0.748 (0.457–0.952) | +0.059 |
| +Stage/Grade +TME+Gene | 82 | 0.744 (0.581–0.892) | +0.075 | 0.685 (0.455–0.864) | −0.004 |

**Supplementary Table 35. Performance of different modality combinations on the HBFH-CRC dataset for prognostic stratification.** C-index: Concordance Index; AUC: area under the ROC curve at 36 months (Methods); Δ: difference from the pathology-only model (Path). N: number of samples. Values are reported with 95% CIs.

| Model | N | C-index | ΔC-index | AUC | ΔAUC |
|---|---|---|---|---|---|
| UNI | 49 | 0.616 (0.477–0.748) | −0.057 | 0.603 (0.425–0.759) | −0.141 |
| Prov-GigaPath | 49 | 0.660 (0.533–0.782) | −0.013 | 0.665 (0.490–0.823) | −0.079 |
| CHIEF | 49 | 0.620 (0.457–0.766) | −0.054 | 0.660 (0.482–0.824) | −0.083 |
| TITAN | 49 | 0.620 (0.464–0.778) | −0.054 | 0.626 (0.433–0.816) | −0.118 |
| GPFM | 49 | 0.600 (0.466–0.732) | −0.073 | 0.635 (0.463–0.806) | −0.109 |
| *TEAM* | | | | | |
| Path | 49 | 0.673 (0.546–0.802) | +0.000 | 0.744 (0.594–0.875) | +0.000 |
| +Stage/Grade | 49 | 0.735 (0.625–0.845) | +0.062 | 0.786 (0.630–0.912) | +0.043 |
| +TME | 49 | 0.746 (0.638–0.865) | +0.073 | 0.774 (0.626–0.900) | +0.030 |
| +Gene | 49 | 0.738 (0.621–0.852) | +0.065 | 0.846 (0.723–0.943) | +0.103 |
| +Stage/Grade+TME | 49 | 0.758 (0.641–0.885) | +0.085 | 0.797 (0.639–0.928) | +0.053 |
| +Stage/Grade+Gene | 49 | 0.753 (0.639–0.863) | +0.080 | 0.814 (0.691–0.923) | +0.071 |
| +TME+Gene | 49 | 0.750 (0.623–0.865) | +0.076 | 0.769 (0.625–0.890) | +0.026 |
| +Stage/Grade +TME+Gene | 49 | 0.792 (0.668–0.917) | +0.119 | 0.835 (0.667–0.962) | +0.092 |

**Supplementary Table 36. Performance of different modality combinations on the NFH-CRC dataset for prognostic stratification.** C-index: Concordance Index; AUC: area under the ROC curve at 36 months (Methods); Δ: difference from the pathology-only model (Path). N: number of samples. Values are reported with 95% CIs.

| Model | N | C-index | ΔC-index | AUC | ΔAUC |
|---|---|---|---|---|---|
| UNI | 458 | 0.661 (0.594–0.723) | −0.026 | 0.706 (0.610–0.791) | −0.014 |
| Prov-GigaPath | 458 | 0.662 (0.594–0.724) | −0.025 | 0.681 (0.581–0.771) | −0.039 |
| CHIEF | 458 | 0.638 (0.567–0.702) | −0.050 | 0.685 (0.593–0.783) | −0.035 |
| TITAN | 458 | 0.630 (0.560–0.696) | −0.058 | 0.764 (0.679–0.843) | +0.045 |
| GPFM | 458 | 0.649 (0.586–0.717) | −0.038 | 0.672 (0.586–0.757) | −0.048 |
| *TEAM* | | | | | |
| Path | 458 | 0.688 (0.627–0.742) | +0.000 | 0.720 (0.637–0.804) | +0.000 |
| +Stage/Grade | 458 | 0.720 (0.655–0.782) | +0.032 | 0.760 (0.662–0.839) | +0.040 |
| +TME | 458 | 0.734 (0.679–0.787) | +0.046 | 0.776 (0.693–0.849) | +0.057 |
| +Gene | 458 | 0.722 (0.669–0.773) | +0.034 | 0.689 (0.612–0.760) | −0.030 |
| +Stage/Grade+TME | 458 | 0.723 (0.671–0.772) | +0.036 | 0.767 (0.696–0.836) | +0.048 |
| +Stage/Grade+Gene | 458 | 0.752 (0.702–0.798) | +0.064 | 0.778 (0.707–0.841) | +0.058 |
| +TME+Gene | 458 | 0.727 (0.666–0.788) | +0.039 | 0.716 (0.623–0.802) | −0.004 |
| +Stage/Grade +TME+Gene | 458 | 0.740 (0.680–0.797) | +0.052 | 0.744 (0.654–0.833) | +0.024 |

**Supplementary Table 37. Performance of different modality combinations on the SYSH-CRC dataset for prognostic stratification.** C-index: Concordance Index; AUC: area under the ROC curve at 36 months (Methods); Δ: difference from the pathology-only model (Path). N: number of samples. Values are reported with 95% CIs.

| Model | N | C-index | ΔC-index | AUC | ΔAUC |
|---|---|---|---|---|---|
| UNI | 34 | 0.599 (0.336–0.812) | −0.088 | 0.542 (0.171–0.828) | −0.125 |
| Prov-GigaPath | 34 | 0.610 (0.292–0.856) | −0.077 | 0.525 (0.063–0.873) | −0.142 |
| CHIEF | 34 | 0.599 (0.380–0.788) | −0.088 | 0.725 (0.545–0.875) | +0.058 |
| TITAN | 34 | 0.621 (0.392–0.857) | −0.066 | 0.508 (0.250–0.742) | −0.158 |
| GPFM | 34 | 0.593 (0.355–0.842) | −0.093 | 0.500 (0.248–0.766) | −0.167 |
| *TEAM* | | | | | |
| Path | 34 | 0.687 (0.427–0.919) | +0.000 | 0.667 (0.257–0.968) | +0.000 |
| +Stage/Grade | 34 | 0.731 (0.431–0.961) | +0.044 | 0.692 (0.207–1.000) | +0.025 |
| +TME | 34 | 0.797 (0.570–0.967) | +0.110 | 0.875 (0.689–1.000) | +0.208 |
| +Gene | 34 | 0.780 (0.520–0.992) | +0.093 | 0.742 (0.308–1.000) | +0.075 |
| +Stage/Grade+TME | 34 | 0.813 (0.644–0.939) | +0.126 | 0.867 (0.694–1.000) | +0.200 |
| +Stage/Grade+Gene | 34 | 0.808 (0.544–0.983) | +0.121 | 0.750 (0.332–0.986) | +0.083 |
| +TME+Gene | 34 | 0.802 (0.625–0.956) | +0.115 | 0.758 (0.498–1.000) | +0.092 |
| +Stage/Grade +TME+Gene | 34 | 0.824 (0.674–0.958) | +0.137 | 0.750 (0.548–0.925) | +0.083 |

**Supplementary Table 38. Performance of different modality combinations on the TCGA-GBM dataset for prognostic stratification.** C-index: Concordance Index; AUC: area under the ROC curve at 36 months (Methods); Δ: difference from the pathology-only model (Path). N: number of samples. Values are reported with 95% CIs.

| Model | N | C-index | ΔC-index | AUC | ΔAUC |
|---|---|---|---|---|---|
| UNI | 278 | 0.626 (0.584–0.667) | −0.064 | 0.880 (0.833–0.924) | −0.050 |
| Prov-GigaPath | 278 | 0.610 (0.567–0.653) | −0.080 | 0.826 (0.761–0.890) | −0.104 |
| CHIEF | 278 | 0.576 (0.534–0.618) | −0.113 | 0.812 (0.750–0.870) | −0.118 |
| TITAN | 278 | 0.602 (0.558–0.647) | −0.087 | 0.822 (0.757–0.884) | −0.108 |
| GPFM | 278 | 0.660 (0.621–0.703) | −0.029 | 0.889 (0.833–0.940) | −0.041 |
| *TEAM* | | | | | |
| Path | 278 | 0.689 (0.647–0.727) | +0.000 | 0.930 (0.891–0.965) | +0.000 |
| +Stage/Grade | 278 | 0.721 (0.684–0.758) | +0.032 | 0.948 (0.913–0.978) | +0.018 |
| +TME | 278 | 0.734 (0.698–0.770) | +0.045 | 0.952 (0.918–0.980) | +0.022 |
| +Gene | 278 | 0.725 (0.688–0.761) | +0.036 | 0.949 (0.915–0.978) | +0.019 |
| +Stage/Grade+TME | 278 | 0.761 (0.727–0.793) | +0.072 | 0.964 (0.937–0.987) | +0.034 |
| +Stage/Grade+Gene | 278 | 0.754 (0.719–0.787) | +0.065 | 0.961 (0.932–0.985) | +0.031 |
| +TME+Gene | 278 | 0.769 (0.735–0.802) | +0.080 | 0.966 (0.940–0.988) | +0.037 |
| +Stage/Grade +TME+Gene | 278 | 0.786 (0.753–0.815) | +0.096 | 0.971 (0.946–0.991) | +0.041 |

**Supplementary Table 39. Performance of different modality combinations on the CPTAC-GBM dataset for prognostic stratification.** C-index: Concordance Index; AUC: area under the ROC curve at 36 months (Methods); Δ: difference from the pathology-only model (Path). N: number of samples. Values are reported with 95% CIs.

| Model | N | C-index | ΔC-index | AUC | ΔAUC |
|---|---|---|---|---|---|
| UNI | 373 | 0.550 (0.508–0.587) | −0.062 | 0.524 (0.434–0.608) | −0.100 |
| Prov-GigaPath | 373 | 0.569 (0.530–0.607) | −0.043 | 0.522 (0.430–0.611) | −0.102 |
| CHIEF | 373 | 0.580 (0.539–0.616) | −0.032 | 0.565 (0.478–0.650) | −0.058 |
| TITAN | 373 | 0.570 (0.531–0.608) | −0.042 | 0.523 (0.431–0.613) | −0.101 |
| GPFM | 373 | 0.532 (0.491–0.568) | −0.080 | 0.485 (0.393–0.571) | −0.139 |
| *TEAM* | | | | | |
| Path | 373 | 0.612 (0.574–0.646) | +0.000 | 0.624 (0.537–0.708) | +0.000 |
| +Stage/Grade | 373 | 0.642 (0.606–0.674) | +0.030 | 0.668 (0.586–0.749) | +0.044 |
| +TME | 373 | 0.656 (0.620–0.687) | +0.044 | 0.685 (0.606–0.765) | +0.061 |
| +Gene | 373 | 0.647 (0.610–0.679) | +0.035 | 0.674 (0.594–0.756) | +0.050 |
| +Stage/Grade+TME | 373 | 0.688 (0.655–0.717) | +0.076 | 0.728 (0.652–0.801) | +0.105 |
| +Stage/Grade+Gene | 373 | 0.678 (0.644–0.708) | +0.066 | 0.712 (0.635–0.789) | +0.088 |
| +TME+Gene | 373 | 0.696 (0.664–0.726) | +0.084 | 0.741 (0.666–0.814) | +0.117 |
| +Stage/Grade +TME+Gene | 373 | 0.716 (0.685–0.744) | +0.104 | 0.767 (0.697–0.835) | +0.143 |

**Supplementary Table 40. Performance of different modality combinations on the TCGA-HNSC dataset for prognostic stratification.** C-index: Concordance Index; AUC: area under the ROC curve at 36 months (Methods); Δ: difference from the pathology-only model (Path). N: number of samples. Values are reported with 95% CIs.

| Model | N | C-index | ΔC-index | AUC | ΔAUC |
|---|---|---|---|---|---|
| UNI | 137 | 0.707 (0.641–0.762) | −0.057 | 0.821 (0.725–0.897) | −0.009 |
| Prov-GigaPath | 137 | 0.733 (0.666–0.794) | −0.031 | 0.846 (0.763–0.918) | +0.017 |
| CHIEF | 137 | 0.695 (0.624–0.758) | −0.069 | 0.779 (0.682–0.865) | −0.051 |
| TITAN | 137 | 0.744 (0.672–0.804) | −0.020 | 0.814 (0.716–0.891) | −0.016 |
| GPFM | 137 | 0.722 (0.654–0.782) | −0.042 | 0.781 (0.681–0.862) | −0.048 |
| *TEAM* | | | | | |
| Path | 137 | 0.764 (0.699–0.817) | +0.000 | 0.829 (0.737–0.900) | +0.000 |
| +Stage/Grade | 137 | 0.817 (0.764–0.861) | +0.053 | 0.884 (0.812–0.944) | +0.055 |
| +TME | 137 | 0.830 (0.778–0.874) | +0.066 | 0.895 (0.828–0.950) | +0.066 |
| +Gene | 137 | 0.820 (0.769–0.867) | +0.056 | 0.887 (0.816–0.944) | +0.057 |
| +Stage/Grade+TME | 137 | 0.841 (0.795–0.882) | +0.077 | 0.926 (0.869–0.970) | +0.097 |
| +Stage/Grade+Gene | 137 | 0.838 (0.792–0.882) | +0.074 | 0.923 (0.866–0.969) | +0.094 |
| +TME+Gene | 137 | 0.834 (0.788–0.878) | +0.070 | 0.924 (0.868–0.969) | +0.095 |
| +Stage/Grade +TME+Gene | 137 | 0.845 (0.798–0.886) | +0.081 | 0.930 (0.874–0.972) | +0.101 |

**Supplementary Table 41. Performance of different modality combinations on the CPTAC-HNSC dataset for prognostic stratification.** C-index: Concordance Index; AUC: area under the ROC curve at 36 months (Methods); Δ: difference from the pathology-only model (Path). N: number of samples. Values are reported with 95% CIs.

| Model | N | C-index | ΔC-index | AUC | ΔAUC |
|---|---|---|---|---|---|
| UNI | 308 | 0.588 (0.518–0.657) | −0.059 | 0.614 (0.528–0.693) | −0.031 |
| Prov-GigaPath | 308 | 0.615 (0.550–0.680) | −0.033 | 0.612 (0.530–0.693) | −0.033 |
| CHIEF | 308 | 0.537 (0.461–0.607) | −0.110 | 0.550 (0.460–0.636) | −0.095 |
| TITAN | 308 | 0.625 (0.554–0.685) | −0.023 | 0.648 (0.564–0.725) | +0.004 |
| GPFM | 308 | 0.595 (0.531–0.658) | −0.053 | 0.554 (0.465–0.636) | −0.090 |
| *TEAM* | | | | | |
| Path | 308 | 0.648 (0.580–0.713) | +0.000 | 0.644 (0.557–0.724) | +0.000 |
| +Stage/Grade | 308 | 0.682 (0.625–0.735) | +0.034 | 0.773 (0.701–0.840) | +0.129 |
| +TME | 308 | 0.673 (0.626–0.719) | +0.025 | 0.740 (0.658–0.816) | +0.096 |
| +Gene | 308 | 0.678 (0.622–0.733) | +0.030 | 0.776 (0.704–0.845) | +0.132 |
| +Stage/Grade+TME | 308 | 0.686 (0.637–0.733) | +0.038 | 0.755 (0.677–0.830) | +0.110 |
| +Stage/Grade+Gene | 308 | 0.686 (0.632–0.737) | +0.038 | 0.724 (0.646–0.797) | +0.079 |
| +TME+Gene | 308 | 0.691 (0.632–0.743) | +0.044 | 0.766 (0.690–0.831) | +0.121 |
| +Stage/Grade +TME+Gene | 308 | 0.705 (0.651–0.758) | +0.057 | 0.798 (0.728–0.861) | +0.154 |

**Supplementary Table 42. Performance of different modality combinations on the QDPH-HNSC dataset for prognostic stratification.** C-index: Concordance Index; AUC: area under the ROC curve at 36 months (Methods); Δ: difference from the pathology-only model (Path). N: number of samples. Values are reported with 95% CIs.

| **Model** | **N** | **C-index** | **ΔC-index** | **AUC** | **ΔAUC** |
|---|---|---|---|---|---|
| UNI | 201 | 0.661 (0.590–0.726) | −0.018 | 0.763 (0.675–0.842) | −0.038 |
| Prov-GigaPath | 201 | 0.618 (0.549–0.682) | −0.061 | 0.689 (0.583–0.775) | −0.112 |
| CHIEF | 201 | 0.641 (0.569–0.705) | −0.038 | 0.739 (0.648–0.826) | −0.062 |
| TITAN | 201 | 0.610 (0.535–0.674) | −0.069 | 0.748 (0.655–0.828) | −0.053 |
| GPFM | 201 | 0.652 (0.578–0.718) | −0.027 | 0.780 (0.694–0.855) | −0.021 |
| *TEAM* | | | | | |
| Path | 201 | 0.679 (0.607–0.742) | +0.000 | 0.801 (0.709–0.868) | +0.000 |
| +Stage/Grade | 201 | 0.700 (0.636–0.756) | +0.021 | 0.777 (0.682–0.853) | −0.024 |
| +TME | 201 | 0.719 (0.658–0.773) | +0.040 | 0.807 (0.723–0.882) | +0.006 |
| +Gene | 201 | 0.725 (0.671–0.779) | +0.046 | 0.799 (0.716–0.882) | −0.002 |
| +Stage/Grade+TME | 201 | 0.719 (0.667–0.771) | +0.040 | 0.814 (0.735–0.885) | +0.013 |
| +Stage/Grade+Gene | 201 | 0.713 (0.649–0.774) | +0.033 | 0.726 (0.632–0.812) | −0.075 |
| +TME+Gene | 201 | 0.746 (0.693–0.796) | +0.067 | 0.846 (0.774–0.909) | +0.045 |
| +Stage/Grade +TME+Gene | 201 | 0.746 (0.689–0.802) | +0.067 | 0.823 (0.746–0.888) | +0.022 |

**Supplementary Table 43. Performance of different modality combinations on the TCGA-KIRC dataset for prognostic stratification.** C-index: Concordance Index; AUC: area under the ROC curve at 36 months (Methods); Δ: difference from the pathology-only model (Path). N: number of samples. Values are reported with 95% CIs.

| **Model** | **N** | **C-index** | **ΔC-index** | **AUC** | **ΔAUC** |
|---|---|---|---|---|---|
| UNI | 154 | 0.580 (0.501–0.653) | −0.109 | 0.584 (0.463–0.701) | −0.143 |
| Prov-GigaPath | 154 | 0.599 (0.523–0.672) | −0.090 | 0.603 (0.495–0.711) | −0.124 |
| CHIEF | 154 | 0.620 (0.543–0.692) | −0.068 | 0.646 (0.525–0.761) | −0.081 |
| TITAN | 154 | 0.517 (0.437–0.591) | −0.171 | 0.518 (0.407–0.626) | −0.209 |
| GPFM | 154 | 0.686 (0.609–0.757) | −0.002 | 0.726 (0.624–0.823) | −0.001 |
| *TEAM* | | | | | |
| Path | 154 | 0.688 (0.612–0.759) | +0.000 | 0.727 (0.625–0.823) | +0.000 |
| +Stage/Grade | 154 | 0.723 (0.657–0.792) | +0.034 | 0.717 (0.614–0.820) | −0.010 |
| +TME | 154 | 0.733 (0.667–0.800) | +0.045 | 0.729 (0.629–0.829) | +0.002 |
| +Gene | 154 | 0.729 (0.665–0.798) | +0.041 | 0.725 (0.625–0.820) | −0.002 |
| +Stage/Grade+TME | 154 | 0.740 (0.677–0.807) | +0.052 | 0.739 (0.641–0.835) | +0.012 |
| +Stage/Grade+Gene | 154 | 0.738 (0.674–0.803) | +0.050 | 0.736 (0.634–0.833) | +0.009 |
| +TME+Gene | 154 | 0.736 (0.672–0.803) | +0.048 | 0.733 (0.631–0.831) | +0.006 |
| +Stage/Grade +TME+Gene | 154 | 0.745 (0.685–0.810) | +0.057 | 0.743 (0.646–0.837) | +0.016 |

**Supplementary Table 44. Performance of different modality combinations on the CPTAC-KIRC dataset for prognostic stratification.** C-index: Concordance Index; AUC: area under the ROC curve at 36 months (Methods); Δ: difference from the pathology-only model (Path). N: number of samples. Values are reported with 95% CIs.

| Model | N | C-index | ΔC-index | AUC | ΔAUC |
|---|---|---|---|---|---|
| UNI | 608 | 0.635 (0.584–0.688) | −0.025 | 0.616 (0.553–0.685) | −0.085 |
| Prov-GigaPath | 608 | 0.633 (0.582–0.688) | −0.027 | 0.613 (0.548–0.681) | −0.088 |
| CHIEF | 608 | 0.647 (0.597–0.701) | −0.013 | 0.633 (0.572–0.699) | −0.068 |
| TITAN | 608 | 0.611 (0.558–0.667) | −0.050 | 0.586 (0.522–0.654) | −0.116 |
| GPFM | 608 | 0.601 (0.548–0.658) | −0.060 | 0.575 (0.507–0.644) | −0.126 |
| *TEAM* | | | | | |
| Path | 608 | 0.661 (0.608–0.709) | +0.000 | 0.701 (0.628–0.769) | +0.000 |
| +Stage/Grade | 608 | 0.693 (0.642–0.743) | +0.033 | 0.775 (0.709–0.842) | +0.073 |
| +TME | 608 | 0.681 (0.633–0.724) | +0.020 | 0.781 (0.714–0.848) | +0.080 |
| +Gene | 608 | 0.695 (0.646–0.740) | +0.034 | 0.820 (0.753–0.883) | +0.118 |
| +Stage/Grade+TME | 608 | 0.709 (0.658–0.755) | +0.048 | 0.815 (0.748–0.876) | +0.114 |
| +Stage/Grade+Gene | 608 | 0.701 (0.658–0.741) | +0.040 | 0.819 (0.750–0.886) | +0.117 |
| +TME+Gene | 608 | 0.733 (0.689–0.776) | +0.073 | 0.846 (0.788–0.895) | +0.145 |
| +Stage/Grade +TME+Gene | 608 | 0.729 (0.689–0.771) | +0.069 | 0.842 (0.784–0.898) | +0.141 |

**Supplementary Table 45. Performance of different modality combinations on the TCGA-LIHC dataset for prognostic stratification.** C-index: Concordance Index; AUC: area under the ROC curve at 36 months (Methods); Δ: difference from the pathology-only model (Path). N: number of samples. Values are reported with 95% CIs.

| Model | N | C-index | ΔC-index | AUC | ΔAUC |
|---|---|---|---|---|---|
| UNI | 112 | 0.623 (0.509–0.723) | −0.174 | 0.696 (0.560–0.810) | −0.271 |
| Prov-GigaPath | 112 | 0.632 (0.516–0.731) | −0.165 | 0.755 (0.629–0.862) | −0.211 |
| CHIEF | 112 | 0.676 (0.576–0.764) | −0.121 | 0.816 (0.695–0.911) | −0.151 |
| TITAN | 112 | 0.631 (0.510–0.734) | −0.166 | 0.736 (0.604–0.850) | −0.230 |
| GPFM | 112 | 0.655 (0.537–0.748) | −0.142 | 0.798 (0.664–0.901) | −0.169 |
| *TEAM* | | | | | |
| Path | 112 | 0.797 (0.721–0.860) | +0.000 | 0.967 (0.901–1.000) | +0.000 |
| +Stage/Grade | 112 | 0.868 (0.813–0.917) | +0.071 | 0.945 (0.884–0.987) | −0.022 |
| +TME | 112 | 0.884 (0.840–0.928) | +0.088 | 0.967 (0.924–0.994) | +0.000 |
| +Gene | 112 | 0.876 (0.826–0.923) | +0.079 | 0.964 (0.917–0.992) | −0.003 |
| +Stage/Grade+TME | 112 | 0.882 (0.834–0.929) | +0.085 | 0.951 (0.897–0.988) | −0.016 |
| +Stage/Grade+Gene | 112 | 0.883 (0.834–0.929) | +0.086 | 0.949 (0.894–0.986) | −0.018 |
| +TME+Gene | 112 | 0.890 (0.843–0.935) | +0.093 | 0.957 (0.908–0.991) | −0.009 |
| +Stage/Grade +TME+Gene | 112 | 0.886 (0.838–0.931) | +0.089 | 0.952 (0.899–0.988) | −0.015 |

**Supplementary Table 46. Performance of different modality combinations on the ZJH-LIHC dataset for prognostic stratification.** C-index: Concordance Index; AUC: area under the ROC curve at 36 months (Methods); Δ: difference from the pathology-only model (Path). N: number of samples. Values are reported with 95% CIs.

| Model | N | C-index | ΔC-index | AUC | ΔAUC |
|---|---|---|---|---|---|
| UNI | 75 | 0.620 (0.329–0.835) | −0.062 | 0.636 (0.319–0.913) | −0.168 |
| Prov-GigaPath | 75 | 0.640 (0.277–0.868) | −0.042 | 0.709 (0.333–0.937) | −0.095 |
| CHIEF | 75 | 0.620 (0.467–0.801) | −0.062 | 0.714 (0.553–0.854) | −0.091 |
| TITAN | 75 | 0.649 (0.418–0.882) | −0.034 | 0.682 (0.400–0.929) | −0.123 |
| GPFM | 75 | 0.640 (0.413–0.928) | −0.042 | 0.632 (0.335–0.900) | −0.173 |
| *TEAM* | | | | | |
| Path | 75 | 0.683 (0.524–0.779) | +0.000 | 0.805 (0.621–0.941) | +0.000 |
| +Stage/Grade | 75 | 0.703 (0.459–0.863) | +0.020 | 0.814 (0.595–0.979) | +0.009 |
| +TME | 75 | 0.725 (0.560–0.864) | +0.042 | 0.918 (0.799–1.000) | +0.114 |
| +Gene | 75 | 0.717 (0.578–0.913) | +0.034 | 0.891 (0.781–0.970) | +0.086 |
| +Stage/Grade+TME | 75 | 0.734 (0.613–0.870) | +0.051 | 0.864 (0.756–0.957) | +0.059 |
| +Stage/Grade+Gene | 75 | 0.745 (0.599–0.886) | +0.062 | 0.895 (0.771–0.986) | +0.091 |
| +TME+Gene | 75 | 0.722 (0.569–0.846) | +0.040 | 0.823 (0.705–0.925) | +0.018 |
| +Stage/Grade +TME+Gene | 75 | 0.748 (0.631–0.877) | +0.065 | 0.882 (0.777–0.963) | +0.077 |

**Supplementary Table 47. Performance of different modality combinations on the TCGA-LUAD dataset for prognostic stratification.** C-index: Concordance Index; AUC: area under the ROC curve at 36 months (Methods); Δ: difference from the pathology-only model (Path). N: number of samples. Values are reported with 95% CIs.

| Model | N | C-index | ΔC-index | AUC | ΔAUC |
|---|---|---|---|---|---|
| UNI | 165 | 0.629 (0.552–0.700) | −0.074 | 0.761 (0.665–0.855) | −0.088 |
| Prov-GigaPath | 165 | 0.592 (0.522–0.659) | −0.112 | 0.747 (0.655–0.848) | −0.102 |
| CHIEF | 165 | 0.620 (0.556–0.684) | −0.084 | 0.753 (0.661–0.848) | −0.096 |
| TITAN | 165 | 0.589 (0.515–0.663) | −0.114 | 0.709 (0.599–0.811) | −0.141 |
| GPFM | 165 | 0.563 (0.493–0.631) | −0.140 | 0.683 (0.575–0.797) | −0.166 |
| *TEAM* | | | | | |
| Path | 165 | 0.703 (0.631–0.772) | +0.000 | 0.849 (0.773–0.928) | +0.000 |
| +Stage/Grade | 165 | 0.737 (0.666–0.802) | +0.034 | 0.878 (0.807–0.949) | +0.028 |
| +TME | 165 | 0.752 (0.684–0.818) | +0.049 | 0.884 (0.815–0.953) | +0.035 |
| +Gene | 165 | 0.744 (0.674–0.810) | +0.041 | 0.882 (0.812–0.953) | +0.032 |
| +Stage/Grade+TME | 165 | 0.774 (0.709–0.837) | +0.071 | 0.896 (0.831–0.959) | +0.047 |
| +Stage/Grade+Gene | 165 | 0.767 (0.701–0.830) | +0.063 | 0.894 (0.827–0.959) | +0.044 |
| +TME+Gene | 165 | 0.781 (0.717–0.842) | +0.078 | 0.900 (0.836–0.961) | +0.051 |
| +Stage/Grade +TME+Gene | 165 | 0.796 (0.735–0.854) | +0.093 | 0.909 (0.849–0.966) | +0.060 |

**Supplementary Table 48. Performance of different modality combinations on the CPTAC-LUAD dataset for prognostic stratification.** C-index: Concordance Index; AUC: area under the ROC curve at 36 months (Methods); Δ: difference from the pathology-only model (Path). N: number of samples. Values are reported with 95% CIs.

| Model | N | C-index | ΔC-index | AUC | ΔAUC |
|---|---|---|---|---|---|
| UNI | 909 | 0.570 (0.522–0.617) | −0.071 | 0.577 (0.512–0.635) | −0.137 |
| Prov-GigaPath | 909 | 0.587 (0.540–0.632) | −0.054 | 0.589 (0.525–0.646) | −0.125 |
| CHIEF | 909 | 0.618 (0.572–0.663) | −0.023 | 0.633 (0.572–0.690) | −0.081 |
| TITAN | 909 | 0.638 (0.599–0.681) | −0.002 | 0.656 (0.601–0.711) | −0.058 |
| GPFM | 909 | 0.606 (0.560–0.650) | −0.035 | 0.575 (0.518–0.631) | −0.139 |
| *TEAM* | | | | | |
| Path | 909 | 0.641 (0.603–0.679) | +0.000 | 0.714 (0.662–0.764) | +0.000 |
| +Stage/Grade | 909 | 0.676 (0.640–0.712) | +0.035 | 0.749 (0.700–0.797) | +0.035 |
| +TME | 909 | 0.690 (0.655–0.725) | +0.049 | 0.762 (0.713–0.808) | +0.048 |
| +Gene | 909 | 0.684 (0.648–0.719) | +0.043 | 0.756 (0.707–0.803) | +0.042 |
| +Stage/Grade+TME | 909 | 0.713 (0.680–0.747) | +0.073 | 0.783 (0.737–0.829) | +0.069 |
| +Stage/Grade+Gene | 909 | 0.704 (0.670–0.738) | +0.063 | 0.774 (0.727–0.820) | +0.060 |
| +TME+Gene | 909 | 0.721 (0.688–0.754) | +0.081 | 0.790 (0.744–0.835) | +0.076 |
| +Stage/Grade +TME+Gene | 909 | 0.735 (0.702–0.767) | +0.094 | 0.803 (0.758–0.846) | +0.088 |

**Supplementary Table 49. Performance of different modality combinations on the TCGA-LUSC dataset for prognostic stratification.** C-index: Concordance Index; AUC: area under the ROC curve at 36 months (Methods); Δ: difference from the pathology-only model (Path). N: number of samples. Values are reported with 95% CIs.

| Model | N | C-index | ΔC-index | AUC | ΔAUC |
|---|---|---|---|---|---|
| UNI | 153 | 0.645 (0.567–0.718) | −0.044 | 0.751 (0.655–0.838) | −0.107 |
| Prov-GigaPath | 153 | 0.640 (0.563–0.712) | −0.049 | 0.756 (0.655–0.841) | −0.103 |
| CHIEF | 153 | 0.614 (0.530–0.691) | −0.075 | 0.673 (0.568–0.772) | −0.186 |
| TITAN | 153 | 0.669 (0.601–0.732) | −0.020 | 0.836 (0.754–0.909) | −0.023 |
| GPFM | 153 | 0.624 (0.547–0.700) | −0.065 | 0.706 (0.605–0.801) | −0.153 |
| *TEAM* | | | | | |
| Path | 153 | 0.689 (0.618–0.754) | +0.000 | 0.859 (0.778–0.927) | +0.000 |
| +Stage/Grade | 153 | 0.721 (0.658–0.775) | +0.032 | 0.885 (0.817–0.947) | +0.026 |
| +TME | 153 | 0.737 (0.675–0.791) | +0.048 | 0.893 (0.828–0.954) | +0.035 |
| +Gene | 153 | 0.728 (0.666–0.781) | +0.039 | 0.892 (0.826–0.952) | +0.033 |
| +Stage/Grade+TME | 153 | 0.761 (0.702–0.809) | +0.072 | 0.911 (0.851–0.966) | +0.053 |
| +Stage/Grade+Gene | 153 | 0.753 (0.693–0.802) | +0.064 | 0.906 (0.843–0.962) | +0.047 |
| +TME+Gene | 153 | 0.767 (0.710–0.814) | +0.078 | 0.916 (0.856–0.968) | +0.057 |
| +Stage/Grade +TME+Gene | 153 | 0.781 (0.726–0.825) | +0.091 | 0.921 (0.862–0.971) | +0.062 |

**Supplementary Table 50. Performance of different modality combinations on the CPTAC-LUSC dataset for prognostic stratification.** C-index: Concordance Index; AUC: area under the ROC curve at 36 months (Methods); Δ: difference from the pathology-only model (Path). N: number of samples. Values are reported with 95% CIs.

| Model | N | C-index | ΔC-index | AUC | ΔAUC |
|---|---|---|---|---|---|
| UNI | 435 | 0.526 (0.473–0.582) | −0.069 | 0.572 (0.502–0.634) | −0.096 |
| Prov-GigaPath | 435 | 0.551 (0.496–0.604) | −0.045 | 0.579 (0.510–0.645) | −0.088 |
| CHIEF | 435 | 0.563 (0.511–0.614) | −0.033 | 0.596 (0.532–0.661) | −0.072 |
| TITAN | 435 | 0.566 (0.512–0.618) | −0.029 | 0.588 (0.520–0.657) | −0.080 |
| GPFM | 435 | 0.535 (0.484–0.589) | −0.060 | 0.561 (0.492–0.625) | −0.106 |
| *TEAM* | | | | | |
| Path | 435 | 0.596 (0.546–0.648) | +0.000 | 0.667 (0.604–0.730) | +0.000 |
| +Stage/Grade | 435 | 0.629 (0.578–0.681) | +0.033 | 0.707 (0.638–0.766) | +0.040 |
| +TME | 435 | 0.647 (0.597–0.695) | +0.051 | 0.728 (0.665–0.785) | +0.061 |
| +Gene | 435 | 0.639 (0.588–0.688) | +0.043 | 0.719 (0.652–0.776) | +0.051 |
| +Stage/Grade+TME | 435 | 0.661 (0.610–0.708) | +0.065 | 0.743 (0.681–0.798) | +0.076 |
| +Stage/Grade+Gene | 435 | 0.659 (0.609–0.706) | +0.063 | 0.743 (0.681–0.798) | +0.075 |
| +TME+Gene | 435 | 0.669 (0.621–0.716) | +0.074 | 0.754 (0.695–0.809) | +0.087 |
| +Stage/Grade +TME+Gene | 435 | 0.689 (0.642–0.733) | +0.093 | 0.777 (0.720–0.829) | +0.109 |

**Supplementary Table 51. Performance of different modality combinations on the TCGA-nCCRCC dataset for prognostic stratification.** C-index: Concordance Index; AUC: area under the ROC curve at 36 months (Methods); Δ: difference from the pathology-only model (Path). N: number of samples. Values are reported with 95% CIs.

| Model | N | C-index | ΔC-index | AUC | ΔAUC |
|---|---|---|---|---|---|
| UNI | 120 | 0.594 (0.470–0.712) | −0.247 | 0.759 (0.609–0.894) | −0.160 |
| Prov-GigaPath | 120 | 0.805 (0.678–0.912) | −0.036 | 0.883 (0.752–0.991) | −0.035 |
| CHIEF | 120 | 0.780 (0.647–0.882) | −0.062 | 0.904 (0.774–0.997) | −0.014 |
| TITAN | 120 | 0.823 (0.677–0.930) | −0.018 | 0.894 (0.746–1.000) | −0.025 |
| GPFM | 120 | 0.814 (0.707–0.915) | −0.028 | 0.920 (0.819–0.995) | +0.002 |
| *TEAM* | | | | | |
| Path | 120 | 0.841 (0.702–0.935) | +0.000 | 0.918 (0.801–0.997) | +0.000 |
| +Stage/Grade | 120 | 0.874 (0.759–0.957) | +0.033 | 0.940 (0.844–1.000) | +0.021 |
| +TME | 120 | 0.889 (0.781–0.964) | +0.048 | 0.954 (0.880–1.000) | +0.035 |
| +Gene | 120 | 0.882 (0.770–0.960) | +0.040 | 0.949 (0.867–1.000) | +0.030 |
| +Stage/Grade+TME | 120 | 0.915 (0.815–0.978) | +0.073 | 0.970 (0.911–1.000) | +0.051 |
| +Stage/Grade+Gene | 120 | 0.905 (0.802–0.973) | +0.064 | 0.963 (0.898–1.000) | +0.044 |
| +TME+Gene | 120 | 0.923 (0.826–0.983) | +0.082 | 0.972 (0.918–1.000) | +0.053 |
| +Stage/Grade +TME+Gene | 120 | 0.935 (0.856–0.983) | +0.094 | 0.980 (0.940–1.000) | +0.062 |

**Supplementary Table 52. Performance of different modality combinations on the CPTAC-nCCRCC dataset for prognostic stratification.** C-index: Concordance Index; AUC: area under the ROC curve at 36 months (Methods); Δ: difference from the pathology-only model (Path). N: number of samples. Values are reported with 95% CIs.

| Model | N | C-index | ΔC-index | AUC | ΔAUC |
|---|---|---|---|---|---|
| UNI | 89 | 0.659 (0.385–0.856) | −0.072 | 0.701 (0.283–0.986) | −0.030 |
| Prov-GigaPath | 89 | 0.679 (0.436–0.861) | −0.051 | 0.706 (0.329–0.975) | −0.026 |
| CHIEF | 89 | 0.663 (0.384–0.862) | −0.068 | 0.703 (0.283–0.988) | −0.028 |
| TITAN | 89 | 0.677 (0.403–0.881) | −0.054 | 0.697 (0.289–0.980) | −0.035 |
| GPFM | 89 | 0.729 (0.502–0.903) | −0.001 | 0.732 (0.371–0.986) | +0.000 |
| *TEAM* | | | | | |
| Path | 89 | 0.731 (0.502–0.903) | +0.000 | 0.732 (0.371–0.986) | +0.000 |
| +Stage/Grade | 89 | 0.721 (0.507–0.885) | −0.010 | 0.729 (0.386–0.976) | −0.002 |
| +TME | 89 | 0.735 (0.527–0.897) | +0.004 | 0.738 (0.397–0.979) | +0.006 |
| +Gene | 89 | 0.728 (0.516–0.894) | −0.003 | 0.734 (0.390–0.982) | +0.002 |
| +Stage/Grade+TME | 89 | 0.750 (0.549–0.905) | +0.019 | 0.745 (0.414–0.982) | +0.013 |
| +Stage/Grade+Gene | 89 | 0.747 (0.546–0.903) | +0.017 | 0.745 (0.411–0.979) | +0.013 |
| +TME+Gene | 89 | 0.751 (0.551–0.907) | +0.021 | 0.745 (0.414–0.982) | +0.013 |
| +Stage/Grade +TME+Gene | 89 | 0.776 (0.593–0.919) | +0.045 | 0.762 (0.459–0.982) | +0.030 |

**Supplementary Table 53. Performance of different modality combinations on the TCGA-PAAD dataset for prognostic stratification.** C-index: Concordance Index; AUC: area under the ROC curve at 36 months (Methods); Δ: difference from the pathology-only model (Path). N: number of samples. Values are reported with 95% CIs.

| Model | N | C-index | ΔC-index | AUC | ΔAUC |
|---|---|---|---|---|---|
| UNI | 56 | 0.630 (0.508–0.732) | −0.033 | 0.726 (0.479–0.936) | −0.068 |
| Prov-GigaPath | 56 | 0.646 (0.521–0.767) | −0.017 | 0.731 (0.500–0.931) | −0.064 |
| CHIEF | 56 | 0.566 (0.433–0.700) | −0.097 | 0.739 (0.500–0.967) | −0.056 |
| TITAN | 56 | 0.578 (0.458–0.690) | −0.085 | 0.795 (0.588–0.961) | +0.000 |
| GPFM | 56 | 0.603 (0.468–0.734) | −0.061 | 0.744 (0.523–0.928) | −0.051 |
| *TEAM* | | | | | |
| Path | 56 | 0.663 (0.536–0.782) | +0.000 | 0.795 (0.558–0.971) | +0.000 |
| +Stage/Grade | 56 | 0.696 (0.572–0.811) | +0.032 | 0.812 (0.568–0.978) | +0.017 |
| +TME | 56 | 0.705 (0.578–0.815) | +0.042 | 0.825 (0.579–0.981) | +0.030 |
| +Gene | 56 | 0.697 (0.573–0.810) | +0.033 | 0.825 (0.576–0.982) | +0.030 |
| +Stage/Grade+TME | 56 | 0.729 (0.609–0.838) | +0.066 | 0.850 (0.612–0.991) | +0.056 |
| +Stage/Grade+Gene | 56 | 0.728 (0.609–0.834) | +0.065 | 0.850 (0.612–0.991) | +0.056 |
| +TME+Gene | 56 | 0.736 (0.617–0.840) | +0.073 | 0.846 (0.611–0.990) | +0.051 |
| +Stage/Grade +TME+Gene | 56 | 0.753 (0.636–0.853) | +0.090 | 0.868 (0.660–0.993) | +0.073 |

**Supplementary Table 54. Performance of different modality combinations on the CPTAC-PAAD dataset for prognostic stratification.** C-index: Concordance Index; AUC: area under the ROC curve at 36 months (Methods); Δ: difference from the pathology-only model (Path). N: number of samples. Values are reported with 95% CIs.

| Model | N | C-index | ΔC-index | AUC | ΔAUC |
|---|---|---|---|---|---|
| UNI | 418 | 0.534 (0.491–0.571) | −0.019 | 0.597 (0.509–0.684) | −0.030 |
| Prov-GigaPath | 418 | 0.525 (0.484–0.563) | −0.027 | 0.599 (0.516–0.674) | −0.029 |
| CHIEF | 418 | 0.516 (0.479–0.552) | −0.036 | 0.595 (0.514–0.671) | −0.033 |
| TITAN | 418 | 0.526 (0.482–0.566) | −0.026 | 0.576 (0.491–0.660) | −0.051 |
| GPFM | 418 | 0.506 (0.469–0.543) | −0.046 | 0.565 (0.486–0.642) | −0.063 |
| *TEAM* | | | | | |
| Path | 418 | 0.552 (0.509–0.591) | +0.000 | 0.628 (0.541–0.710) | +0.000 |
| +Stage/Grade | 418 | 0.587 (0.551–0.628) | +0.035 | 0.675 (0.592–0.750) | +0.048 |
| +TME | 418 | 0.604 (0.569–0.645) | +0.051 | 0.702 (0.622–0.774) | +0.075 |
| +Gene | 418 | 0.596 (0.560–0.636) | +0.044 | 0.691 (0.609–0.765) | +0.063 |
| +Stage/Grade+TME | 418 | 0.618 (0.583–0.658) | +0.066 | 0.721 (0.642–0.792) | +0.093 |
| +Stage/Grade+Gene | 418 | 0.613 (0.577–0.654) | +0.061 | 0.716 (0.636–0.788) | +0.088 |
| +TME+Gene | 418 | 0.624 (0.590–0.664) | +0.072 | 0.731 (0.655–0.801) | +0.104 |
| +Stage/Grade +TME+Gene | 418 | 0.645 (0.610–0.683) | +0.093 | 0.760 (0.689–0.825) | +0.132 |

**Supplementary Table 55. Performance of different modality combinations on the TCGA-UCEC dataset for prognostic stratification.** C-index: Concordance Index; AUC: area under the ROC curve at 36 months (Methods); Δ: difference from the pathology-only model (Path). N: number of samples. Values are reported with 95% CIs.

| Model | N | C-index | ΔC-index | AUC | ΔAUC |
|---|---|---|---|---|---|
| UNI | 167 | 0.676 (0.542–0.796) | −0.172 | 0.742 (0.596–0.880) | −0.192 |
| Prov-GigaPath | 167 | 0.665 (0.539–0.780) | −0.184 | 0.719 (0.571–0.863) | −0.215 |
| CHIEF | 167 | 0.691 (0.570–0.793) | −0.157 | 0.777 (0.646–0.887) | −0.157 |
| TITAN | 167 | 0.828 (0.746–0.898) | −0.020 | 0.931 (0.860–0.981) | −0.003 |
| GPFM | 167 | 0.744 (0.639–0.837) | −0.104 | 0.834 (0.720–0.931) | −0.100 |
| *TEAM* | | | | | |
| Path | 167 | 0.848 (0.755–0.918) | +0.000 | 0.934 (0.838–0.995) | +0.000 |
| +Stage/Grade | 167 | 0.830 (0.745–0.906) | −0.018 | 0.879 (0.785–0.950) | −0.055 |
| +TME | 167 | 0.831 (0.738–0.914) | −0.017 | 0.891 (0.802–0.960) | −0.043 |
| +Gene | 167 | 0.852 (0.776–0.920) | +0.004 | 0.906 (0.823–0.965) | −0.028 |
| +Stage/Grade+TME | 167 | 0.860 (0.791–0.925) | +0.012 | 0.925 (0.856–0.975) | −0.009 |
| +Stage/Grade+Gene | 167 | 0.874 (0.811–0.935) | +0.026 | 0.937 (0.869–0.982) | +0.003 |
| +TME+Gene | 167 | 0.876 (0.815–0.937) | +0.028 | 0.937 (0.870–0.982) | +0.003 |
| +Stage/Grade +TME+Gene | 167 | 0.872 (0.807–0.933) | +0.024 | 0.933 (0.863–0.978) | −0.002 |

**Supplementary Table 56. Performance of different modality combinations on the CPTAC-UCEC dataset for prognostic stratification.** C-index: Concordance Index; AUC: area under the ROC curve at 36 months (Methods); Δ: difference from the pathology-only model (Path). N: number of samples. Values are reported with 95% CIs.

| **Model** | **N** | **C-index** | **ΔC-index** | **AUC** | **ΔAUC** |
|---|---|---|---|---|---|
| UNI | 727 | 0.600 (0.536–0.668) | −0.090 | 0.598 (0.501–0.684) | −0.076 |
| Prov-GigaPath | 727 | 0.635 (0.567–0.701) | −0.055 | 0.618 (0.522–0.703) | −0.056 |
| CHIEF | 727 | 0.548 (0.479–0.620) | −0.142 | 0.548 (0.450–0.640) | −0.126 |
| TITAN | 727 | 0.658 (0.590–0.722) | −0.032 | 0.643 (0.552–0.725) | −0.031 |
| GPFM | 727 | 0.623 (0.559–0.689) | −0.067 | 0.608 (0.514–0.691) | −0.066 |
| *TEAM* | | | | | |
| Path | 727 | 0.690 (0.633–0.753) | +0.000 | 0.674 (0.579–0.766) | +0.000 |
| +Stage/Grade | 727 | 0.718 (0.663–0.771) | +0.028 | 0.827 (0.738–0.898) | +0.153 |
| +TME | 727 | 0.713 (0.655–0.768) | +0.023 | 0.887 (0.805–0.955) | +0.213 |
| +Gene | 727 | 0.726 (0.664–0.780) | +0.036 | 0.854 (0.770–0.932) | +0.180 |
| +Stage/Grade+TME | 727 | 0.733 (0.682–0.779) | +0.043 | 0.903 (0.834–0.960) | +0.229 |
| +Stage/Grade+Gene | 727 | 0.749 (0.691–0.802) | +0.059 | 0.889 (0.815–0.955) | +0.215 |
| +TME+Gene | 727 | 0.749 (0.701–0.796) | +0.059 | 0.919 (0.858–0.967) | +0.245 |
| +Stage/Grade +TME+Gene | 727 | 0.733 (0.685–0.781) | +0.043 | 0.927 (0.864–0.975) | +0.253 |

**Supplementary Table 57. Performance of different modality combinations on the Yale HER2+ dataset (internal test set) for therapeutic response prediction.** OR: mean odds ratio for responder enrichment (Methods); AP: Average Precision; Δ: difference from the pathology-only model (Path); N: number of samples. Values are reported with 95% CIs.

| **Model** | **N** | **OR** | **ΔOR** | **AP** | **ΔAP** |
|---|---|---|---|---|---|
| UNI | 26 | 1.06 (0.292–6.42) | −6.1 | 0.499 (0.258–0.764) | −0.269 |
| Prov-GigaPath | 26 | 1.43 (0.461–10.5) | −5.8 | 0.634 (0.394–0.860) | −0.134 |
| CHIEF | 26 | 3.14 (1.07–16.4) | −4.0 | 0.568 (0.357–0.877) | −0.201 |
| TITAN | 26 | 1.72 (0.469–9.60) | −5.5 | 0.542 (0.294–0.804) | −0.227 |
| GPFM | 26 | 3.16 (0.798–14.7) | −4.0 | 0.582 (0.354–0.874) | −0.187 |
| *TEAM* | | | | | |
| Path | 26 | 7.19 (1.79–26.3) | +0.0 | 0.768 (0.546–0.935) | +0.000 |
| +Stage/Grade | 26 | 15.7 (5.82–68.6) | +8.5 | 0.865 (0.661–1.000) | +0.097 |
| +TME | 26 | 15.3 (7.18–56.7) | +8.1 | 0.890 (0.742–0.991) | +0.121 |
| +Gene | 26 | 16.8 (5.05–68.6) | +9.6 | 0.895 (0.728–1.000) | +0.127 |
| +Stage/Grade+TME | 26 | 15.3 (7.98–56.6) | +8.1 | 0.893 (0.730–0.989) | +0.125 |
| +Stage/Grade+Gene | 26 | 15.1 (4.92–68.6) | +7.9 | 0.861 (0.646–1.000) | +0.093 |
| +TME+Gene | 26 | 21.3 (8.81–62.3) | +14.1 | 0.903 (0.759–1.000) | +0.135 |
| +Stage/Grade +TME+Gene | 26 | 53.4 (23.2–132) | +46.2 | 0.981 (0.917–1.000) | +0.212 |

**Supplementary Table 58. Performance of different modality combinations on the IMPRESS HER2+ dataset (external test set) for therapeutic response prediction.** OR: mean odds ratio for responder enrichment (Methods); AP: Average Precision; Δ: difference from the pathology-only model (Path); N: number of samples. Values are reported with 95% CIs.

| **Model** | **N** | **OR** | **ΔOR** | **AP** | **ΔAP** |
|---|---|---|---|---|---|
| UNI | 62 | 1.27 (0.449–3.77) | −6.2 | 0.648 (0.515–0.803) | −0.161 |
| Prov-GigaPath | 62 | 1.10 (0.522–3.47) | −6.4 | 0.649 (0.515–0.817) | −0.161 |
| CHIEF | 62 | 1.12 (0.433–3.52) | −6.4 | 0.631 (0.491–0.784) | −0.178 |
| TITAN | 62 | 1.27 (0.560–3.69) | −6.3 | 0.671 (0.539–0.827) | −0.138 |
| GPFM | 62 | 1.04 (0.492–3.11) | −6.5 | 0.634 (0.505–0.792) | −0.176 |
| *TEAM* | | | | | |
| Path | 62 | 7.52 (1.96–17.7) | +0.0 | 0.809 (0.674–0.920) | +0.000 |
| +Stage/Grade | 62 | 32.6 (15.1–73.6) | +25.1 | 0.948 (0.899–0.986) | +0.139 |
| +TME | 62 | 25.1 (13.2–75.5) | +17.6 | 0.951 (0.897–0.987) | +0.141 |
| +Gene | 62 | 25.9 (11.2–105) | +18.4 | 0.945 (0.870–0.991) | +0.136 |
| +Stage/Grade+TME | 62 | 32.3 (20.8–123) | +24.8 | 0.965 (0.919–0.994) | +0.156 |
| +Stage/Grade+Gene | 62 | 43.6 (25.1–138) | +36.1 | 0.960 (0.908–0.997) | +0.151 |
| +TME+Gene | 62 | 26.3 (11.3–118) | +18.8 | 0.938 (0.857–0.994) | +0.129 |
| +Stage/Grade +TME+Gene | 62 | 46.7 (26.0–181) | +39.2 | 0.965 (0.908–0.998) | +0.155 |

**Supplementary Table 59. Performance of different modality combinations on the TransNEO HER2+ dataset (external test set) for therapeutic response prediction.** OR: mean odds ratio for responder enrichment (Methods); AP: Average Precision; Δ: difference from the pathology-only model (Path); N: number of samples. Values are reported with 95% CIs.

| **Model** | **N** | **OR** | **ΔOR** | **AP** | **ΔAP** |
|---|---|---|---|---|---|
| UNI | 76 | 1.84 (0.695–5.27) | −3.9 | 0.535 (0.363–0.713) | −0.147 |
| Prov-GigaPath | 76 | 2.08 (0.775–5.82) | −3.7 | 0.536 (0.369–0.728) | −0.146 |
| CHIEF | 76 | 1.44 (0.652–5.10) | −4.3 | 0.531 (0.359–0.710) | −0.151 |
| TITAN | 76 | 1.96 (0.768–5.37) | −3.8 | 0.544 (0.369–0.723) | −0.138 |
| GPFM | 76 | 2.05 (0.843–9.01) | −3.7 | 0.578 (0.426–0.762) | −0.103 |
| *TEAM* | | | | | |
| Path | 76 | 5.74 (2.59–16.7) | +0.0 | 0.681 (0.509–0.833) | +0.000 |
| +Stage/Grade | 76 | 9.84 (4.25–53.7) | +4.1 | 0.819 (0.692–0.932) | +0.137 |
| +TME | 76 | 14.9 (6.61–60.1) | +9.1 | 0.757 (0.604–0.923) | +0.076 |
| +Gene | 76 | 19.1 (9.22–65.2) | +13.4 | 0.774 (0.614–0.940) | +0.093 |
| +Stage/Grade+TME | 76 | 16.7 (7.19–76.6) | +10.9 | 0.818 (0.677–0.942) | +0.137 |
| +Stage/Grade+Gene | 76 | 17.4 (7.79–90.1) | +11.7 | 0.838 (0.696–0.958) | +0.157 |
| +TME+Gene | 76 | 15.1 (7.52–69.1) | +9.3 | 0.806 (0.661–0.932) | +0.124 |
| +Stage/Grade +TME+Gene | 76 | 20.5 (9.96–80.3) | +14.8 | 0.846 (0.705–0.952) | +0.164 |

**Supplementary Table 60. Performance of different modality combinations on the BEV-OV dataset for therapeutic response prediction.** OR: mean odds ratio for responder enrichment (Methods); AP: Average Precision; Δ: difference from the pathology-only model (Path); N: number of samples. Values are reported with 95% CIs.

| **Model** | **N** | **OR** | **ΔOR** | **AP** | **ΔAP** |
|---|---|---|---|---|---|
| UNI | 86 | 7.48 (3.48–22.2) | −17.6 | 0.808 (0.688–0.915) | −0.093 |
| Prov-GigaPath | 86 | 13.1 (6.69–51.9) | −11.9 | 0.817 (0.695–0.956) | −0.083 |
| CHIEF | 86 | 10.6 (4.98–39.1) | −14.4 | 0.868 (0.777–0.949) | −0.032 |
| TITAN | 86 | 6.57 (3.16–21.1) | −18.5 | 0.827 (0.713–0.924) | −0.073 |
| GPFM | 86 | 8.12 (3.40–25.6) | −16.9 | 0.790 (0.667–0.910) | −0.111 |
| *TEAM* | | | | | |
| Path | 86 | 25.0 (10.9–109) | +0.0 | 0.900 (0.799–0.984) | +0.000 |
| +Stage/Grade | 86 | 35.4 (18.3–169) | +10.4 | 0.934 (0.855–0.993) | +0.034 |
| +TME | 86 | 43.9 (26.8–169) | +18.8 | 0.922 (0.820–0.995) | +0.022 |
| +Gene | 86 | 43.1 (22.4–163) | +18.1 | 0.933 (0.858–0.993) | +0.033 |
| +Stage/Grade+TME | 86 | 42.2 (18.4–184) | +17.1 | 0.920 (0.824–0.994) | +0.020 |
| +Stage/Grade+Gene | 86 | 41.1 (20.6–155) | +16.1 | 0.928 (0.833–0.993) | +0.028 |
| +TME+Gene | 86 | 39.7 (17.3–176) | +14.7 | 0.920 (0.821–0.995) | +0.019 |
| +Stage/Grade +TME+Gene | 86 | 48.2 (29.5–162) | +23.2 | 0.953 (0.899–0.993) | +0.053 |

**Supplementary Table 61. Performance of different modality combinations on the PDL1-NSCLC dataset for therapeutic response prediction.** OR: mean odds ratio for responder enrichment (Methods); AP: Average Precision; Δ: difference from the pathology-only model (Path). N: number of samples. Values are reported with 95% CIs.

| **Model** | **N** | **OR** | **ΔOR** | **AP** | **ΔAP** |
|---|---|---|---|---|---|
| UNI | 58 | 3.62 (1.54–14.8) | −4.5 | 0.866 (0.750–0.966) | −0.018 |
| Prov-GigaPath | 58 | 1.99 (0.819–9.20) | −6.1 | 0.844 (0.723–0.949) | −0.040 |
| CHIEF | 58 | 3.87 (1.47–16.0) | −4.2 | 0.865 (0.753–0.963) | −0.019 |
| TITAN | 58 | 3.32 (1.30–15.8) | −4.8 | 0.835 (0.700–0.961) | −0.049 |
| GPFM | 58 | 2.32 (1.02–10.9) | −5.8 | 0.864 (0.749–0.955) | −0.020 |
| *TEAM* | | | | | |
| Path | 58 | 8.11 (3.15–43.5) | +0.0 | 0.884 (0.758–0.991) | +0.000 |
| +Stage/Grade | 58 | 18.4 (5.72–85.8) | +10.3 | 0.922 (0.818–0.997) | +0.038 |
| +TME | 58 | 20.3 (5.67–79.6) | +12.2 | 0.938 (0.848–0.997) | +0.054 |
| +Gene | 58 | 19.4 (6.49–89.9) | +11.3 | 0.921 (0.815–0.998) | +0.037 |
| +Stage/Grade+TME | 58 | 14.4 (4.51–71.5) | +6.3 | 0.918 (0.807–0.996) | +0.034 |
| +Stage/Grade+Gene | 58 | 21.0 (7.28–98.6) | +12.9 | 0.926 (0.827–0.999) | +0.042 |
| +TME+Gene | 58 | 20.8 (7.92–93.7) | +12.7 | 0.923 (0.821–0.998) | +0.039 |
| +Stage/Grade +TME+Gene | 58 | 14.9 (5.51–80.5) | +6.8 | 0.922 (0.814–0.997) | +0.038 |

**Supplementary Table 62. Performance of different modality combinations on the CRT-CESC dataset for therapeutic response prediction.** OR: mean odds ratio for responder enrichment (Methods); AP: Average Precision; Δ: difference from the pathology-only model (Path). N: number of samples. Values are reported with 95% CIs.

| Model | N | OR | ΔOR | AP | ΔAP |
|---|---|---|---|---|---|
| UNI | 54 | 25.7 (15.0–63.9) | −14.7 | 0.542 (0.381–0.815) | −0.395 |
| Prov-GigaPath | 54 | 24.8 (6.91–103) | −15.6 | 0.837 (0.654–0.981) | −0.100 |
| CHIEF | 54 | 22.0 (7.14–149) | −18.4 | 0.882 (0.720–1.000) | −0.055 |
| TITAN | 54 | 17.8 (9.17–68.1) | −22.6 | 0.702 (0.487–0.949) | −0.236 |
| GPFM | 54 | 21.3 (6.67–112) | −19.1 | 0.867 (0.692–0.981) | −0.071 |
| *TEAM* | | | | | |
| Path | 54 | 40.4 (12.6–173) | +0.0 | 0.937 (0.825–1.000) | +0.000 |
| +Stage/Grade | 54 | 72.7 (54.0–380) | +32.2 | 0.986 (0.943–1.000) | +0.049 |
| +TME | 54 | 64.5 (18.9–380) | +24.1 | 0.961 (0.873–1.000) | +0.023 |
| +Gene | 54 | 62.1 (21.4–269) | +21.7 | 0.960 (0.868–1.000) | +0.023 |
| +Stage/Grade+TME | 54 | 94.4 (55.9–380) | +53.9 | 0.993 (0.964–1.000) | +0.056 |
| +Stage/Grade+Gene | 54 | 75.7 (31.5–267) | +35.3 | 0.973 (0.907–1.000) | +0.036 |
| +TME+Gene | 54 | 68.6 (19.9–269) | +28.2 | 0.961 (0.876–1.000) | +0.024 |
| +Stage/Grade +TME+Gene | 54 | 94.4 (34.8–267) | +53.9 | 0.973 (0.905–1.000) | +0.035 |

**Supplementary Table 63. Attention weight allocation for the prognostic stratification task.** Average normalized attention weights for each modality source across all prognostic stratification datasets. Higher values indicate greater importance in the prediction.

| Model | Slide | Stage/Grade | TME | Gene |
|---|---|---|---|---|
| Path | 1.000 | – | – | – |
| +Stage/Grade | 0.613 | 0.387 | – | – |
| +TME | 0.614 | – | 0.386 | – |
| +Gene | 0.614 | – | – | 0.386 |
| +Stage/Grade+TME | 0.598 | 0.198 | 0.204 | – |
| +Stage/Grade+Gene | 0.594 | 0.202 | – | 0.204 |
| +TME+Gene | 0.596 | – | 0.208 | 0.197 |
| +Stage/Grade+TME+Gene | 0.620 | 0.123 | 0.133 | 0.123 |

**Supplementary Table 64. Attention weight allocation for the therapeutic response prediction task.** Average normalized attention weights for each modality source across all therapeutic response datasets. Higher values indicate greater importance in the prediction.

| **Model** | **Slide** | **Stage/Grade** | **TME** | **Gene** |
|---|---|---|---|---|
| Path | 1.000 | – | – | – |
| +Stage/Grade | 0.537 | 0.463 | – | – |
| +TME | 0.536 | – | 0.464 | – |
| +Gene | 0.533 | – | – | 0.467 |
| +Stage/Grade+TME | 0.456 | 0.270 | 0.274 | – |
| +Stage/Grade+Gene | 0.459 | 0.261 | – | 0.280 |
| +TME+Gene | 0.449 | – | 0.274 | 0.277 |
| +Stage/Grade+TME+Gene | 0.422 | 0.188 | 0.193 | 0.197 |

**Supplementary Table 65. Performance of different foundation models on the TCGA-BRCA dataset for cancer staging.** Comparison of foundation model encoders on TCGA-BRCA. Metric values are reported with 95% CIs.

| **Model** | **AUC** | **95% CI** |
|---|---|---|
| UNI | 0.904 | 0.885–0.923 |
| Prov-GigaPath | 0.925 | 0.907–0.941 |
| CHIEF | 0.917 | 0.897–0.933 |
| TITAN | 0.934 | 0.917–0.950 |
| GPFM | 0.906 | 0.886–0.925 |
| TEAM | 0.950 | 0.936–0.965 |

**Supplementary Table 66. Performance of different foundation models on the CPTAC-BRCA dataset for cancer staging.** Comparison of foundation model encoders on CPTAC-BRCA. Metric values are reported with 95% CIs.

| **Model** | **AUC** | **95% CI** |
|---|---|---|
| UNI | 0.769 | 0.677–0.855 |
| Prov-GigaPath | 0.732 | 0.635–0.822 |
| CHIEF | 0.669 | 0.584–0.754 |
| TITAN | 0.749 | 0.659–0.826 |
| GPFM | 0.753 | 0.666–0.824 |
| TEAM | 0.797 | 0.733–0.859 |

**Supplementary Table 67. Performance of different foundation models on the NFH-BRCA dataset for cancer staging.** Comparison of foundation model encoders on NFH-BRCA. Metric values are reported with 95% CIs.

| Model | AUC | 95% CI |
|---|---|---|
| UNI | 0.812 | 0.757–0.863 |
| Prov-GigaPath | 0.870 | 0.830–0.906 |
| CHIEF | 0.849 | 0.808–0.887 |
| TITAN | 0.855 | 0.806–0.906 |
| GPFM | 0.873 | 0.837–0.907 |
| TEAM | 0.884 | 0.847–0.922 |

**Supplementary Table 68. Performance of different foundation models on the TCGA-CESC dataset for cancer staging.** Comparison of foundation model encoders on TCGA-CESC. Metric values are reported with 95% CIs.

| Model | AUC | 95% CI |
|---|---|---|
| UNI | 0.967 | 0.937–0.988 |
| Prov-GigaPath | 0.970 | 0.948–0.989 |
| CHIEF | 0.939 | 0.899–0.972 |
| TITAN | 0.989 | 0.974–0.998 |
| GPFM | 0.987 | 0.970–1.000 |
| TEAM | 1.000 | 1.000–1.000 |

**Supplementary Table 69. Performance of different foundation models on the NFH-CESC dataset for cancer staging.** Comparison of foundation model encoders on NFH-CESC. Metric values are reported with 95% CIs.

| Model | AUC | 95% CI |
|---|---|---|
| UNI | 0.793 | 0.740–0.841 |
| Prov-GigaPath | 0.836 | 0.789–0.880 |
| CHIEF | 0.814 | 0.762–0.862 |
| TITAN | 0.813 | 0.764–0.861 |
| GPFM | 0.821 | 0.770–0.866 |
| TEAM | 0.853 | 0.808–0.896 |

**Supplementary Table 70. Performance of different foundation models on the TCGA-CRC dataset for cancer staging.** Comparison of foundation model encoders on TCGA-CRC. Metric values are reported with 95% CIs.

| Model | AUC | 95% CI |
|---|---|---|
| UNI | 0.835 | 0.796–0.870 |
| Prov-GigaPath | 0.865 | 0.833–0.898 |
| CHIEF | 0.848 | 0.807–0.882 |
| TITAN | 0.846 | 0.813–0.881 |
| GPFM | 0.854 | 0.817–0.889 |
| TEAM | 0.883 | 0.846–0.916 |

**Supplementary Table 71. Performance of different foundation models on the CPTAC-CRC dataset for cancer staging.** Comparison of foundation model encoders on CPTAC-CRC. Metric values are reported with 95% CIs.

| Model | AUC | 95% CI |
|---|---|---|
| UNI | 0.800 | 0.735–0.867 |
| Prov-GigaPath | 0.768 | 0.692–0.838 |
| CHIEF | 0.750 | 0.656–0.839 |
| TITAN | 0.767 | 0.694–0.839 |
| GPFM | 0.715 | 0.626–0.794 |
| TEAM | 0.814 | 0.748–0.870 |

**Supplementary Table 72. Performance of different foundation models on the NFH-CRC dataset for cancer staging.** Comparison of foundation model encoders on NFH-CRC. Metric values are reported with 95% CIs.

| Model | AUC | 95% CI |
|---|---|---|
| UNI | 0.583 | 0.524–0.640 |
| Prov-GigaPath | 0.595 | 0.539–0.653 |
| CHIEF | 0.654 | 0.598–0.707 |
| TITAN | 0.649 | 0.597–0.704 |
| GPFM | 0.652 | 0.595–0.703 |
| TEAM | 0.709 | 0.658–0.757 |

**Supplementary Table 73. Performance of different foundation models on the TCGA-HNSC dataset for cancer staging.** Comparison of foundation model encoders on TCGA-HNSC. Metric values are reported with 95% CIs.

| Model | AUC | 95% CI |
|---|---|---|
| UNI | 0.894 | 0.861–0.925 |
| Prov-GigaPath | 0.905 | 0.876–0.933 |
| CHIEF | 0.868 | 0.836–0.901 |
| TITAN | 0.929 | 0.903–0.952 |
| GPFM | 0.838 | 0.798–0.876 |
| TEAM | 0.935 | 0.909–0.961 |

**Supplementary Table 74. Performance of different foundation models on the CPTAC-HNSC dataset for cancer staging.** Comparison of foundation model encoders on CPTAC-HNSC. Metric values are reported with 95% CIs.

| Model | AUC | 95% CI |
|---|---|---|
| UNI | 0.820 | 0.781–0.851 |
| Prov-GigaPath | 0.836 | 0.800–0.871 |
| CHIEF | 0.806 | 0.768–0.847 |
| TITAN | 0.855 | 0.825–0.882 |
| GPFM | 0.826 | 0.789–0.863 |
| TEAM | 0.883 | 0.861–0.904 |

**Supplementary Table 75. Performance of different foundation models on the TCGA-LIHC dataset for cancer staging.** Comparison of foundation model encoders on TCGA-LIHC. Metric values are reported with 95% CIs.

| Model | AUC | 95% CI |
|---|---|---|
| UNI | 0.760 | 0.686–0.829 |
| Prov-GigaPath | 0.785 | 0.716–0.847 |
| CHIEF | 0.766 | 0.703–0.829 |
| TITAN | 0.743 | 0.677–0.815 |
| GPFM | 0.774 | 0.699–0.840 |
| TEAM | 0.810 | 0.746–0.874 |

**Supplementary Table 76. Performance of different foundation models on the ZJH-LIHC dataset for cancer staging.** Comparison of foundation model encoders on ZJH-LIHC. Metric values are reported with 95% CIs.

| Model | AUC | 95% CI |
|---|---|---|
| UNI | 0.798 | 0.692–0.892 |
| Prov-GigaPath | 0.836 | 0.761–0.903 |
| CHIEF | 0.828 | 0.757–0.888 |
| TITAN | 0.855 | 0.775–0.927 |
| GPFM | 0.866 | 0.791–0.933 |
| TEAM | 0.894 | 0.823–0.950 |

**Supplementary Table 77. Performance of different foundation models on the TCGA-OV dataset for cancer staging.** Comparison of foundation model encoders on TCGA-OV. Metric values are reported with 95% CIs.

| Model | AUC | 95% CI |
|---|---|---|
| UNI | 0.985 | 0.954–1.000 |
| Prov-GigaPath | 0.986 | 0.956–1.000 |
| CHIEF | 0.974 | 0.912–1.000 |
| TITAN | 0.972 | 0.926–1.000 |
| GPFM | 0.982 | 0.944–1.000 |
| TEAM | 0.994 | 0.972–1.000 |

**Supplementary Table 78. Performance of different foundation models on the HGSOC-OV dataset for cancer staging.** Comparison of foundation model encoders on HGSOC-OV. Metric values are reported with 95% CIs.

| Model | AUC | 95% CI |
|---|---|---|
| UNI | 0.685 | 0.544–0.823 |
| Prov-GigaPath | 0.672 | 0.554–0.797 |
| CHIEF | 0.693 | 0.591–0.815 |
| TITAN | 0.635 | 0.520–0.731 |
| GPFM | 0.677 | 0.603–0.751 |
| TEAM | 0.771 | 0.717–0.825 |

**Supplementary Table 79. Performance of different foundation models on the SUQH-BRCA dataset for cancer grading.** Comparison of foundation model encoders on SUQH-BRCA. Metric values are reported with 95% CIs.

| Model | AUC | 95% CI |
|---|---|---|
| UNI | 0.902 | 0.885–0.917 |
| Prov-GigaPath | 0.936 | 0.925–0.948 |
| CHIEF | 0.851 | 0.827–0.873 |
| TITAN | 0.945 | 0.933–0.955 |
| GPFM | 0.895 | 0.878–0.911 |
| TEAM | 0.974 | 0.966–0.981 |

**Supplementary Table 80. Performance of different foundation models on the QDUH-BRCA dataset for cancer grading.** Comparison of foundation model encoders on QDUH-BRCA. Metric values are reported with 95% CIs.

| Model | AUC | 95% CI |
|---|---|---|
| UNI | 0.782 | 0.746–0.820 |
| Prov-GigaPath | 0.816 | 0.783–0.850 |
| CHIEF | 0.796 | 0.759–0.829 |
| TITAN | 0.805 | 0.769–0.838 |
| GPFM | 0.796 | 0.759–0.832 |
| TEAM | 0.824 | 0.791–0.855 |

**Supplementary Table 81. Performance of different foundation models on the SHSU-BRCA dataset for cancer grading.** Comparison of foundation model encoders on SHSU-BRCA. Metric values are reported with 95% CIs.

| Model | AUC | 95% CI |
|---|---|---|
| UNI | 0.782 | 0.734–0.828 |
| Prov-GigaPath | 0.749 | 0.701–0.797 |
| CHIEF | 0.682 | 0.629–0.728 |
| TITAN | 0.765 | 0.721–0.807 |
| GPFM | 0.743 | 0.699–0.789 |
| TEAM | 0.803 | 0.762–0.843 |

**Supplementary Table 82. Performance of different foundation models on the TCGA-PRAD dataset for cancer grading.** Comparison of foundation model encoders on TCGA-PRAD. Metric values are reported with 95% CIs.

| Model | AUC | 95% CI |
|---|---|---|
| UNI | 0.926 | 0.898–0.951 |
| Prov-GigaPath | 0.947 | 0.925–0.966 |
| CHIEF | 0.944 | 0.921–0.968 |
| TITAN | 0.952 | 0.929–0.972 |
| GPFM | 0.933 | 0.908–0.958 |
| TEAM | 0.970 | 0.952–0.985 |

**Supplementary Table 83. Performance of different foundation models on the PANDA-PRAD dataset for cancer grading.** Comparison of foundation model encoders on PANDA-PRAD. Metric values are reported with 95% CIs.

| Model | AUC | 95% CI |
|---|---|---|
| UNI | 0.860 | 0.855–0.865 |
| Prov-GigaPath | 0.801 | 0.795–0.807 |
| CHIEF | 0.853 | 0.848–0.859 |
| TITAN | 0.823 | 0.817–0.829 |
| GPFM | 0.864 | 0.859–0.869 |
| TEAM | 0.877 | 0.872–0.881 |

**Supplementary Table 84. Performance of different foundation models on the SICAP-PRAD dataset for cancer grading.** Comparison of foundation model encoders on SICAP-PRAD. Metric values are reported with 95% CIs.

| Model | AUC | 95% CI |
|---|---|---|
| UNI | 0.760 | 0.678–0.826 |
| Prov-GigaPath | 0.745 | 0.667–0.811 |
| CHIEF | 0.774 | 0.718–0.830 |
| TITAN | 0.798 | 0.742–0.852 |
| GPFM | 0.710 | 0.656–0.768 |
| TEAM | 0.832 | 0.784–0.877 |

**Supplementary Table 85. Performance of different foundation models for PD-L1 prediction on internal validation cohorts.** Comparison of foundation model encoders for predicting PD-L1. Pearson correlation coefficient (PCC) values are reported with 95% CIs.

| Model | PCC | 95% CI |
|---|---|---|
| UNI | 0.503 | 0.448–0.551 |
| Prov-GigaPath | 0.475 | 0.415–0.526 |
| CHIEF | 0.559 | 0.511–0.606 |
| TITAN | 0.540 | 0.491–0.588 |
| GPFM | 0.505 | 0.449–0.559 |
| TEAM | 0.591 | 0.544–0.638 |

**Supplementary Table 86. Performance of different foundation models for PD-L1 prediction on external validation cohorts.** Comparison of foundation model encoders for predicting PD-L1. Pearson correlation coefficient (PCC) values are reported with 95% CIs.

| Model | PCC | 95% CI |
|---|---|---|
| UNI | 0.485 | 0.417–0.549 |
| Prov-GigaPath | 0.524 | 0.463–0.587 |
| CHIEF | 0.476 | 0.411–0.540 |
| TITAN | 0.530 | 0.464–0.594 |
| GPFM | 0.486 | 0.424–0.542 |
| TEAM | 0.577 | 0.520–0.633 |

**Supplementary Table 87. Performance of different foundation models for CD8/CD4 ratio prediction on internal validation cohorts.** Comparison of foundation model encoders for predicting CD8/CD4 ratio. Pearson correlation coefficient (PCC) values are reported with 95% CIs.

| Model | PCC | 95% CI |
|---|---|---|
| UNI | 0.316 | 0.241–0.386 |
| Prov-GigaPath | 0.330 | 0.247–0.408 |
| CHIEF | 0.314 | 0.237–0.391 |
| TITAN | 0.355 | 0.268–0.434 |
| GPFM | 0.313 | 0.240–0.385 |
| TEAM | 0.405 | 0.315–0.479 |

**Supplementary Table 88. Performance of different foundation models for CD8/CD4 ratio prediction on external validation cohorts.** Comparison of foundation model encoders for predicting CD8/CD4 ratio. Pearson correlation coefficient (PCC) values are reported with 95% CIs.

| **Model** | **PCC** | **95% CI** |
|---|---|---|
| UNI | 0.485 | 0.423–0.546 |
| Prov-GigaPath | 0.482 | 0.417–0.543 |
| CHIEF | 0.430 | 0.362–0.503 |
| TITAN | 0.469 | 0.398–0.541 |
| GPFM | 0.441 | 0.370–0.506 |
| TEAM | 0.533 | 0.476–0.592 |

**Supplementary Table 89. Performance of different foundation models for TILs prediction on internal validation cohorts.** Comparison of foundation model encoders for predicting TIL density. Pearson correlation coefficient (PCC) values are reported with 95% CIs.

| **Model** | **PCC** | **95% CI** |
|---|---|---|
| UNI | 0.702 | 0.651–0.752 |
| Prov-GigaPath | 0.739 | 0.686–0.787 |
| CHIEF | 0.729 | 0.676–0.777 |
| TITAN | 0.727 | 0.675–0.774 |
| GPFM | 0.696 | 0.646–0.742 |
| TEAM | 0.786 | 0.738–0.829 |

**Supplementary Table 90. Performance of different foundation models for TILs prediction on external validation cohorts.** Comparison of foundation model encoders for predicting TIL density. Pearson correlation coefficient (PCC) values are reported with 95% CIs.

| **Model** | **PCC** | **95% CI** |
|---|---|---|
| UNI | 0.445 | 0.375–0.508 |
| Prov-GigaPath | 0.449 | 0.389–0.511 |
| CHIEF | 0.424 | 0.359–0.487 |
| TITAN | 0.458 | 0.392–0.524 |
| GPFM | 0.417 | 0.352–0.481 |
| TEAM | 0.503 | 0.441–0.564 |

**Supplementary Table 91. Performance of different foundation models for Ki-67 prediction on internal validation cohorts.** Comparison of foundation model encoders for predicting Ki-67. Pearson correlation coefficient (PCC) values are reported with 95% CIs.

| Model | PCC | 95% CI |
|---|---|---|
| UNI | 0.510 | 0.450–0.570 |
| Prov-GigaPath | 0.512 | 0.448–0.572 |
| CHIEF | 0.499 | 0.439–0.561 |
| TITAN | 0.462 | 0.399–0.519 |
| GPFM | 0.507 | 0.444–0.562 |
| TEAM | 0.566 | 0.504–0.627 |

**Supplementary Table 92. Performance of different foundation models for Ki-67 prediction on external validation cohorts.** Comparison of foundation model encoders for predicting Ki-67. Pearson correlation coefficient (PCC) values are reported with 95% CIs.

| Model | PCC | 95% CI |
|---|---|---|
| UNI | 0.510 | 0.447–0.574 |
| Prov-GigaPath | 0.491 | 0.426–0.548 |
| CHIEF | 0.476 | 0.414–0.542 |
| TITAN | 0.492 | 0.426–0.554 |
| GPFM | 0.538 | 0.472–0.594 |
| TEAM | 0.571 | 0.515–0.627 |

**Supplementary Table 93. Performance of different foundation models for TMB prediction on internal validation cohorts.** Comparison of foundation model encoders for predicting TMB. Pearson correlation coefficient (PCC) values are reported with 95% CIs.

| Model | PCC | 95% CI |
|---|---|---|
| UNI | 0.412 | 0.346–0.478 |
| Prov-GigaPath | 0.413 | 0.343–0.481 |
| CHIEF | 0.417 | 0.348–0.485 |
| TITAN | 0.407 | 0.342–0.471 |
| GPFM | 0.383 | 0.308–0.454 |
| TEAM | 0.488 | 0.418–0.545 |

**Supplementary Table 94. Performance of different foundation models for TMB prediction on external validation cohorts.** Comparison of foundation model encoders for predicting TMB. Pearson correlation coefficient (PCC) values are reported with 95% CIs.

| Model | PCC | 95% CI |
|---|---|---|
| UNI | 0.458 | 0.278–0.606 |
| Prov-GigaPath | 0.496 | 0.286–0.656 |
| CHIEF | 0.453 | 0.250–0.609 |
| TITAN | 0.528 | 0.322–0.675 |
| GPFM | 0.472 | 0.276–0.628 |
| TEAM | 0.560 | 0.344–0.709 |

**Supplementary Table 95. Performance of different foundation models for MSI prediction on internal validation cohorts.** Comparison of foundation model encoders for predicting MSI status. Pearson correlation coefficient (PCC) values are reported with 95% CIs.

| Model | PCC | 95% CI |
|---|---|---|
| UNI | 0.471 | 0.409–0.527 |
| Prov-GigaPath | 0.552 | 0.491–0.606 |
| CHIEF | 0.551 | 0.487–0.606 |
| TITAN | 0.535 | 0.474–0.592 |
| GPFM | 0.482 | 0.418–0.539 |
| TEAM | 0.598 | 0.537–0.653 |

**Supplementary Table 96. Performance of different foundation models for MSI prediction on external validation cohorts.** Comparison of foundation model encoders for predicting MSI status. Pearson correlation coefficient (PCC) values are reported with 95% CIs.

| Model | PCC | 95% CI |
|---|---|---|
| UNI | 0.507 | 0.430–0.576 |
| Prov-GigaPath | 0.507 | 0.432–0.577 |
| CHIEF | 0.507 | 0.430–0.572 |
| TITAN | 0.499 | 0.423–0.569 |
| GPFM | 0.512 | 0.440–0.584 |
| TEAM | 0.573 | 0.497–0.636 |

**Supplementary Table 97. Performance of different foundation models for HRD prediction on internal validation cohorts.** Comparison of foundation model encoders for predicting HRD score. Pearson correlation coefficient (PCC) values are reported with 95% CIs.

| Model | PCC | 95% CI |
|---|---|---|
| UNI | 0.515 | 0.453–0.574 |
| Prov-GigaPath | 0.490 | 0.427–0.553 |
| CHIEF | 0.538 | 0.478–0.593 |
| TITAN | 0.511 | 0.455–0.571 |
| GPFM | 0.521 | 0.463–0.574 |
| TEAM | 0.573 | 0.518–0.629 |

**Supplementary Table 98. Performance of different foundation models for HRD prediction on external validation cohorts.** Comparison of foundation model encoders for predicting HRD score. Pearson correlation coefficient (PCC) values are reported with 95% CIs.

| Model | PCC | 95% CI |
|---|---|---|
| UNI | 0.408 | 0.317–0.499 |
| Prov-GigaPath | 0.379 | 0.283–0.476 |
| CHIEF | 0.410 | 0.321–0.499 |
| TITAN | 0.411 | 0.323–0.497 |
| GPFM | 0.401 | 0.302–0.481 |
| TEAM | 0.461 | 0.374–0.550 |

**Supplementary Table 99. Performance of different foundation models for hypoxia score prediction on internal validation cohorts.** Comparison of foundation model encoders for predicting hypoxia score. Pearson correlation coefficient (PCC) values are reported with 95% CIs.

| Model | PCC | 95% CI |
|---|---|---|
| UNI | 0.511 | 0.451–0.568 |
| Prov-GigaPath | 0.562 | 0.494–0.616 |
| CHIEF | 0.506 | 0.443–0.570 |
| TITAN | 0.530 | 0.469–0.589 |
| GPFM | 0.532 | 0.473–0.585 |
| TEAM | 0.611 | 0.554–0.663 |

**Supplementary Table 100. Performance of different foundation models for hypoxia score prediction on external validation cohorts.** Comparison of foundation model encoders for predicting hypoxia score. Pearson correlation coefficient (PCC) values are reported with 95% CIs.

| Model | PCC | 95% CI |
|---|---|---|
| UNI | 0.469 | 0.403–0.537 |
| Prov-GigaPath | 0.492 | 0.418–0.565 |
| CHIEF | 0.486 | 0.424–0.551 |
| TITAN | 0.525 | 0.461–0.584 |
| GPFM | 0.467 | 0.400–0.535 |
| TEAM | 0.574 | 0.512–0.630 |

**Supplementary Table 101. Performance of different models for gene expression prediction on the CPTAC-BRCA dataset.** Pearson correlation coefficient (PCC) values across all genes and top 1,000 prognostically associated genes are reported with 95% CIs.

| Gene Set | Model | PCC | 95% CI |
|---|---|---|---|
| All Genes | DeepPT | 0.191 | 0.187–0.194 |
| | UNI | 0.191 | 0.188–0.195 |
| | Prov-GigaPath | 0.195 | 0.192–0.199 |
| | CHIEF | 0.193 | 0.190–0.197 |
| | TITAN | 0.191 | 0.187–0.195 |
| | GPFM | 0.193 | 0.189–0.197 |
| | TEAM | 0.197 | 0.194–0.201 |
| Top 1,000 Genes | DeepPT | 0.226 | 0.212–0.240 |
| | UNI | 0.198 | 0.183–0.212 |
| | Prov-GigaPath | 0.181 | 0.167–0.196 |
| | CHIEF | 0.198 | 0.184–0.212 |
| | TITAN | 0.198 | 0.183–0.213 |
| | GPFM | 0.198 | 0.183–0.212 |
| | TEAM | 0.260 | 0.244–0.275 |

**Supplementary Table 102. Performance of different models for gene expression prediction on the CPTAC-CRC dataset.** Pearson correlation coefficient (PCC) values across all genes and top 1,000 prognostically associated genes are reported with 95% CIs.

| Gene Set | Model | PCC | 95% CI |
|---|---|---|---|
| All Genes | DeepPT | 0.066 | 0.063–0.069 |
| | UNI | 0.096 | 0.093–0.099 |
| | Prov-GigaPath | 0.091 | 0.088–0.095 |
| | CHIEF | 0.051 | 0.048–0.054 |
| | TITAN | 0.125 | 0.122–0.128 |
| | GPFM | 0.125 | 0.122–0.129 |
| | TEAM | 0.133 | 0.129–0.136 |
| Top 1,000 Genes | DeepPT | 0.059 | 0.048–0.070 |
| | UNI | 0.103 | 0.091–0.114 |
| | Prov-GigaPath | 0.088 | 0.075–0.101 |
| | CHIEF | 0.058 | 0.047–0.069 |
| | TITAN | 0.119 | 0.107–0.132 |
| | GPFM | 0.134 | 0.122–0.145 |
| | TEAM | 0.129 | 0.116–0.142 |

**Supplementary Table 103. Performance of different models for gene expression prediction on the CPTAC-GBM dataset.** Pearson correlation coefficient (PCC) values across all genes and top 1,000 prognostically associated genes are reported with 95% CIs.

| Gene Set | Model | PCC | 95% CI |
|---|---|---|---|
| All Genes | DeepPT | 0.035 | 0.033–0.037 |
| | UNI | 0.056 | 0.053–0.058 |
| | Prov-GigaPath | 0.034 | 0.031–0.036 |
| | CHIEF | 0.062 | 0.060–0.064 |
| | TITAN | 0.023 | 0.021–0.025 |
| | GPFM | 0.029 | 0.027–0.031 |
| | TEAM | 0.086 | 0.084–0.088 |
| Top 1,000 Genes | DeepPT | 0.032 | 0.025–0.039 |
| | UNI | 0.055 | 0.046–0.064 |
| | Prov-GigaPath | 0.038 | 0.028–0.047 |
| | CHIEF | 0.063 | 0.053–0.072 |
| | TITAN | 0.025 | 0.016–0.033 |
| | GPFM | 0.029 | 0.021–0.038 |
| | TEAM | 0.091 | 0.083–0.098 |

**Supplementary Table 104. Performance of different models for gene expression prediction on the CPTAC-HNSC dataset.** Pearson correlation coefficient (PCC) values across all genes and top 1,000 prognostically associated genes are reported with 95% CIs.

| Gene Set | Model | PCC | 95% CI |
|---|---|---|---|
| All Genes | DeepPT | 0.122 | 0.120–0.124 |
| | UNI | 0.124 | 0.122–0.126 |
| | Prov-GigaPath | 0.123 | 0.121–0.125 |
| | CHIEF | 0.125 | 0.123–0.127 |
| | TITAN | 0.124 | 0.122–0.126 |
| | GPFM | 0.124 | 0.122–0.126 |
| | TEAM | 0.126 | 0.124–0.128 |
| Top 1,000 Genes | DeepPT | 0.130 | 0.123–0.137 |
| | UNI | 0.128 | 0.120–0.135 |
| | Prov-GigaPath | 0.123 | 0.116–0.131 |
| | CHIEF | 0.129 | 0.121–0.136 |
| | TITAN | 0.135 | 0.128–0.143 |
| | GPFM | 0.128 | 0.121–0.136 |
| | TEAM | 0.130 | 0.122–0.137 |

**Supplementary Table 105. Performance of different models for gene expression prediction on the CPTAC-KIRC dataset.** Pearson correlation coefficient (PCC) values across all genes and top 1,000 prognostically associated genes are reported with 95% CIs.

| Gene Set | Model | PCC | 95% CI |
|---|---|---|---|
| All Genes | DeepPT | 0.074 | 0.071–0.076 |
| | UNI | 0.053 | 0.051–0.056 |
| | Prov-GigaPath | 0.070 | 0.067–0.072 |
| | CHIEF | 0.064 | 0.062–0.066 |
| | TITAN | 0.070 | 0.068–0.072 |
| | GPFM | 0.057 | 0.055–0.060 |
| | TEAM | 0.070 | 0.067–0.072 |
| Top 1,000 Genes | DeepPT | 0.146 | 0.137–0.155 |
| | UNI | 0.052 | 0.043–0.061 |
| | Prov-GigaPath | 0.067 | 0.058–0.076 |
| | CHIEF | 0.064 | 0.056–0.073 |
| | TITAN | 0.074 | 0.065–0.083 |
| | GPFM | 0.058 | 0.050–0.066 |
| | TEAM | 0.129 | 0.119–0.139 |

**Supplementary Table 106. Performance of different models for gene expression prediction on the CPTAC-LUAD dataset.** Pearson correlation coefficient (PCC) values across all genes and top 1,000 prognostically associated genes are reported with 95% CIs.

| Gene Set | Model | PCC | 95% CI |
|---|---|---|---|
| All Genes | DeepPT | 0.070 | 0.068–0.072 |
| | UNI | 0.069 | 0.067–0.071 |
| | Prov-GigaPath | 0.063 | 0.061–0.065 |
| | CHIEF | 0.057 | 0.055–0.058 |
| | TITAN | 0.077 | 0.075–0.079 |
| | GPFM | 0.076 | 0.074–0.078 |
| | TEAM | 0.078 | 0.076–0.080 |
| Top 1,000 Genes | DeepPT | 0.109 | 0.101–0.117 |
| | UNI | 0.073 | 0.067–0.080 |
| | Prov-GigaPath | 0.061 | 0.054–0.068 |
| | CHIEF | 0.061 | 0.054–0.068 |
| | TITAN | 0.081 | 0.074–0.088 |
| | GPFM | 0.080 | 0.072–0.087 |
| | TEAM | 0.111 | 0.104–0.119 |

**Supplementary Table 107. Performance of different models for gene expression prediction on the TCGA-BLCA dataset.** Pearson correlation coefficient (PCC) values across all genes and top 1,000 prognostically associated genes are reported with 95% CIs.

| Gene Set | Model | PCC | 95% CI |
|---|---|---|---|
| All Genes | DeepPT | 0.120 | 0.118–0.122 |
| | UNI | 0.102 | 0.100–0.104 |
| | Prov-GigaPath | 0.171 | 0.168–0.173 |
| | CHIEF | 0.238 | 0.236–0.240 |
| | TITAN | 0.166 | 0.163–0.168 |
| | GPFM | 0.096 | 0.094–0.098 |
| | TEAM | 0.241 | 0.238–0.243 |
| Top 1,000 Genes | DeepPT | 0.126 | 0.119–0.132 |
| | UNI | 0.108 | 0.099–0.116 |
| | Prov-GigaPath | 0.173 | 0.164–0.182 |
| | CHIEF | 0.250 | 0.242–0.259 |
| | TITAN | 0.174 | 0.165–0.182 |
| | GPFM | 0.097 | 0.089–0.105 |
| | TEAM | 0.252 | 0.243–0.261 |

**Supplementary Table 108. Performance of different models for gene expression prediction on the TCGA-BRCA dataset.** Pearson correlation coefficient (PCC) values across all genes and top 1,000 prognostically associated genes are reported with 95% CIs.

| Gene Set | Model | PCC | 95% CI |
|---|---|---|---|
| All Genes | DeepPT | 0.189 | 0.188–0.190 |
| | UNI | 0.468 | 0.466–0.470 |
| | Prov-GigaPath | 0.193 | 0.192–0.195 |
| | CHIEF | 0.121 | 0.119–0.122 |
| | TITAN | 0.139 | 0.137–0.140 |
| | GPFM | 0.105 | 0.104–0.107 |
| | TEAM | 0.482 | 0.481–0.484 |
| Top 1,000 Genes | DeepPT | 0.210 | 0.207–0.214 |
| | UNI | 0.471 | 0.465–0.477 |
| | Prov-GigaPath | 0.193 | 0.187–0.199 |
| | CHIEF | 0.124 | 0.118–0.130 |
| | TITAN | 0.140 | 0.134–0.146 |
| | GPFM | 0.108 | 0.103–0.114 |
| | TEAM | 0.517 | 0.512–0.522 |

**Supplementary Table 109. Performance of different models for gene expression prediction on the TCGA-CESC dataset.** Pearson correlation coefficient (PCC) values across all genes and top 1,000 prognostically associated genes are reported with 95% CIs.

| Gene Set | Model | PCC | 95% CI |
|---|---|---|---|
| All Genes | DeepPT | 0.123 | 0.121–0.126 |
| | UNI | 0.054 | 0.052–0.056 |
| | Prov-GigaPath | 0.186 | 0.184–0.188 |
| | CHIEF | 0.058 | 0.056–0.060 |
| | TITAN | 0.094 | 0.092–0.096 |
| | GPFM | 0.252 | 0.250–0.254 |
| | TEAM | 0.328 | 0.326–0.330 |
| Top 1,000 Genes | DeepPT | 0.133 | 0.125–0.141 |
| | UNI | 0.056 | 0.049–0.064 |
| | Prov-GigaPath | 0.187 | 0.179–0.194 |
| | CHIEF | 0.060 | 0.052–0.068 |
| | TITAN | 0.091 | 0.083–0.099 |
| | GPFM | 0.246 | 0.238–0.254 |
| | TEAM | 0.346 | 0.338–0.354 |

**Supplementary Table 110. Performance of different models for gene expression prediction on the TCGA-CRC dataset.** Pearson correlation coefficient (PCC) values across all genes and top 1,000 prognostically associated genes are reported with 95% CIs.

| Gene Set | Model | PCC | 95% CI |
|---|---|---|---|
| All Genes | DeepPT | 0.138 | 0.137–0.140 |
| | UNI | 0.130 | 0.128–0.132 |
| | Prov-GigaPath | 0.194 | 0.192–0.195 |
| | CHIEF | 0.201 | 0.199–0.203 |
| | TITAN | 0.110 | 0.108–0.112 |
| | GPFM | 0.126 | 0.124–0.128 |
| | TEAM | 0.218 | 0.216–0.220 |
| Top 1,000 Genes | DeepPT | 0.115 | 0.109–0.121 |
| | UNI | 0.133 | 0.126–0.140 |
| | Prov-GigaPath | 0.193 | 0.186–0.200 |
| | CHIEF | 0.207 | 0.200–0.214 |
| | TITAN | 0.108 | 0.101–0.115 |
| | GPFM | 0.123 | 0.116–0.130 |
| | TEAM | 0.199 | 0.193–0.206 |

**Supplementary Table 111. Performance of different models for gene expression prediction on the TCGA-GBM dataset.** Pearson correlation coefficient (PCC) values across all genes and top 1,000 prognostically associated genes are reported with 95% CIs.

| Gene Set | Model | PCC | 95% CI |
|---|---|---|---|
| All Genes | DeepPT | −0.019 | (−0.022)–(−0.017) |
| | UNI | −0.085 | (−0.088)–(−0.082) |
| | Prov-GigaPath | −0.059 | (−0.062)–(−0.056) |
| | CHIEF | 0.105 | 0.103–0.108 |
| | TITAN | 0.061 | 0.059–0.064 |
| | GPFM | 0.101 | 0.098–0.104 |
| | TEAM | 0.170 | 0.167–0.173 |
| Top 1,000 Genes | DeepPT | −0.014 | (−0.024)–(−0.003) |
| | UNI | −0.073 | (−0.084)–(−0.063) |
| | Prov-GigaPath | −0.062 | (−0.073)–(−0.052) |
| | CHIEF | 0.114 | 0.103–0.124 |
| | TITAN | 0.060 | 0.050–0.071 |
| | GPFM | 0.105 | 0.093–0.116 |
| | TEAM | 0.198 | 0.187–0.210 |

**Supplementary Table 112. Performance of different models for gene expression prediction on the TCGA-HNSC dataset.** Pearson correlation coefficient (PCC) values across all genes and top 1,000 prognostically associated genes are reported with 95% CIs.

| Gene Set | Model | PCC | 95% CI |
|---|---|---|---|
| All Genes | DeepPT | 0.205 | 0.203–0.207 |
| | UNI | 0.210 | 0.209–0.212 |
| | Prov-GigaPath | 0.246 | 0.244–0.247 |
| | CHIEF | 0.177 | 0.176–0.179 |
| | TITAN | 0.251 | 0.249–0.252 |
| | GPFM | 0.241 | 0.239–0.242 |
| | TEAM | 0.284 | 0.282–0.285 |
| Top 1,000 Genes | DeepPT | 0.220 | 0.213–0.227 |
| | UNI | 0.216 | 0.210–0.222 |
| | Prov-GigaPath | 0.247 | 0.241–0.254 |
| | CHIEF | 0.181 | 0.175–0.187 |
| | TITAN | 0.254 | 0.248–0.260 |
| | GPFM | 0.246 | 0.240–0.252 |
| | TEAM | 0.277 | 0.271–0.284 |

**Supplementary Table 113. Performance of different models for gene expression prediction on the TCGA-KIRC dataset.** Pearson correlation coefficient (PCC) values across all genes and top 1,000 prognostically associated genes are reported with 95% CIs.

| Gene Set | Model | PCC | 95% CI |
|---|---|---|---|
| All Genes | DeepPT | 0.072 | 0.071–0.074 |
| | UNI | 0.024 | 0.022–0.025 |
| | Prov-GigaPath | 0.103 | 0.102–0.105 |
| | CHIEF | 0.040 | 0.038–0.041 |
| | TITAN | 0.281 | 0.280–0.283 |
| | GPFM | 0.031 | 0.030–0.033 |
| | TEAM | 0.303 | 0.302–0.305 |
| Top 1,000 Genes | DeepPT | 0.073 | 0.068–0.079 |
| | UNI | 0.028 | 0.023–0.033 |
| | Prov-GigaPath | 0.102 | 0.096–0.107 |
| | CHIEF | 0.043 | 0.038–0.048 |
| | TITAN | 0.283 | 0.277–0.288 |
| | GPFM | 0.029 | 0.023–0.034 |
| | TEAM | 0.301 | 0.297–0.305 |

**Supplementary Table 114. Performance of different models for gene expression prediction on the TCGA-LIHC dataset.** Pearson correlation coefficient (PCC) values across all genes and top 1,000 prognostically associated genes are reported with 95% CIs.

| Gene Set | Model | PCC | 95% CI |
|---|---|---|---|
| All Genes | DeepPT | 0.152 | 0.150–0.154 |
| | UNI | 0.207 | 0.206–0.208 |
| | Prov-GigaPath | 0.320 | 0.319–0.322 |
| | CHIEF | 0.297 | 0.295–0.298 |
| | TITAN | 0.184 | 0.183–0.186 |
| | GPFM | 0.215 | 0.214–0.217 |
| | TEAM | 0.338 | 0.337–0.340 |
| Top 1,000 Genes | DeepPT | 0.158 | 0.151–0.166 |
| | UNI | 0.209 | 0.204–0.214 |
| | Prov-GigaPath | 0.319 | 0.313–0.324 |
| | CHIEF | 0.300 | 0.294–0.306 |
| | TITAN | 0.181 | 0.176–0.187 |
| | GPFM | 0.211 | 0.206–0.217 |
| | TEAM | 0.358 | 0.353–0.363 |

**Supplementary Table 115. Performance of different models for gene expression prediction on the TCGA-LUAD dataset.** Pearson correlation coefficient (PCC) values across all genes and top 1,000 prognostically associated genes are reported with 95% CIs.

| Gene Set | Model | PCC | 95% CI |
|---|---|---|---|
| All Genes | DeepPT | 0.125 | 0.124–0.127 |
| | UNI | 0.222 | 0.221–0.223 |
| | Prov-GigaPath | 0.200 | 0.199–0.201 |
| | CHIEF | 0.153 | 0.152–0.154 |
| | TITAN | 0.127 | 0.126–0.128 |
| | GPFM | 0.194 | 0.193–0.195 |
| | TEAM | 0.260 | 0.259–0.261 |
| Top 1,000 Genes | DeepPT | 0.134 | 0.128–0.139 |
| | UNI | 0.226 | 0.222–0.230 |
| | Prov-GigaPath | 0.200 | 0.195–0.204 |
| | CHIEF | 0.155 | 0.150–0.160 |
| | TITAN | 0.127 | 0.123–0.131 |
| | GPFM | 0.195 | 0.190–0.199 |
| | TEAM | 0.270 | 0.265–0.274 |

**Supplementary Table 116. Performance of different models for gene expression prediction on the TCGA-PRAD dataset.** Pearson correlation coefficient (PCC) values across all genes and top 1,000 prognostically associated genes are reported with 95% CIs.

| Gene Set | Model | PCC | 95% CI |
|---|---|---|---|
| All Genes | DeepPT | 0.019 | 0.017–0.021 |
| | UNI | −0.043 | (−0.045)–(−0.041) |
| | Prov-GigaPath | 0.079 | 0.077–0.081 |
| | CHIEF | 0.247 | 0.245–0.249 |
| | TITAN | 0.206 | 0.204–0.208 |
| | GPFM | −0.056 | (−0.058)–(−0.054) |
| | TEAM | 0.277 | 0.275–0.279 |
| Top 1,000 Genes | DeepPT | 0.027 | 0.016–0.037 |
| | UNI | −0.041 | (−0.049)–(−0.033) |
| | Prov-GigaPath | 0.078 | 0.070–0.086 |
| | CHIEF | 0.251 | 0.244–0.259 |
| | TITAN | 0.206 | 0.198–0.214 |
| | GPFM | −0.053 | (−0.060)–(−0.046) |
| | TEAM | 0.254 | 0.247–0.261 |

**Supplementary Table 117. Performance of different models for gene expression prediction on the TCGA-STAD dataset.** Pearson correlation coefficient (PCC) values across all genes and top 1,000 prognostically associated genes are reported with 95% CIs.

| Gene Set | Model | PCC | 95% CI |
|---|---|---|---|
| All Genes | DeepPT | 0.081 | 0.079–0.082 |
| | UNI | 0.154 | 0.153–0.156 |
| | Prov-GigaPath | 0.246 | 0.245–0.247 |
| | CHIEF | 0.130 | 0.129–0.131 |
| | TITAN | 0.288 | 0.287–0.290 |
| | GPFM | 0.267 | 0.266–0.268 |
| | TEAM | 0.377 | 0.376–0.378 |
| Top 1,000 Genes | DeepPT | 0.062 | 0.056–0.068 |
| | UNI | 0.156 | 0.151–0.160 |
| | Prov-GigaPath | 0.248 | 0.243–0.252 |
| | CHIEF | 0.136 | 0.131–0.141 |
| | TITAN | 0.289 | 0.284–0.293 |
| | GPFM | 0.266 | 0.261–0.271 |
| | TEAM | 0.378 | 0.374–0.381 |

**Supplementary Table 118. Cohort-level gene expression comparison under a common regression head.** Cells, mean gene-wise PCC for all genes / the top 1,000 prognostically associated genes (15,717 genes for TCGA and the genes shared with TCGA for CPTAC).

| Cohort | UNI | Prov-GigaPath | CHIEF | TITAN | GPFM | DeepPT | TEAM |
|---|---|---|---|---|---|---|---|
| CPTAC-BRCA | 0.191 / 0.198 | 0.195 / 0.181 | 0.193 / 0.198 | 0.191 / 0.198 | 0.193 / 0.198 | 0.191 / 0.226 | 0.197 / 0.260 |
| CPTAC-CRC | 0.096 / 0.103 | 0.091 / 0.088 | 0.051 / 0.058 | 0.125 / 0.119 | 0.125 / 0.134 | 0.066 / 0.059 | 0.133 / 0.129 |
| CPTAC-GBM | 0.056 / 0.055 | 0.034 / 0.038 | 0.062 / 0.063 | 0.023 / 0.025 | 0.029 / 0.029 | 0.035 / 0.032 | 0.086 / 0.091 |
| CPTAC-HNSC | 0.124 / 0.128 | 0.123 / 0.123 | 0.125 / 0.129 | 0.124 / 0.135 | 0.124 / 0.128 | 0.122 / 0.130 | 0.126 / 0.130 |
| CPTAC-KIRC | 0.053 / 0.052 | 0.070 / 0.067 | 0.064 / 0.064 | 0.070 / 0.074 | 0.057 / 0.058 | 0.074 / 0.146 | 0.070 / 0.129 |
| CPTAC-LUAD | 0.069 / 0.073 | 0.063 / 0.061 | 0.057 / 0.061 | 0.077 / 0.081 | 0.076 / 0.080 | 0.070 / 0.109 | 0.078 / 0.111 |
| TCGA-BLCA | 0.102 / 0.108 | 0.171 / 0.173 | 0.238 / 0.250 | 0.166 / 0.174 | 0.096 / 0.097 | 0.120 / 0.126 | 0.241 / 0.252 |
| TCGA-BRCA | 0.468 / 0.471 | 0.193 / 0.193 | 0.121 / 0.124 | 0.139 / 0.140 | 0.105 / 0.108 | 0.189 / 0.210 | 0.482 / 0.517 |
| TCGA-CESC | 0.054 / 0.056 | 0.186 / 0.187 | 0.058 / 0.060 | 0.094 / 0.091 | 0.252 / 0.246 | 0.123 / 0.133 | 0.328 / 0.346 |
| TCGA-CRC | 0.130 / 0.133 | 0.194 / 0.193 | 0.201 / 0.207 | 0.110 / 0.108 | 0.126 / 0.123 | 0.138 / 0.115 | 0.218 / 0.199 |
| TCGA-GBM | −0.085/−0.073 | −0.059/−0.062 | 0.105 / 0.114 | 0.061 / 0.060 | 0.101 / 0.105 | −0.019/−0.014 | 0.170 / 0.198 |
| TCGA-HNSC | 0.210 / 0.216 | 0.246 / 0.247 | 0.177 / 0.181 | 0.251 / 0.254 | 0.241 / 0.246 | 0.205 / 0.220 | 0.284 / 0.277 |
| TCGA-KIRC | 0.024 / 0.028 | 0.103 / 0.102 | 0.040 / 0.043 | 0.281 / 0.283 | 0.031 / 0.029 | 0.072 / 0.073 | 0.303 / 0.301 |
| TCGA-LIHC | 0.207 / 0.209 | 0.320 / 0.319 | 0.297 / 0.300 | 0.184 / 0.181 | 0.215 / 0.211 | 0.152 / 0.158 | 0.338 / 0.358 |
| TCGA-LUAD | 0.222 / 0.226 | 0.200 / 0.200 | 0.153 / 0.155 | 0.127 / 0.127 | 0.194 / 0.195 | 0.125 / 0.134 | 0.260 / 0.270 |
| TCGA-PRAD | −0.043/−0.041 | 0.079 / 0.078 | 0.247 / 0.251 | 0.206 / 0.206 | −0.056/−0.053 | 0.019 / 0.027 | 0.277 / 0.254 |
| TCGA-STAD | 0.154 / 0.156 | 0.246 / 0.248 | 0.130 / 0.136 | 0.288 / 0.289 | 0.267 / 0.266 | 0.081 / 0.062 | 0.377 / 0.378 |

**Supplementary Table 119. Model comparison at matched coverage across downstream tasks.** Mean metric across evaluation units with 95% CIs from 1,000 unit-level bootstrap resamples, on all cases, on the identical population (all models evaluated on the cases retained by TEAM*) and after model-specific case selection (each baseline excludes its own most unstable cases, ranked by prediction instability (Methods), until it retains the same number of cases as TEAM*). Coverage, mean proportion of cases retained by TEAM*; superscripts, number of evaluable units when fewer than the task total.

| Model | All cases | Identical population | Model-specific selection |
|---|---|---|---|
| *Prognostic stratification (31 cohorts; C-index); TEAM* coverage 74.0%* | | | |
| UNI | 0.622 (0.607–0.637) | 0.692 (0.671–0.716) | 0.611 (0.585–0.633) |
| Prov-GigaPath | 0.636 (0.617–0.655) | 0.707 (0.684–0.733) | 0.614 (0.583–0.642) |
| CHIEF | 0.626 (0.608–0.644) | 0.695 (0.665–0.724) | 0.613 (0.580–0.638) |
| TITAN | 0.645 (0.620–0.668) | 0.714 (0.679–0.746) | 0.632 (0.604–0.659) |
| GPFM | 0.638 (0.615–0.661) | 0.712 (0.683–0.742) | 0.627 (0.598–0.656) |
| TEAM | 0.701 (0.678–0.724) | 0.880 (0.863–0.898) | 0.880 (0.863–0.898) |
| *Therapeutic response prediction (6 cohorts; Log OR); TEAM* coverage 66.4%* | | | |
| UNI | 1.24 (0.42–2.18) | 1.46 (0.58–2.29) | 1.24 (0.38–2.20) |
| Prov-GigaPath | 1.28 (0.39–2.21) | 1.53 (0.60–2.41) | 1.12 (0.14–2.21) |
| CHIEF | 1.41 (0.62–2.23) | 1.48 (0.62–2.19) | 1.19 (0.43–2.00) |
| TITAN | 1.24 (0.57–1.99) | 1.29 (0.56–2.07) | 1.51 (0.65–2.49) |

*Continued on next page*

| Model | All cases | Identical population | Model-specific selection |
|---|---|---|---|
| GPFM | 1.32 (0.59–2.10) | 1.88 (1.11–2.46) | 1.29 (0.41–2.09) |
| TEAM | 2.46 (1.95–3.05) | 3.71 (2.80–4.65) | 3.71 (2.80–4.65) |
| *Therapeutic response prediction (6 cohorts; AP); TEAM* coverage 66.4%* | | | |
| UNI | 0.650 (0.539–0.759) | 0.690 (0.601–0.792) | 0.693 (0.609–0.777) |
| Prov-GigaPath | 0.719 (0.622–0.806) | 0.735 (0.648–0.816) | 0.723 (0.609–0.806) |
| CHIEF | 0.724 (0.609–0.832) | 0.742 (0.641–0.841) | 0.677 (0.538–0.805) |
| TITAN | 0.687 (0.591–0.782) | 0.735 (0.667–0.803) | 0.702 (0.570–0.816) |
| GPFM | 0.719 (0.624–0.816) | 0.773 (0.697–0.845) | 0.683 (0.552–0.800) |
| TEAM | 0.830 (0.758–0.892) | 0.930 (0.859–0.989) | 0.930 (0.859–0.989) |
| *Cancer staging/grading (20 cohorts; AUC); TEAM* coverage 62.1%* | | | |
| UNI | 0.821 (0.781–0.862) | 0.830 (0.792–0.868) | 0.826 (0.786–0.867) |
| Prov-GigaPath | 0.829 (0.786–0.873) | 0.835 (0.790–0.877) | 0.836 (0.794–0.876)[18] |
| CHIEF | 0.814 (0.774–0.855) | 0.813 (0.769–0.858) | 0.818 (0.775–0.860)[19] |
| TITAN | 0.834 (0.793–0.877) | 0.820 (0.772–0.869) | 0.835 (0.788–0.881) |
| GPFM | 0.823 (0.785–0.866) | 0.831 (0.792–0.870) | 0.832 (0.794–0.872) |
| TEAM | 0.873 (0.840–0.907) | 0.907 (0.878–0.935) | 0.907 (0.878–0.935) |
| *TME characterization (16 marker–setting evaluations; PCC); TEAM* coverage 60.1%* | | | |
| UNI | 0.482 (0.447–0.520) | 0.560 (0.505–0.617) | 0.488 (0.454–0.531) |
| Prov-GigaPath | 0.493 (0.453–0.539) | 0.566 (0.502–0.625) | 0.504 (0.458–0.550) |
| CHIEF | 0.486 (0.446–0.531) | 0.557 (0.494–0.621) | 0.476 (0.432–0.526) |
| TITAN | 0.499 (0.461–0.541) | 0.568 (0.511–0.625) | 0.508 (0.466–0.549) |
| GPFM | 0.479 (0.442–0.520) | 0.549 (0.491–0.607) | 0.480 (0.441–0.523) |
| TEAM | 0.561 (0.523–0.603) | 0.622 (0.569–0.671) | 0.622 (0.569–0.671) |
| *Gene expression prediction (17 cohorts; sample-wise PCC); TEAM* coverage 53.4%* | | | |
| UNI | 0.818 (0.780–0.852) | 0.824 (0.785–0.858) | 0.824 (0.787–0.859) |
| Prov-GigaPath | 0.757 (0.720–0.791) | 0.765 (0.726–0.798) | 0.766 (0.729–0.800) |
| CHIEF | 0.772 (0.735–0.806) | 0.779 (0.741–0.814) | 0.781 (0.742–0.815) |
| TITAN | 0.788 (0.750–0.822) | 0.796 (0.757–0.830) | 0.796 (0.758–0.830) |
| GPFM | 0.801 (0.764–0.835) | 0.809 (0.770–0.844) | 0.809 (0.772–0.843) |
| DeepPT | 0.743 (0.705–0.776) | 0.749 (0.712–0.783) | 0.750 (0.712–0.784) |
| TEAM | 0.865 (0.828–0.899) | 0.883 (0.845–0.917) | 0.883 (0.845–0.917) |

**Supplementary Table 120. TEAM on all cases and on the cases retained by TEAM*, by cohort and configuration.** Cell: metric on all cases / on retained cases (proportion retained); C-index for the 31 prognostic cohorts, odds ratio (OR) for the 6 therapeutic cohorts, as plotted in Fig. 4e,f.

| Cohort | Path | +Stage/ Grade | +TME | +Gene | +Stage/ Grade +TME | +Stage/ Grade +Gene | +TME +Gene | +Stage/ Grade +TME +Gene |
|---|---|---|---|---|---|---|---|---|
| **Prognostic stratification (C-index; 31 cohorts)** | | | | | | | | |
| TCGA-BLCA | 0.747 / 0.833 (88%) | 0.810 / 0.871 (94%) | 0.819 / 0.869 (94%) | 0.811 / 0.863 (94%) | 0.867 / 0.911 (94%) | 0.869 / 0.913 (94%) | 0.869 / 0.913 (94%) | 0.868 / 0.912 (94%) |
| QDPH-BLCA | 0.639 / 0.921 (71%) | 0.676 / 0.730 (70%) | 0.685 / 0.735 (70%) | 0.683 / 0.698 (70%) | 0.701 / 0.728 (70%) | 0.710 / 0.721 (70%) | 0.695 / 0.729 (70%) | 0.693 / 0.888 (70%) |
| TCGA-BRCA | 0.787 / 0.890 (73%) | 0.860 / 0.885 (97%) | 0.880 / 0.902 (98%) | 0.862 / 0.888 (98%) | 0.874 / 0.903 (98%) | 0.873 / 0.902 (98%) | 0.872 / 0.904 (98%) | 0.874 / 0.903 (98%) |
| NFH-BRCA | 0.679 / 0.986 (61%) | 0.728 / 0.759 (80%) | 0.763 / 0.861 (80%) | 0.742 / 0.792 (79%) | 0.777 / 0.848 (80%) | 0.770 / 0.825 (81%) | 0.766 / 0.795 (80%) | 0.783 / 0.910 (80%) |
| TCGA-CESC | 0.804 / 0.893 (71%) | 0.865 / 0.947 (82%) | 0.875 / 0.957 (79%) | 0.871 / 0.940 (79%) | 0.887 / 0.968 (80%) | 0.883 / 0.958 (82%) | 0.879 / 0.954 (79%) | 0.892 / 0.968 (80%) |
| NFH-CESC | 0.712 / 0.885 (77%) | 0.739 / 0.757 (83%) | 0.756 / 0.785 (83%) | 0.766 / 0.797 (83%) | 0.777 / 0.811 (83%) | 0.766 / 0.796 (83%) | 0.770 / 0.791 (83%) | 0.796 / 0.879 (83%) |
| TCGA-CRC | 0.792 / 0.904 (80%) | 0.836 / 0.869 (91%) | 0.837 / 0.867 (93%) | 0.841 / 0.873 (92%) | 0.876 / 0.937 (92%) | 0.875 / 0.937 (92%) | 0.874 / 0.936 (93%) | 0.874 / 0.936 (92%) |
| APH-CRC | 0.693 / 0.919 (75%) | 0.730 / 0.786 (81%) | 0.741 / 0.770 (82%) | 0.734 / 0.772 (81%) | 0.727 / 0.778 (81%) | 0.752 / 0.777 (81%) | 0.746 / 0.779 (82%) | 0.760 / 0.894 (82%) |
| GDPH-CRC | 0.669 / 0.958 (38%) | 0.703 / 0.653 (72%) | 0.720 / 0.693 (73%) | 0.722 / 0.700 (68%) | 0.742 / 0.770 (72%) | 0.739 / 0.670 (67%) | 0.713 / 0.670 (68%) | 0.744 / 0.917 (72%) |
| HBFH-CRC | 0.673 / 0.876 (71%) | 0.735 / 0.753 (92%) | 0.746 / 0.757 (92%) | 0.738 / 0.726 (90%) | 0.758 / 0.763 (92%) | 0.753 / 0.757 (90%) | 0.750 / 0.765 (92%) | 0.792 / 0.909 (92%) |
| NFH-CRC | 0.688 / 0.896 (66%) | 0.720 / 0.748 (76%) | 0.734 / 0.766 (75%) | 0.722 / 0.769 (76%) | 0.723 / 0.748 (77%) | 0.752 / 0.767 (76%) | 0.727 / 0.756 (76%) | 0.740 / 0.903 (76%) |
| SYSH-CRC | 0.687 / 0.933 (79%) | 0.731 / 0.741 (94%) | 0.797 / 0.788 (94%) | 0.780 / 0.776 (94%) | 0.813 / 0.800 (94%) | 0.808 / 0.806 (94%) | 0.802 / 0.818 (94%) | 0.824 / 0.865 (94%) |
| TCGA-GBM | 0.689 / 0.789 (85%) | 0.721 / 0.755 (95%) | 0.734 / 0.773 (94%) | 0.725 / 0.761 (95%) | 0.761 / 0.798 (94%) | 0.754 / 0.792 (94%) | 0.769 / 0.803 (95%) | 0.786 / 0.816 (95%) |
| CPTAC-GBM | 0.612 / 0.777 (74%) | 0.642 / 0.726 (86%) | 0.656 / 0.741 (86%) | 0.647 / 0.728 (86%) | 0.688 / 0.771 (86%) | 0.678 / 0.762 (86%) | 0.696 / 0.783 (85%) | 0.716 / 0.797 (86%) |
| TCGA-HNSC | 0.764 / 0.836 (91%) | 0.817 / 0.831 (96%) | 0.830 / 0.840 (96%) | 0.820 / 0.831 (96%) | 0.841 / 0.872 (96%) | 0.838 / 0.868 (96%) | 0.834 / 0.865 (96%) | 0.845 / 0.875 (96%) |
| CPTAC-HNSC | 0.648 / 0.851 (80%) | 0.682 / 0.691 (77%) | 0.673 / 0.660 (77%) | 0.678 / 0.721 (78%) | 0.686 / 0.672 (77%) | 0.686 / 0.708 (78%) | 0.691 / 0.713 (78%) | 0.705 / 0.848 (77%) |
| QDPH-HNSC | 0.679 / 0.864 (76%) | 0.700 / 0.725 (84%) | 0.719 / 0.722 (85%) | 0.725 / 0.726 (86%) | 0.719 / 0.738 (86%) | 0.713 / 0.711 (85%) | 0.746 / 0.760 (86%) | 0.746 / 0.823 (86%) |
| TCGA-KIRC | 0.688 / 0.846 (75%) | 0.723 / 0.832 (90%) | 0.733 / 0.836 (90%) | 0.729 / 0.830 (90%) | 0.740 / 0.842 (90%) | 0.738 / 0.841 (90%) | 0.736 / 0.838 (90%) | 0.745 / 0.842 (90%) |
| CPTAC-KIRC | 0.661 / 0.901 (68%) | 0.693 / 0.722 (74%) | 0.681 / 0.719 (73%) | 0.695 / 0.724 (72%) | 0.709 / 0.745 (74%) | 0.701 / 0.739 (73%) | 0.733 / 0.781 (73%) | 0.729 / 0.870 (72%) |
| TCGA-LIHC | 0.797 / 0.856 (84%) | 0.868 / 0.885 (95%) | 0.884 / 0.901 (95%) | 0.876 / 0.895 (95%) | 0.882 / 0.901 (95%) | 0.883 / 0.904 (95%) | 0.890 / 0.910 (95%) | 0.886 / 0.906 (95%) |
| ZJH-LIHC | 0.683 / 0.903 (69%) | 0.703 / 0.773 (71%) | 0.725 / 0.773 (72%) | 0.717 / 0.832 (72%) | 0.734 / 0.835 (72%) | 0.745 / 0.829 (71%) | 0.722 / 0.725 (72%) | 0.748 / 0.945 (72%) |
| TCGA-LUAD | 0.703 / 0.858 (77%) | 0.737 / 0.827 (86%) | 0.752 / 0.841 (86%) | 0.744 / 0.834 (86%) | 0.774 / 0.860 (86%) | 0.767 / 0.854 (86%) | 0.781 / 0.865 (86%) | 0.796 / 0.877 (86%) |
| CPTAC-LUAD | 0.641 / 0.852 (66%) | 0.676 / 0.786 (84%) | 0.690 / 0.804 (83%) | 0.684 / 0.803 (83%) | 0.713 / 0.826 (83%) | 0.704 / 0.817 (83%) | 0.721 / 0.833 (83%) | 0.735 / 0.845 (83%) |
| TCGA-LUSC | 0.689 / 0.836 (70%) | 0.721 / 0.793 (83%) | 0.737 / 0.810 (82%) | 0.728 / 0.800 (83%) | 0.761 / 0.829 (82%) | 0.753 / 0.822 (83%) | 0.767 / 0.835 (82%) | 0.781 / 0.843 (83%) |
| CPTAC-LUSC | 0.596 / 0.856 (60%) | 0.629 / 0.767 (77%) | 0.647 / 0.772 (78%) | 0.639 / 0.777 (77%) | 0.661 / 0.785 (78%) | 0.659 / 0.791 (77%) | 0.669 / 0.799 (77%) | 0.689 / 0.818 (77%) |

*Continued on next page*

| Cohort | Path | +Stage/ Grade | +TME | +Gene | +Stage/ Grade +TME | +Stage/ Grade +Gene | +TME +Gene | +Stage/ Grade +TME +Gene |
|---|---|---|---|---|---|---|---|---|
| TCGA-nCCRCC | 0.841 / 0.948 (84%) | 0.874 / 0.945 (89%) | 0.889 / 0.955 (88%) | 0.882 / 0.949 (89%) | 0.915 / 0.972 (88%) | 0.905 / 0.963 (89%) | 0.923 / 0.977 (88%) | 0.935 / 0.979 (89%) |
| CPTAC-nCCRCC | 0.731 / 0.965 (84%) | 0.721 / 0.939 (84%) | 0.735 / 0.938 (87%) | 0.728 / 0.948 (83%) | 0.750 / 0.949 (87%) | 0.747 / 0.961 (84%) | 0.751 / 0.958 (85%) | 0.776 / 0.973 (84%) |
| TCGA-PAAD | 0.663 / 0.862 (77%) | 0.696 / 0.795 (89%) | 0.705 / 0.806 (89%) | 0.697 / 0.795 (89%) | 0.729 / 0.824 (89%) | 0.728 / 0.823 (89%) | 0.736 / 0.833 (89%) | 0.753 / 0.846 (89%) |
| CPTAC-PAAD | 0.552 / 0.795 (64%) | 0.587 / 0.724 (81%) | 0.604 / 0.740 (81%) | 0.596 / 0.729 (81%) | 0.618 / 0.748 (82%) | 0.613 / 0.747 (81%) | 0.624 / 0.759 (81%) | 0.645 / 0.769 (82%) |
| TCGA-UCEC | 0.848 / 0.904 (87%) | 0.830 / 0.852 (89%) | 0.831 / 0.866 (89%) | 0.852 / 0.900 (89%) | 0.860 / 0.893 (89%) | 0.874 / 0.891 (89%) | 0.876 / 0.907 (89%) | 0.872 / 0.903 (89%) |
| CPTAC-UCEC | 0.690 / 0.897 (73%) | 0.718 / 0.747 (75%) | 0.713 / 0.758 (76%) | 0.726 / 0.761 (75%) | 0.733 / 0.769 (76%) | 0.749 / 0.774 (75%) | 0.749 / 0.800 (75%) | 0.733 / 0.887 (75%) |
| **Therapeutic response prediction (OR; 6 cohorts)** | | | | | | | | |
| Yale HER2+ | 7.2 / 16 (58%) | 16 / 22 (73%) | 15 / 22 (50%) | 17 / 30 (38%) | 15 / 63 (65%) | 15 / 21 (35%) | 21 / 38 (46%) | 53 / 62 (62%) |
| IMPRESS HER2+ | 7.5 / 12 (50%) | 33 / 120 (50%) | 25 / 25 (42%) | 26 / 31 (47%) | 32 / 150 (45%) | 44 / 67 (39%) | 26 / 34 (55%) | 47 / 60 (52%) |
| TransNEO HER2+ | 5.7 / 9.5 (53%) | 9.8 / 40 (66%) | 15 / 33 (75%) | 19 / 53 (58%) | 17 / 66 (71%) | 17 / 70 (75%) | 15 / 17 (96%) | 21 / 52 (84%) |
| BEV-OV | 25 / 230 (79%) | 35 / 49 (97%) | 44 / 50 (97%) | 43 / 49 (98%) | 42 / 65 (97%) | 41 / 69 (98%) | 40 / 67 (97%) | 48 / 45 (97%) |
| PDL1-NSCLC | 8.1 / 120 (79%) | 18 / 18 (98%) | 20 / 20 (98%) | 19 / 19 (98%) | 14 / 14 (98%) | 21 / 21 (98%) | 21 / 21 (98%) | 15 / 15 (98%) |
| CRT-CESC | 40 / 96 (80%) | 73 / 73 (100%) | 65 / 65 (100%) | 62 / 62 (100%) | 94 / 94 (100%) | 76 / 76 (100%) | 69 / 69 (100%) | 94 / 94 (100%) |

**Supplementary Table 121. Paired differences between TEAM and the best-performing baseline of each evaluation unit.** n, evaluation units; CIs from 1,000 unit-level bootstrap resamples; $P$, two-sided Wilcoxon signed-rank test.

| Task | Metric | n | Mean paired difference (95% CI) | Wilcoxon $P$ |
|---|---|---|---|---|
| Prognostic stratification | C-index | 31 | 0.033 (0.024–0.042) | $1.17 \times 10^{-6}$ |
| Therapeutic response prediction | Log OR | 6 | 0.910 (0.628–1.287) | 0.031 |
| Cancer staging/grading | AUC | 20 | 0.023 (0.016–0.031) | $1.91 \times 10^{-6}$ |
| TME characterization | PCC | 16 | 0.047 (0.042–0.052) | $3.05 \times 10^{-5}$ |
| Gene expression prediction | Gene-wise PCC | 17 | 0.026 (0.014–0.039) | $1.53 \times 10^{-5}$ |

**Supplementary Table 122. Sensitivity to $T$ by task.** Cost, total inference time relative to $T = 3$.

| T | Prognostic stratification, C-index (31 cohorts) | Therapeutic response prediction, Log OR / AP (6 cohorts) | Relative cost |
|---|---|---|---|
| 3 | 0.701 | 2.46 / 0.830 | 1.00× |
| 5 | 0.706 | 2.68 / 0.836 | 1.09× |
| 20 | 0.708 | 2.64 / 0.847 | 1.84× |

**Supplementary Table 123. Performance of different vision-language models on the QuiltVQA dataset for visual question answering.** Comparison of vision-language models on QuiltVQA closed-ended questions. Performance is measured using accuracy and F1-score (%). TEAM-based variants integrate TEAM's visual encoder with different language models.

| Model | Accuracy (%) | F1-Score (%) |
|---|---|---|
| Quilt-Llava | 62.97 | 73.38 |
| TEAM-Llava | 72.89 | 84.32 |
| DeepSeek-R1 | – | – |
| TEAM-DeepSeek | 68.22 | 71.39 |
| GPT-4o | 55.10 | 65.50 |
| TEAM-Agent | 65.26 | 73.17 |

**Supplementary Table 124. Performance of different vision-language models on the PathVQA dataset for visual question answering.** Comparison of vision-language models on PathVQA closed-ended questions. Performance is measured using accuracy and F1-score (%). TEAM-based variants integrate TEAM's visual encoder with different language models.

| Model | Accuracy (%) | F1-Score (%) |
|---|---|---|
| Quilt-Llava | 54.85 | 63.40 |
| TEAM-Llava | 54.41 | 70.36 |
| DeepSeek-R1 | – | – |
| TEAM-DeepSeek | 62.02 | 75.33 |
| GPT-4o | 63.49 | 53.49 |
| TEAM-Agent | 82.22 | 81.12 |

**Supplementary Table 125. Performance of different vision-language models on the PathMMU-EduContent dataset for visual question answering.** Comparison of vision-language models on PathMMU educational questions. Performance is measured using accuracy and F1-score (%). TEAM-based variants integrate TEAM's visual encoder with different language models.

| Model | Accuracy (%) | F1-Score (%) |
|---|---|---|
| Quilt-Llava | 34.90 | 33.92 |
| TEAM-Llava | 50.20 | 49.46 |
| DeepSeek-R1 | – | – |
| TEAM-DeepSeek | 55.30 | 55.39 |
| GPT-4o | 56.08 | 53.14 |
| TEAM-Agent | 71.37 | 71.18 |

**Supplementary Table 126. Performance of different vision-language models on the PathMMU-PubMed dataset for visual question answering.** Comparison of vision-language models on PathMMU-PubMed questions. Performance is measured using accuracy and F1-score (%). TEAM-based variants integrate TEAM's visual encoder with different language models.

| Model | Accuracy (%) | F1-Score (%) |
|---|---|---|
| Quilt-Llava | 33.81 | 30.90 |
| TEAM-Llava | 48.75 | 48.70 |
| DeepSeek-R1 | – | – |
| TEAM-DeepSeek | 62.98 | 62.97 |
| GPT-4o | 59.07 | 57.79 |
| TEAM-Agent | 69.40 | 69.37 |

**Supplementary Table 127. Overall performance of vision-language models evaluated by clinicians using Borda count method.** Five pathologists ranked the model responses to 260 questions across 52 pathology images (5 questions per image) on a 1–5 scale (5 = best), with tied responses sharing the mean rank. Model rankings were aggregated using the Borda count method, which assigns points based on relative rankings across evaluators. TEAM-based variants integrate TEAM's visual encoder with different language models (Llava, DeepSeek-R1, GPT-4o). Statistics are computed across all evaluations (260 questions × 5 pathologists = 1,300 evaluations per model). 95% CIs were calculated using the t-distribution.

| **Model** | **N** | **Mean** | **s.d.** | **95% CI** |
|---|---|---|---|---|
| Quilt-Llava | 1,300 | 2.75 | 1.13 | (2.69–2.81) |
| TEAM-Llava | 1,300 | 3.03 | 1.17 | (2.96–3.09) |
| DeepSeek-R1 | – | – | – | – |
| TEAM-DeepSeek | 1,300 | 2.64 | 1.16 | (2.57–2.70) |
| GPT-4o | 1,300 | 2.42 | 1.30 | (2.35–2.49) |
| TEAM-Agent | 1,300 | 4.00 | 1.25 | (3.93–4.07) |

**Supplementary Table 128. Head-to-head comparison of TEAM-based variants against baseline models.** Each TEAM model variant was compared against baseline models across 1,300 evaluations (260 questions × 5 pathologists). Win/Tie/Loss percentages indicate the proportion of evaluations where the TEAM model was rated better/equal/worse than the baseline. Win rates among non-tied evaluations, Win/(Win+Loss), are 86.0% for TEAM-Agent versus GPT-4o and 57.7% for TEAM-Llava versus Quilt-Llava. Statistical significance was assessed using two-sided Wilcoxon signed-rank tests. ***$P$<0.001; **$P$<0.01; ns, not significant.

| **TEAM Model** | **Baseline** | **Win (%)** | **Tie (%)** | **Loss (%)** | **Significance** |
|---|---|---|---|---|---|
| *TEAM-Llava Performance* | | | | | |
| TEAM-Llava | Quilt-Llava | 39.2 | 32.0 | 28.8 | $P$<0.001*** |
| TEAM-Llava | GPT-4o | 52.0 | 21.2 | 26.8 | $P$<0.001*** |
| *TEAM-DeepSeek Performance* | | | | | |
| TEAM-DeepSeek | Quilt-Llava | 31.8 | 30.2 | 38.1 | ns |
| TEAM-DeepSeek | GPT-4o | 46.2 | 19.5 | 34.3 | $P$=0.001** |
| *TEAM-Agent Performance* | | | | | |
| TEAM-Agent | Quilt-Llava | 68.2 | 15.8 | 16.0 | $P$<0.001*** |
| TEAM-Agent | GPT-4o | 70.3 | 18.2 | 11.5 | $P$<0.001*** |

**Supplementary Table 129. Quantitative assessment of clinician-in-the-loop refinement across clinical tasks.** Performance before (Baseline) and after (Refined) pathologist-guided refinement on the 40 refined cases per cohort (TCGA-CESC, CPTAC-KIRC, and BEV-OV; Supplementary Table 130). Metrics: C-index for prognostic stratification, OR for therapeutic response prediction, AUC for cancer staging, and PCC for TME characterization and gene expression prediction. Refined, mean across five pathologists; Gain, change from Baseline.

| **Clinical Task** | **Cohort** | **Endpoint** | **Baseline** | **Refined** | **Gain (Δ)** |
|---|---|---|---|---|---|
| Prognosis | TCGA-CESC | Survival | 0.936 | 0.989 | +0.053 |
| | CPTAC-KIRC | Survival | 0.708 | 0.773 | +0.065 |
| Therapeutic Response | BEV-OV | Response | 3.575 | 8.190 | +4.614 |
| Cancer Staging | TCGA-CESC | Stage | 0.724 | 0.783 | +0.058 |
| | CPTAC-KIRC | Stage | 0.635 | 0.730 | +0.096 |
| TME Biomarkers | TCGA-CESC | PD-L1 | 0.878 | 0.934 | +0.056 |
| | TCGA-CESC | CD8/CD4 | 0.772 | 0.813 | +0.041 |
| | TCGA-CESC | TILs | 0.926 | 0.943 | +0.017 |
| | TCGA-CESC | Ki-67 | 0.765 | 0.825 | +0.059 |
| | TCGA-CESC | TMB | 0.600 | 0.641 | +0.041 |
| | TCGA-CESC | MSI | 0.135 | 0.195 | +0.060 |
| | TCGA-CESC | HRD | 0.775 | 0.797 | +0.022 |
| | TCGA-CESC | Hypoxia | 0.264 | 0.296 | +0.031 |
| | CPTAC-KIRC | PD-L1 | 0.132 | 0.193 | +0.061 |
| | CPTAC-KIRC | CD8/CD4 | 0.162 | 0.215 | +0.052 |
| | CPTAC-KIRC | TILs | 0.187 | 0.228 | +0.041 |
| | CPTAC-KIRC | Ki-67 | 0.217 | 0.275 | +0.058 |
| | CPTAC-KIRC | TMB | 0.269 | 0.297 | +0.028 |
| | CPTAC-KIRC | MSI | 0.217 | 0.276 | +0.059 |
| | CPTAC-KIRC | HRD | 0.167 | 0.189 | +0.022 |
| | CPTAC-KIRC | Hypoxia | 0.242 | 0.271 | +0.029 |
| Gene Expression | TCGA-CESC | All genes | 0.327 | 0.414 | +0.087 |
| | TCGA-CESC | Top 1,000 | 0.346 | 0.433 | +0.087 |
| | CPTAC-KIRC | All genes | 0.068 | 0.142 | +0.074 |
| | CPTAC-KIRC | Top 1,000 | 0.123 | 0.196 | +0.073 |

**Supplementary Table 130. Pathologist interaction frequency during the model refinement process.** Summary statistics (mean ± s.d.) of refinement iterations per pathologist (P1–P5) across three cohorts (TCGA-CESC, CPTAC-KIRC, BEV-OV), with $n = 40$ cases per cohort.

| **Cohort** | **P1** | **P2** | **P3** | **P4** | **P5** |
|---|---|---|---|---|---|
| TCGA-CESC | $2.12 \pm 0.40$ | $1.73 \pm 0.60$ | $1.90 \pm 0.50$ | $3.10 \pm 1.22$ | $1.07 \pm 0.27$ |
| CPTAC-KIRC | $2.85 \pm 1.12$ | $2.08 \pm 0.66$ | $2.12 \pm 0.72$ | $2.50 \pm 0.88$ | $2.08 \pm 1.00$ |
| BEV-OV | $2.55 \pm 0.75$ | $2.17 \pm 0.75$ | $1.85 \pm 0.66$ | $2.38 \pm 0.70$ | $1.95 \pm 0.45$ |
| **Overall** | $2.51 \pm 0.86$ | $1.99 \pm 0.69$ | $1.96 \pm 0.64$ | $2.66 \pm 1.00$ | $1.70 \pm 0.78$ |

**Supplementary Table 131. Cancer type abbreviations.** Abbreviations of cancer types used in the main and Extended Data figures.

| Abbreviation | Cancer type |
|---|---|
| BLCA | Bladder urothelial carcinoma |
| BRCA | Breast cancer |
| CESC | Cervical squamous cell carcinoma |
| CHOL | Cholangiocarcinoma |
| COAD | Colon adenocarcinoma |
| CRC | Colorectal cancer |
| DLBC | Lymphoid neoplasm diffuse large B-cell lymphoma |
| EAC | Esophageal adenocarcinoma |
| ESCA | Esophageal carcinoma |
| ESCC | Esophageal squamous cell carcinoma |
| ESC-RCC | Eosinophilic solid and cystic renal cell carcinoma |
| GBM | Glioblastoma multiforme |
| GC | Gastric carcinoma |
| HNSC | Head and neck squamous cell carcinoma |
| KICH | Kidney chromophobe |
| KIRC | Kidney renal clear cell carcinoma |
| KIRP | Kidney renal papillary cell carcinoma |
| LAML | Acute myeloid leukemia |
| LGG | Brain lower grade glioma |
| LIHC | Liver hepatocellular carcinoma |
| LUAD | Lung adenocarcinoma |
| LUSC | Lung squamous cell carcinoma |
| MESO | Mesothelioma |
| nCCRCC | Non-clear cell renal cell carcinoma |
| NSCLC | Non-small cell lung cancer |
| OV | Ovarian cancer |
| PAAD | Pancreatic adenocarcinoma |
| PRAD | Prostate adenocarcinoma |
| READ | Rectum adenocarcinoma |
| SARC | Sarcoma |
| SKCM | Skin cutaneous melanoma |
| STAD | Stomach adenocarcinoma |
| TGCT | Testicular germ cell tumors |
| THCA | Thyroid carcinoma |
| THYM | Thymoma |
| UCEC | Uterine corpus endometrial carcinoma |
| UCS | Uterine carcinosarcoma |
| UVM | Uveal melanoma |
